# Combining Transdiagnostic and Disorder-Level GWAS Enhances Precision of Psychiatric Genetic Risk Profiles in a Multi-Ancestry Sample

**DOI:** 10.1101/2024.05.09.24307111

**Authors:** Yousef Khan, Christal N. Davis, Zeal Jinwala, Kyra L. Feuer, Sylvanus Toikumo, Emily E. Hartwell, Sandra Sanchez-Roige, Roseann E. Peterson, Alexander S. Hatoum, Henry R. Kranzler, Rachel L. Kember

## Abstract

The etiology of substance use disorders (SUDs) and psychiatric disorders reflects a combination of both transdiagnostic (i.e., common) and disorder-level (i.e., independent) genetic risk factors. We applied genomic structural equation modeling to examine these genetic factors across SUDs, psychotic, mood, and anxiety disorders using genome-wide association studies (GWAS) of European-(EUR) and African-ancestry (AFR) individuals. In EUR individuals, transdiagnostic genetic factors represented SUDs (143 lead single nucleotide polymorphisms [SNPs]), psychotic (162 lead SNPs), and mood/anxiety disorders (112 lead SNPs). We identified two novel SNPs for mood/anxiety disorders that have probable regulatory roles on *FOXP1*, *NECTIN3*, and *BTLA* genes. In AFR individuals, genetic factors represented SUDs (1 lead SNP) and psychiatric disorders (no significant SNPs). The SUD factor lead SNP, although previously significant in EUR- and cross-ancestry GWAS, is a novel finding in AFR individuals. Shared genetic variance accounted for overlap between SUDs and their psychiatric comorbidities, with second-order GWAS identifying up to 12 SNPs not significantly associated with either first-order factor in EUR individuals. Finally, common and independent genetic effects showed different associations with psychiatric, sociodemographic, and medical phenotypes. For example, the independent components of schizophrenia and bipolar disorder had distinct associations with affective and risk-taking behaviors, and phenome-wide association studies identified medical conditions associated with tobacco use disorder independent of the broader SUDs factor. Thus, combining transdiagnostic and disorder-level genetic approaches can improve our understanding of co-occurring conditions and increase the specificity of genetic discovery, which is critical for psychiatric disorders that demonstrate considerable symptom and etiological overlap.

## Introduction

Substance use disorders (SUDs) commonly co-occur with mood, anxiety, and psychotic disorders.^1–3^ For example, about one-quarter of individuals with major depressive disorder (MDD) meet criteria for at least one SUD,^1^ and 20-80% of individuals seeking SUD treatment have MDD.^4^ Similarly, the prevalence of anxiety disorders among individuals with an illicit SUD is almost three times that of the general population.^2^ Additionally, nearly half (42%) of individuals experiencing a first episode of psychosis^5^ and a third of those with bipolar disorder^3^ (BD) have a co-occurring SUD. Such comorbidity complicates the clinical course of affected individuals, resulting in greater healthcare and other costs.^6,7^

Large-scale genome-wide association studies (GWAS)^8–15^ have shown these disorders to be highly polygenic, with individual variants exerting a small influence on risk. These studies also provide evidence of pleiotropy, whereby variants are associated with more than one psychiatric trait. These pleiotropic effects partially account for the co-occurrence of psychiatric traits and disorders.^8–15^ Genomic structural equation modeling (gSEM) can leverage this shared liability to identify common genetic factors that underlie multiple disorders.^16^ In combination with downstream analyses, these multivariate genetic approaches can improve our understanding of the etiology of commonly co-occurring conditions by identifying shared biological pathways and possible risk mechanisms.

Several gSEM studies have modeled pleiotropy across psychiatric disorders in European-ancestry (EUR) individuals. The first modeled shared genetic variance across schizophrenia (SCZ), BD, MDD, posttraumatic stress disorder (PTSD), and anxiety, identifying a single common factor.^16^ Across eight psychiatric disorders—attention-deficit hyperactivity disorder, anorexia nervosa, autism spectrum disorder, BD, MDD, obsessive compulsive disorder, SCZ, and Tourette syndrome^17^–three underlying genetic factors representing mood and psychotic disorders, early-onset neurodevelopmental disorders, and compulsive disorders—best represented the data. More recent work expanded this model by adding problematic alcohol use, anxiety disorders, and PTSD,^18^ which resulted in the previously combined mood and psychotic disorders factor splitting into separate internalizing and psychotic factors.

Similarly, an addiction factor has been identified that underlies cannabis use disorder (CanUD), opioid use disorder (OUD), and measures of problematic alcohol and tobacco use.^19^ Although this study included African-ancestry (AFR) in addition to EUR individuals, limited power precluded the application of gSEM in AFR individuals. Instead, an alternative approach, ASSET,^20^ was taken to identify pleiotropic effects among AFR individuals. However, this approach failed to identify any variants having effects common to all four substance use behaviors in AFR individuals. Other research applying gSEM has derived a common factor underlying measures of substance use in EUR individuals,^21^ reaffirming the shared genetic basis underlying multiple substance use behaviors or disorders.

Although research has demonstrated consistency in the genetic factor structure across models of substance use and psychiatric disorders, these studies have applied gSEM only in EUR individuals. The modeling of complex genetic relationships via structural equation modeling imposes greater demands for statistical power than simpler univariate GWAS analyses, which themselves are often underpowered in AFR and other non-EUR ancestry groups. Thus, the statistical power requirements of the approach, whose strength lies in its ability to enhance our understanding of the complexity of genetic relations, have hampered its application in non-EUR ancestries.

Previous studies have also typically focused on identifying transdiagnostic genetic risk, but gSEM can also enable more precise identification of disorder-specific genetic mechanisms than individual GWAS. In combination with transdiagnostic genetic approaches, GWAS-by-subtraction^22^ can parse associations with single nucleotide polymorphisms (SNPs) into those that influence risk for a disorder through a common genetic factor from those that operate independently of the common factor. The two resulting genetic dimensions can be used to differentiate the associations of a common genetic factor with psychiatric, medical, and social phenotypes from the associations of genetic risk that operates independently of the common factor. Combining transdiagnostic and disorder-level approaches enhances statistical power to detect pleiotropic effects, while identifying patterns of genetic heterogeneity and increasing the specificity of SNP discovery.^23^

To extend previous study findings, we used gSEM to characterize the underlying genetic structure of SUDs, psychotic, and mood and anxiety disorders in EUR and AFR individuals. First, we examined the genetic factor structure of the disorders using exploratory and confirmatory factor analyses, and then we explored each factor’s biological underpinnings by conducting GWAS. Next, we investigated the shared genetic comorbidity of SUDs with psychotic, mood, and anxiety disorders using a second-order gSEM approach, which involves examining the relationships between the lower-level factors to identify genetic risks that are shared across factors. Finally, we characterized the common and independent genetic variance for select disorders using GWAS-by-subtraction. Thus, we sought to address two critical gaps in previous research: first, by applying gSEM among AFR individuals, and second, by employing a hierarchical approach to explore both genetic specificity and transdiagnostic genetic risk.

Ultimately, these techniques were applied to obtain a more comprehensive understanding of the genetic underpinnings of SUDs, psychotic, mood, and anxiety disorders.

## Methods

To explore the genetic relationships among SUDs, psychotic, mood, and anxiety disorders, we used gSEM, which is a statistical technique that integrates summary-level genetic data with structural equation modeling. By estimating the genetic covariance structure among traits, gSEM enables the explicit and flexible modeling of complex relationships.^16^

**Figure 1.**
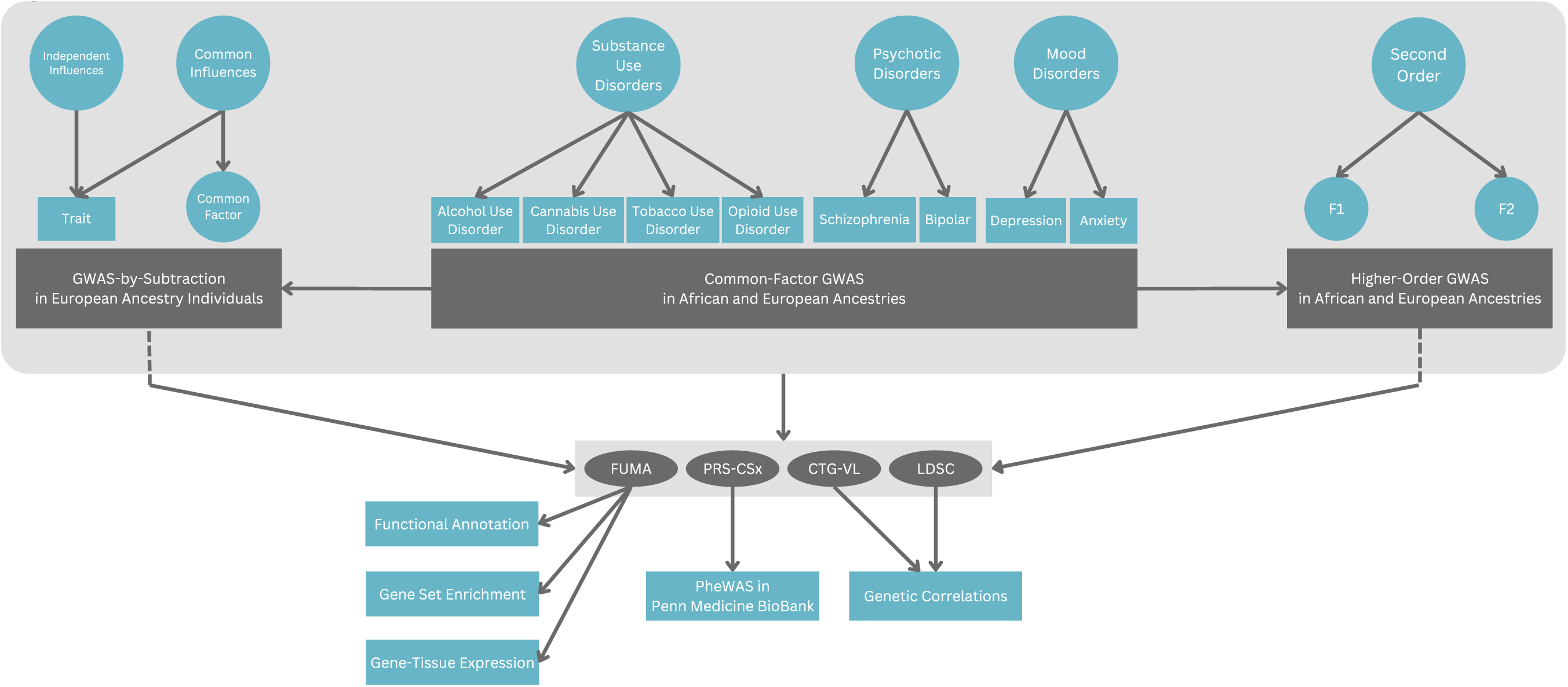
Study schema. FUMA = Functional Mapping and Annotation of GWAS, CTG-VL = Complex Trait Genetics Virtual Lab, LDSC = linkage disequilibrium score regression, PheWAS = phenome-wide association study.

## Summary Statistics

We used large discovery GWAS for SUDs, psychotic, mood, and anxiety disorders in EUR and AFR individuals (Supplementary Tables 1-4) as inputs for gSEM. Genetic ancestry was determined for each input GWAS by the researchers conducting the original study, and these inferences should not be considered proxies for either race or ethnicity. Instead, the groups represent statistical determinations of ancestral similarity, within which there remains heterogeneity. The traits included four SUDs: alcohol use disorder (AUD),^13,24^ tobacco use disorder (TUD),^15^ cannabis use disorder (CanUD),^12^ and opioid use disorder (OUD);^11^ two disorders that can include psychotic features: BD^10,25^ and SCZ;^9,25^ and two mood and anxiety traits: anxiety (ANX)^8,26,27^ and MDD.^14,28^ In EUR individuals, we used multi-trait analysis of GWAS (MTAG)^29^ to enhance the power to detect effects associated with a broad spectrum of anxiety disorders, including generalized anxiety disorder, panic disorder, social phobia, agoraphobia, and specific phobias (see Supplementary Materials and Supplementary Table 5). The resulting ANX summary statistics produced by MTAG were then used as an input for gSEM.

**Table 1.** Genome-wide association studies from which summary statistics were obtained.

| **European Ancestry** |  |  |  |  |  |
| --- | --- | --- | --- | --- | --- |
| **Study** | **Phenotype** | **Type** | **Cases** | **Controls** | **Total** |
| Als et al., 2023, *Nature Medicine* | MDD | Case-control | 387,429 | 976,554 | 1,363,983 |
| Otowa et al., 2016, *Molecular Psychiatry* * | ANX | Case-control | 7,016 | 14,745 | 21,761 |
| Levey et al., 2020, *American Journal of Psychiatry* * | ANX | Continuous |  |  | 175,163 |
| Purves et al., 2020, *Molecular Psychiatry* * | ANX | Case-Control | 25,453 | 58,113 | 83,566 |
| Trubetskoy et al., 2022, *Nature* | SCZ | Case-control | 53,386 | 77,258 | 130,644 |
| Mullins et al., 2021, *Nature Genetics* | BD | Case-control | 41,917 | 371,549 | 413,466 |
| Zhou et al., 2023, *Nature Medicine* | AUD | Case-control | 113,325 | 639,923 | 753,248 |
| Levey et al., 2023 *Nature Genetics* | CanUD | Case-control | 42,281 | 843,744 | 886,025 |
| Toikumo et al., 2024, *Nature Human Behaviour* | TUD | Case-control | 163,734 | 331,271 | 495,005 |
| Kember et al., 2022, *Nature Neuroscience* | ODU | Case-control | 31,473 | 394,471 | 425,944 |
| **African-Ancestry** |  |  |  |  |  |
| Levey et al., 2021, *Nature Neuroscience* | MDD | Case-control | 25,843 | 33,757 | 59,600 |
| Levey et al., 2021, *American Journal of Psychiatry* | ANX | Continuous |  |  | 24,448 |
| Bigdeli et al., 2021, *Schizophrenia Bulletin* | SCZ | Case-control | 7,509 | 8,612 | 16,121 |
| Bigdeli et al., 2021, *Schizophrenia Bulletin* | BD | Case-control | 3,027 | 8,097 | 11,124 |
| Kember et al., 2023, *American Journal of Psychiatry* | AUD | Case-control | 25,012 | 52,313 | 77,325 |
| Levey et al., 2023, *Nature Genetics* | CanUD | Case-control | 19,065 | 104,143 | 123,208 |
| Toikumo et al., 2024, *Nature Human Behaviour* | TUD | Case-control | 45,465 | 68,955 | 114,420 |
| Kember et al., 2022, *Nature Neuroscience* | ODU | Case-control | 8,968 | 79,530 | 88,498 |
| *Note:* MDD = Major Depressive Disorder, ANX = Anxiety Disorders, SCZ = Schizophrenia, BD = Bipolar Disorder, AUD = Alcohol Use Disorder, CanUD = Cannabis Use Disorder, TUD = Tobacco Use Disorder, OUD = Opioid Use Disorder. See Supplementary Tables 1-4 for full description of cohorts.<br>*Summary statistics were jointly analyzed using MTAG to enhance the statistical power of the ANX phenotype prior to their being used in gSEM (see Supplementary Materials). |  |  |  |  |  |

Prior to common factor modeling in accordance with the procedure for performing gSEM, we calculated genetic correlations between the disorders using linkage disequilibrium score regression (LDSC) implemented in GenomicSEM 0.0.5c.^16^ To prevent downward bias in LDSC estimates, when SNP-level sample sizes were not available within each set of summary statistics, we calculated the effective sample size using the sum of effective sample sizes across the input GWAS cohorts (see Supplementary Materials).^30^ For EUR analyses, SNPs were restricted to those contained within the EUR HapMap3 reference panel^31^ with a minor allele frequency (MAF) > 0.01. We then performed LDSC using EUR 1000 Genomes Phase 3 linkage disequilibrium (LD) scores.^32^ Given the statistical challenges associated with including non-EUR individuals in gSEM analyses due in part to differences in LD structure and admixture, we compared the performance of three sets of LD scores before performing LDSC in AFR individuals. We restricted each set of LD scores to well-imputed SNPs with MAF > 0.01 and then compared several parameters, including LD score distribution, LD block length, and the stability of SNP-based heritability and genetic correlation estimates to determine the optimal approach (see Supplementary Materials). This step ensured the selection of an appropriately matched reference panel to avoid biasing estimates. Ultimately, we selected the Pan-UKB AFR reference data,^33^ which enabled the best performance of gSEM models among AFR individuals.

### Genomic Structural Equation Modeling

Consistent with best approaches for gSEM, we performed exploratory factor analysis (EFA) and confirmatory factor analysis (CFA) on independent data to evaluate the reliability of our results. We first performed EFA on the odd chromosomes using LDSC matrices derived from the summary statistics of the previously described SUDs and psychiatric disorders to evaluate the optimal number of factors and the loadings of each disorder in a hypothesis-free manner. We used the *Matrix* R package to correct for the possibility of a non-positive definite matrix^34^ within the LDSC output and then performed EFA for 1-4 latent factors using the *lavaan* R package and a promax rotation.^35^ Following EFA, we examined model fit indices (i.e., chi-square value, Akaike information criterion (AIC), comparative fix index (CFI), and standardized root mean squared residual (SRMR)) and eigenvalues to determine the optimal model.^36^ Traits with a loading ≥0.35 on each latent factor were retained for confirmatory factor analysis (CFA), which was performed on the even chromosomes to avoid overfitting the data. Because of the limited statistical power in AFR models, factor analyses were performed on all chromosomes.

Following CFA, we prepared the input summary statistics for GWAS by standardizing coefficients and standard error (SE) values, such that SNP effects were scaled similarly for binary and continuous phenotypes. For quality control, we included only SNPs with MAF>0.01 and imputation score>0.60. In GWAS, we regressed each SNP on each latent variable using diagonally weighted least squares (DWLS) estimation and standard genomic control. We calculated the effective sample size of each resulting common factor GWAS as described by Mallard et al. (2022).^37^

Next, we constructed second-order common factor models to capture genetic effects that account for co-occurrence among SUDs and their psychiatric comorbidities. For both the EUR and AFR analyses, to ensure identification of the second-order models, we set the loadings of each first-order factor onto its respective second-order factor equal to the square root of their genetic correlation.^38^ We then ran GWAS on each second-order factor using the procedure described for the first-order common factor GWAS.

To identify significant independent SNPs from GWAS, we performed LD clumping with PLINK 1.9^39^ using an r^2^ threshold of 0.1 and physical distance threshold of 3000 kb(Supplementary Materials). For loci not previously significantly associated with a corresponding SUD, psychotic, mood, or anxiety disorder (hereafter referred to as novel), we performed PheWAS on the lead SNP to examine its associations across the phenomic spectrum using the GWAS Atlas (Supplementary Materials).^40^ Additionally, for any novel lead SNPs identified, we used the LD-based Probabilistic Identification of Causal SNPs (PICS) v2.1.1 finemapping tool to assess their potential as the most likely causal variant to be responsible for the observed association in a given locus.

After performing each of the first- and second-order common factor GWAS, we calculated Q_SNP_, a measure of heterogeneity that tests the null hypothesis that a SNP’s effects operate entirely through a common factor. For example, a SNP that primarily influences SUDs through its effects on a single disorder, like TUD, should violate the null hypothesis. To identify SNPs with heterogeneous effects, we examined associations between each SNP and common factor via a common pathway model. Then, separately for each factor, we fit an independent pathway model in which the SNP predicted each of the factor’s indicators. We performed a chi-square difference test on the two models (Supplementary Figure 1) and removed SNPs with *p* < 5 x 10^-8^ (Supplementary Tables 6-10) from the factor’s summary statistics prior to conducting all post-GWAS analyses.

### GWAS-by-Subtraction

To ensure that the GWAS-by-subtraction models were informative and adequately powered, we performed these analyses only on disorders with a standardized unexplained variance ≥ 0.30 in the first-order CFA. Following the paradigm of Demange et al. (2021),^22^ we first specified two latent genetic factors. On the first factor, we loaded only the specific disorder of interest (i.e., depending on the model, TUD, BD, or SCZ), while on the other factor we loaded the specific disorder *and* the other disorders included in its broader common factor. Then, we imposed two constraints by setting to 0 the covariance between: (1) the disorder of interest and all other disorders, and (2) the two latent genetic factors. Finally, to capture genetic effects on the disorder that were not correlated with the common factor, we regressed each SNP on the latent factor with a sole loading for the disorder of interest. To identify genetic effects on the disorder of interest that operated through the common factor, each SNP was regressed on the latent factor on which all common factor traits were loaded. Due to the limited statistical power of the AFR models (max N_eff_ = 6,421), we restricted GWAS-by-subtraction analyses to EUR models. The effective sample size calculations for each of the GWAS-by-subtraction models were adjusted to account for the fact that the GWAS modeled residual heritability.^22^

### Biological Characterization

We used Functional Mapping and Annotation of Genome-Wide Association Studies (FUMA) version 1.6.0^41^ to conduct post-GWAS analyses of each GWAS (i.e., first-order common factors in EUR and AFR individuals, second-order common factors in EUR and AFR individuals, and GWAS-by-subtraction models in EUR individuals). Gene-based tests, gene-set enrichment, and gene-tissue expression analyses were conducted using MAGMA version 1.08.^42^ We examined gene expression in BrainSpan^43^ and GTEx v8^44^ tissue samples. SNP-to-gene associations and gene annotations were evaluated using: (1) expression quantitative trait loci (eQTLs) from PsychENCODE^45^ and GTEx v8^44^ brain tissue samples and (2) chromatin interactions via Hi-C data for the dorsolateral prefrontal cortex, hippocampus, ventricles, and neural progenitor cells.^46^ To annotate the protein products and investigate protein-protein interactions of MAGMA-identified genes, we used the STRING database v12.0 and applied its default parameters.^47^ Enrichment of protein-protein interactions was calculated as the observed number of edges (i.e., interactions) divided by the expected number of edges in the protein network. Significant enrichment would suggest that the proteins that are encoded by genes associated with a factor participate in common pathways relevant to that factor.

### Genetic Correlations with gSEM Factors

Genetic correlations between gSEM output summary statistics and other traits were calculated using LDSC^48,49^ with 1000 Genomes Project phase 3^32^ (for EUR) and PanUKB^33^ (for AFR) data as LD references. For first- and second-order common factors in EUR individuals, we used the Complex-Traits Genetics Virtual Lab^50^ to calculate batch genetic correlations with 1,437 traits across a wide variety of domains assessed via International Classification of Diseases (ICD) codes and self-report. In AFR individuals, genetic correlations were calculated for selected psychiatric and medical phenotypes as the Complex-Traits Genetics Virtual Lab does not currently facilitate LDSC in non-EUR ancestries. For GWAS-by-subtraction models in EUR individuals, we calculated genetic correlations with a selection of relevant psychiatric, social, and physical traits to facilitate comparison of the common and independent genetic effects associated with each disorder. We applied a Benjamini-Hochberg false discovery rate (FDR) correction to each set of genetic correlation analyses to account for multiple testing. Lastly, we calculated trans-ancestry genetic correlations using the regression fit method to compare gSEM common factors across EUR and AFR ancestry individuals with Popcorn v1.0^51^ and ancestry-matched 1000 Genomes reference files. Reference files were prepared by excluding the MHC region, and Popcorn was used to compute LD scores for both populations.

### Polygenic Score-Based PheWAS

Prior to calculating polygenic scores (PGS) in the Penn Medicine Biobank (PMBB), GWAS for the first-order, second-order, and TUD GWAS-by-subtraction models were re-run excluding PMBB to ensure independence of the GWAS and target samples. We calculated PGS from gSEM output summary statistics using PRS-CSx^52^ and conducted a phenome-wide association study (PheWAS) in the PMBB. PheWAS is a hypothesis-free approach to explore the association between genetic variants and traits across the spectrum of human disease and health. PMBB participants are recruited through the University of Pennsylvania Health System and provide access to their electronic health record (EHR) and blood or tissue samples.^53^ Genotyping was performed using the Illumina Global Screening Array. Quality control procedures included removing SNPs with marker call rates <95% and sample call rates <90%, as well as individuals with sex discrepancies. Imputation was performed using Eagle2 (Reference-based phasing using the Haplotype Reference Consortium panel) and Minimac4 on the TOPMed Imputation Server. In the case of related individuals (pi-hat threshold ≥ 0.25), one from each pair was removed from analyses. Genetic ancestry was determined using quantitative discriminant analysis of principal components (PCs) using smartpca.^54,55^ These procedures resulted in 10,383 AFR individuals and 29,355 EUR individuals for inclusion in the PheWAS.

ICD-9 and ICD-10 codes were gathered from EHR and mapped to phecodes. Cases were individuals with at least two instances of a given ICD code (“phecodes”). PGS were standardized, and PheWAS was conducted by fitting a logistic regression predicting each phecode from the PGS, with sex, age, and the top 10 PCs included as covariates using the *PheWAS* package^56^ in R. We used a Benjamini-Hochberg FDR corrected p-value to ascertain significant associations.

## Results

### Genetic Correlations among Input GWAS

#### European Ancestry

Genetic correlations among SUDs ranged from 0.60 (SE = 0.06, *p* < 0.001; TUD and OUD) to 0.92 (SE = 0.05, *p* < 0.001; AUD and OUD). MDD and ANX were strongly genetically correlated (*r*_g_ = 0.91, SE = 0.04, *p* < 0.001), as were BD and SCZ (*r*_g_ = 0.68, SE = 0.03, *p* < 0.001). Furthermore, SUDs were significantly genetically correlated with the other psychiatric disorders, ranging from 0.08 (SE = 0.02, *p* < 0.001; TUD and BD) to 0.51 (SE = 0.03, *p* < 0.001; AUD and MDD; Supplementary Figure 2 and Supplementary Table 11).

#### African Ancestry

All four SUDs exhibited significant genetic correlations with one another, ranging from 0.56 (SE = 0.10, *p* < 0.001; TUD and CanUD) to 0.89 (SE = 0.18, *p* < 0.001; OUD and CanUD). MDD and ANX were significantly correlated (*r*_g_ = 0.89, SE = 0.385, *p* = 0.021), as were BD and SCZ (*r*_g_ = 0.43, SE = 0.17, *p* = 0.011). Across disorder classes, AUD was correlated with all the psychiatric disorders except BD (*r*_g_ = 0.19, SE = 0.15, *p* = 0.21). On the other hand, TUD was only significantly genetically correlated with BD (*r*_g_ = 0.24, SE = 0.12, *p* = 0.037) and SCZ (*r*_g_ = 0.29, SE = 0.10, *p* = 0.005). CanUD was significantly genetically correlated with all the psychiatric disorders except ANX (*r*_g_ = 0.29, SE = 0.20, *p* = 0.145), and OUD was correlated with all (Supplementary Figure 3 and Supplementary Table 12).

### First-Order Common Factors: SUDs, Psychotic, and Mood Disorders

#### European Ancestry

Of the EFA models examined, the 3-factor model fit the data best (Supplementary Table 13). As all loadings were ≥ 0.35 with no significant cross-loadings, all traits were carried forward on their respective factors for CFA, which fit the data well (𝜒^2^(17) = 130.04, *p* = 1.84x10^-19^, AIC = 168.04, CFI = 0.95, SRMR = 0.05). All SUDs loaded onto the first factor, while BD and SCZ loaded onto the second, and MDD and ANX onto the third. The factors were significantly intercorrelated, ranging from 0.39 (F1∼F2) to 0.50 (F2∼F3) (Supplementary Table 14).

**Figure 2.**
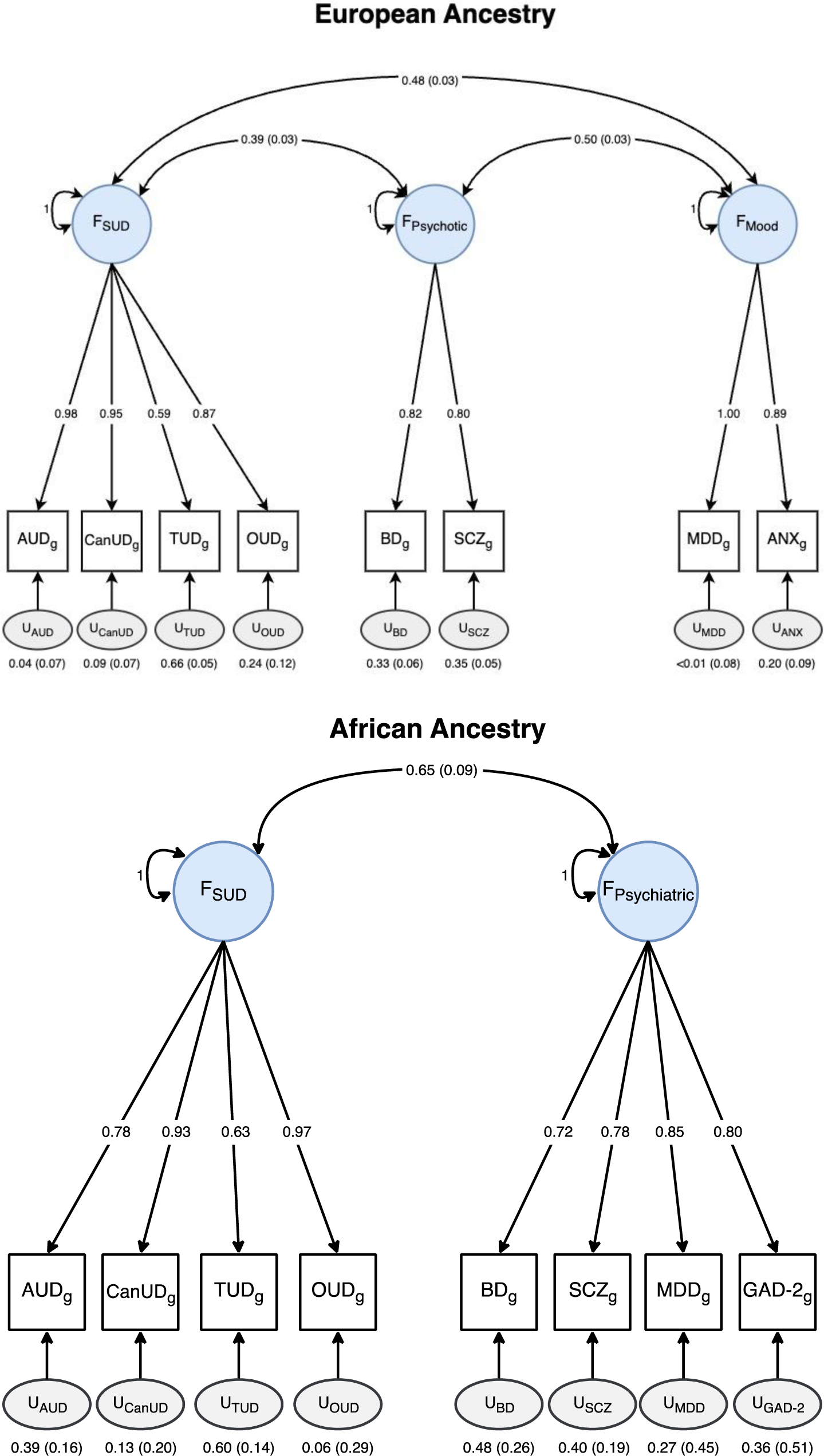
Genomic structural equation models. AUD = alcohol use disorder, CanUD = cannabis use disorder, TUD = tobacco use disorder, OUD = opioid use disorder, BD = bipolar disorder, SCZ = schizophrenia, MDD = major depressive disorder, ANX = anxiety, GAD-2 = 2-item GAD questionnaire.

The SUD factor GWAS identified 143 lead SNPs (Supplementary Table 15), of which 47 were not genome-wide significant (GWS) or in LD with any GWS SNPs in the input GWAS, although they had all been significantly associated with SUDs or SUD-related traits in prior GWAS (Supplementary Table 15).^57^ There were 17 independent Q-SNPs that demonstrated significant heterogeneity across SUDs (Supplementary Table 6). The psychotic disorder factor GWAS identified 9 independent Q-SNPs (Supplementary Table 7) and 162 lead SNPs (Supplementary Table 16), 27 of which had not been identified by the SCZ or BD GWAS, but all of which had previously been GWS or in LD with GWS SNPs in at least one psychotic trait GWAS (Supplementary Table 16). The mood factor GWAS identified 14 independent Q-SNPs (Supplementary Table 8) and 112 lead SNPs (Supplementary Table 17), 13 of which were not identified by the MDD or ANX GWAS, and 2 of which (rs75174029 and rs7652704) were not previously significantly associated with or in LD with GWS SNPs for any mood or anxiety disorder (Supplementary Table 17). PheWAS in the GWAS Atlas identified associations of rs75174029 with the number of non-cancer related illnesses, general risk tolerance, and having recent trouble relaxing; and rs7652704 with sensitivity/hurt feelings, neuroticism, and positive affect, among other related traits (Supplementary Figure 4).

rs75174029 is an intronic variant in *FOXP1*, which serves as a key regulatory gene in neural development.^58,59^ Finemapping identified the SNP as the most likely causal variant accounting for the observed association within the locus (PICS probability = 0.307; Supplementary Table 18). Additionally, Hi-C chromatin interaction data (Supplementary Figure 5) revealed that rs75174029 contacts several regions of *FOXP1*, including its promoter. Taken together, this evidence suggests rs75174029’s association with mood/anxiety disorders may be due to its presence in an enhancer with cis-regulatory function on *FOXP1* during neural development. The second novel SNP, rs7652704 (Supplementary Figure 6), is an intronic variant in an uncharacterized non-coding RNA locus. rs7652704 and 20 other SNPs in strong LD all had PICS probabilities of 0.027, suggesting difficulty in determining the most likely causal variant accounting for the association in the locus (Supplementary Table 19). rs7652704 is an eQTL of *NECTIN3* (also known as *PVLR3)* in cultured fibroblasts and displays chromatin interaction with both *BTLA*, which is a gene involved in immune response,^60^ and *NECTIN3*, which encodes a nectin adhesion molecule that regulates cell organization and modulates stress responses.^61–63^ Thus, ample evidence associates both rs75174029 and rs7652704 with regulatory roles.

Gene-tissue expression analyses showed a role for both SUD- and psychotic disorder-related genes during prenatal brain development (Supplementary Figures 7 and 8), but no developmental period was significant for the mood disorders factor (Supplementary Figure 9). Mapped genes from each of the factor GWAS were significantly enriched for protein-protein interactions, indicating shared biological functions among disorders of the same class (SUDs = 1.37x, psychotic = 1.40x, and mood/anxiety disorders = 1.43x enrichment; *p*s < 1.00 x 10^-16^).

**Figure 3.**
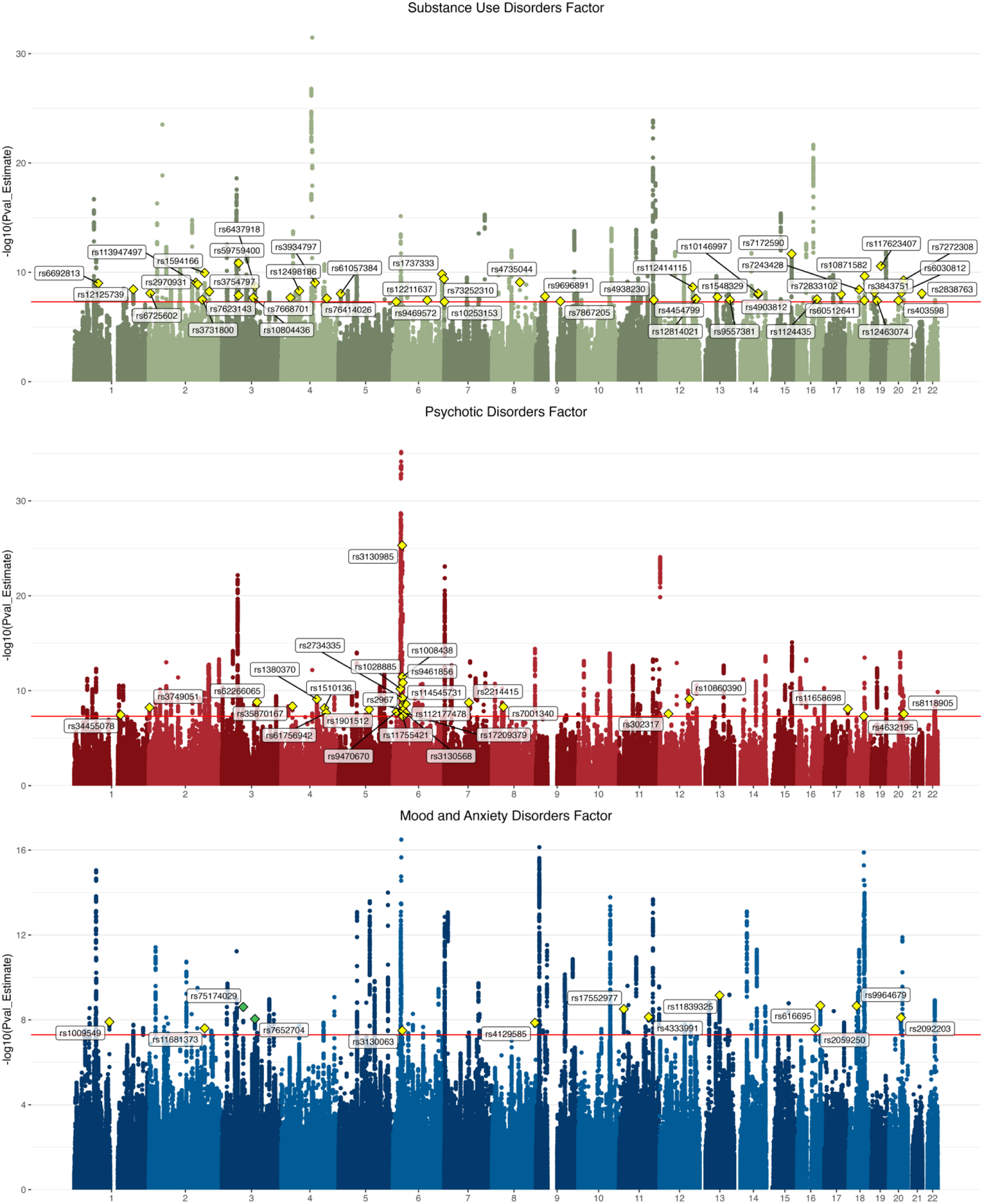
Manhattan plots for common factors. The substance use disorders factor GWAS identified 143 lead SNPs, the psychotic disorders factor identified 162 lead SNPs, and the mood/anxiety disorders factor identified 112 lead SNPs. The lead SNPs for loci that were not significant in the input GWAS are annotated with yellow diamonds, and lead SNPs for loci not previously significantly associated with phenotypes related to the common factor (i.e., novel) are annotated with green diamonds.

#### African Ancestry

Fit was generally poor for each of the EFA models (all CFIs < 0.40; Supplementary Table 20). Upon examining each model’s factor loadings, the proportion of variance explained by each factor, and eigenvalues, we tested two CFA models: a 2-factor model representing SUDs and psychiatric disorders and a 3-factor CFA model replicating the factor structure identified in EUR individuals. Attempting to fit the 3-factor CFA led to a negative residual variance and a correlation >1 between the psychotic and mood disorder factors. Because negative variances and correlations that are out of bounds can indicate issues with model misspecification or overfitting the data, we proceeded with the 2-factor CFA model, which had an adequate fit (𝜒^2^(19) = 21.49, *p* = 0.31, AIC = 55.49, CFI = 0.99, SRMR = 0.10) and required no constraints (Figure 2 and Supplementary Table 21).

The SUD factor GWAS (Supplementary Figure 10) identified 1 lead SNP, rs1944683, positioned within an intergenic region on chromosome 11. To our knowledge, this variant has not previously been GWS or in LD with any GWS SNPs in AFR GWAS of substance use traits. However, the locus has been associated with alcohol consumption, tobacco-related traits, opioid use disorder, and cannabis use disorder in EUR and cross-ancestry studies.^64–66^ The SNP exhibited chromatin interaction with two genes: *BLID* and *C11orf63*. A PheWAS of the lead SNP in the GWAS Atlas identified significant associations with regular smoking, past-month stomach pain, and tobacco-related conditions, such as atrial fibrillation and respiratory function (i.e., forced vital capacity and peak expiratory flow). One lead Q_SNP_, rs10489130, identified on chromosome 4, exhibited significant associations with AUD only and was GWS in a previous AUD GWAS in AFR individuals, suggesting that this SNP may display specificity for AUD rather than influencing SUDs broadly.^66^

For the SUD factor, gene expression was significantly enriched in brain tissues involved in emotion processing, reward signaling, and cognitive control, including the putamen, amygdala, caudate, and hippocampus (Supplementary Figure 11). No significant variants were identified for the psychiatric disorders factor (Supplementary Figure 12). Gene expression was also not significantly enriched for any developmental stage or tissue type, although the top associations were with brain tissues (Supplementary Figure 13).

### Second-Order Common Factors

#### European Ancestry

The SUD factor was significantly genetically correlated with both the psychotic (r_g_ = 0.38, SE = 0.03, *p* < 0.001) and mood (r_g_ = 0.44, SE = 0.03, *p* < 0.001) disorder factors. We used a higher-order CFA model (𝜒^2^(2) = 57.61, *p* = 3.09x10^-13^, AIC = 65.61, CFI = 0.91, SRMR = 0.07; Supplementary Figure 14 and Supplementary Table 22) to examine this shared genetic structure. After accounting for shared genetic risk, there was less standardized residual variance in the SUD factor (u_SUD_ = 0.19, SE = 0.04) than the psychotic (u_Psychotic_ = 0.63, SE = 0.05) or mood disorder factors (u_Mood_ = 0.56, SE = 0.04). A GWAS of the SUD and psychotic disorders factor identified 4 independent Q-SNPs (Supplementary Table 9) and 76 lead SNPs (Supplementary Table 23 and Supplementary Figure 15), 12 of which were not significant in any input or the first-order GWAS. For the SUD and mood factor, 6 independent Q-SNPs (Supplementary Table 10) and 62 lead SNPs were identified (Supplementary Table 24 and Supplementary Figure 16), 5 of which were in loci that were not significant in any input or first-order GWAS. All lead SNPs at the second-order level were previously associated with related traits.

Genetic risk shared among SUD and psychotic disorders implicated several gene sets, including molecular functions such as transcription regulation and sequence-specific DNA binding, and biological processes such as neuron differentiation. Genetic risk shared between SUDs and mood disorders was associated with enrichment in two gene sets involved in the biological processes of mechanosensory behavior and axonal protein transport. For both second-order factors, gene expression was enriched in brain tissues (Supplementary Figures 17 and 18). Genes associated with both second-order factors were significantly enriched for protein-protein interactions (SUDs and psychotic disorder = 1.54x, SUDs and mood/anxiety disorders = 1.57x; *p*s < 1.00 x 10^-16^).

#### African Ancestry

The SUD and psychiatric disorder factors were highly genetically correlated (r_g_ = 0.74, SE = 0.13, *p* < 0.001), and a second-order CFA model accounting for this shared genetic risk fit the data well (𝜒^2^(19) = 21.49, *p* = 0.31, AIC = 55.49, CFI = 0.99, SRMR = 0.10; Supplementary Table 25 and Supplementary Figure 19). Although no significant SNPs were identified by the second-order GWAS (Supplementary Figure 20), there was enriched gene expression in several brain regions, including those associated with reward processing (putamen, caudate, nucleus accumbens), emotion processing and memory (amygdala, hippocampus), and executive functions and decision-making (anterior cingulate cortex; Supplementary Figure 21).

### GWAS-by-Subtraction

Common and independent genetic effects were examined for TUD, SCZ, and BD, which each had a residual variance ≥0.30 in the EUR first-order common factor models. We chose this threshold because analyses performed on traits with less residual variance were underpowered for parsing transdiagnostic from disorder-level effects. We identified 102 GWS lead SNPs for TUD Common, which represented genetic effects on TUD that operate through the SUD factor. There were 20 lead SNPs for TUD Independent, which represented genetic effects on TUD that are not shared with the other three SUDs. 13 of these SNPs did not reach significance in the TUD Common GWAS (Supplementary Table 26). Chromatin interaction mapping identified several of the nicotinic acetylcholine receptor genes (*CHRNA2, CHRNA4,* and *CHRNA5*; Supplementary Figure 22). Gene-set analyses for TUD Independent implicated genes involved in nicotinic acetylcholine reception and behavioral responses to nicotine.

We identified 51 lead SNPs for SCZ Common and 18 for SCZ Independent, of which 15 were not GWS nor in LD with GWS SNPs in the SCZ Common GWAS (Supplementary Table 27). Chromatin interaction mapping identified several genes on chromosome 6, including the *ZSCAN* and *HIST1H* gene families, as possible sources of functional effects related to SCZ risk independent of BD (Supplementary Figure 23). Although after Bonferroni correction no gene-sets were significantly enriched for SCZ Independent, the top set involved up-regulated genes in the prefrontal cortex in mouse models of 22q11.2 microdeletions, which, in humans, are associated with risk of developing SCZ.^67^ Finally, there were 189 significant lead SNPs for BD Common and 13 for BD Independent, 10 of which did not reach significance in the BD Common GWAS (Supplementary Table 28). Although few SNPs were significant in the BD Independent GWAS, chromatin interactions identified potential functional effects of these variants on genes, including *MXI1*^68^ and *ADD3*,^69^ which have been previously associated with BD (Supplementary Figure 24). For BD Independent, one gene-set involved in the regulation of trans-synaptic signaling remained significant after Bonferroni correction. Protein-protein interactions were significantly enriched for TUD, SCZ, and BIP Independent (TUD = 3.31x, *p* = 3.45 x 10^-13^, SCZ = 5.48x, *p* < 1.00 x 10^-16^, BIP = 5.75x, *p* = 2.19 x 10^-11^), indicating potential molecular mechanisms with enhanced specificity (Supplementary Figures 25-27).

**Figure 4.**
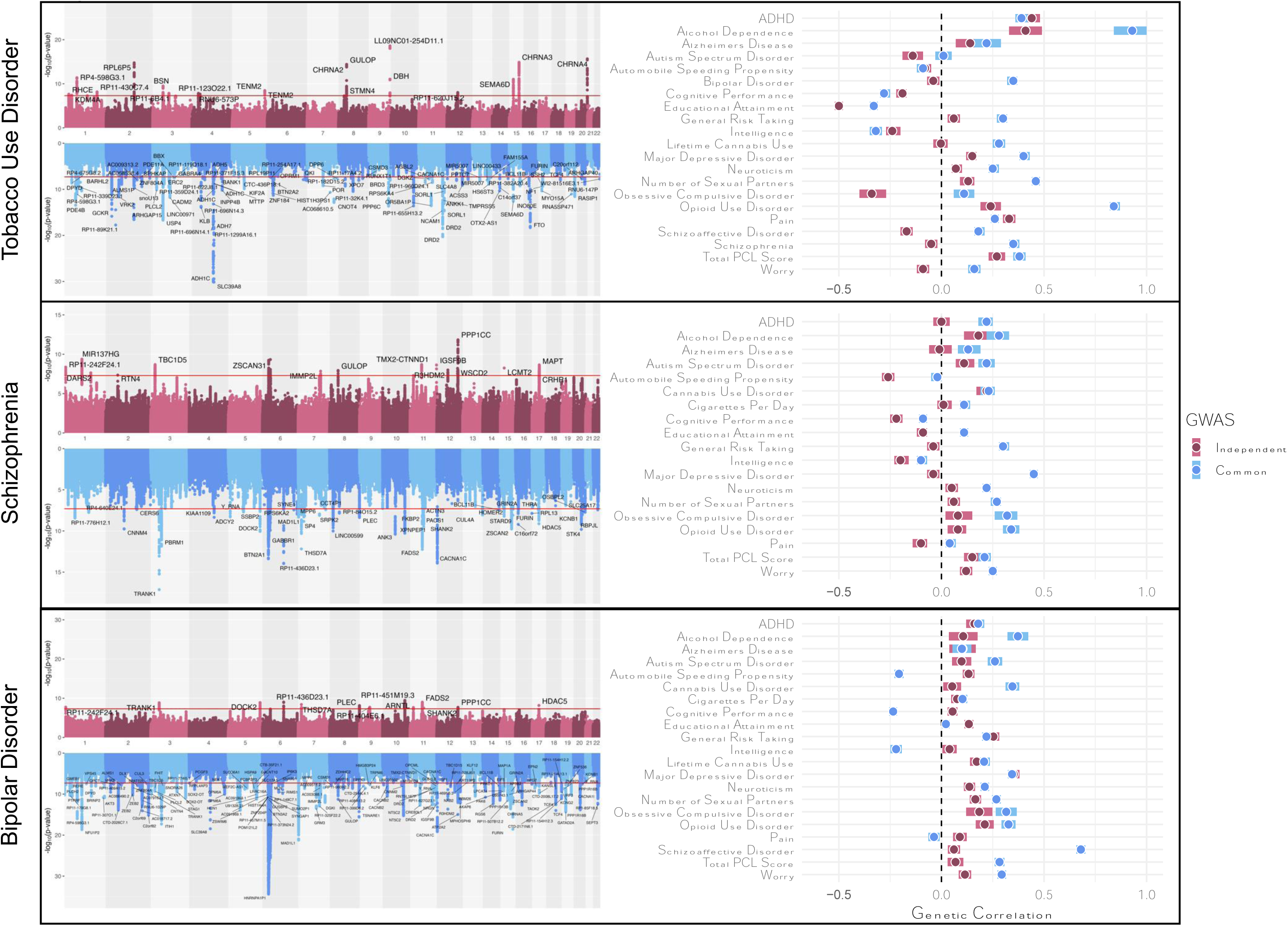
Hudson plots and genetic correlations of GWAS-by-subtraction models. The left panel presents Hudson plots of the GWAS-by-subtraction model results with the mapped gene for lead SNPs annotated. The right panel presents the genetic correlation results. Independent GWAS refers to the influences on a disorder that do not operate through the common factor, while the Common GWAS refers to influences on the disorder that do operate through the common factor.

### Genetic Correlations with gSEM Factors

#### European Ancestry

The SUD factor was, as expected, strongly genetically correlated with smoking and alcohol traits, as well as depression and socioeconomic factors, including reduced educational attainment, unemployment due to sickness/disability, and the Townsend deprivation index (Supplementary Figure 28). The psychotic factor correlated most strongly with related traits, such as SCZ, BD, depression, and anxiety. However, psychotic disorders also exhibited a positive genetic correlation with risk taking and a negative correlation with cognitive performance, which consists of fluid intelligence and the first principal component of scores on neuropsychological tests (Supplementary Figure 29). Although the mood disorders factor correlated most strongly with measures of depression and anxiety, the remaining correlations were predominantly with somatic conditions, such as chronic pain, longstanding illness/disability/infirmity, and prescription medication usage (Supplementary Figure 30).

For the second-order factors, the top genetic correlations were a mixture of SUD-related, psychiatric, and medical traits. The SUD and psychotic disorders factor, for example, correlated most strongly with SCZ and BD. Other significant genetic correlations included smoking-related traits, cognitive measures, and risk taking, as with the first-order psychotic disorder factor. The SUD and mood disorders factor correlated most strongly with mood and anxiety traits and illness and medication use for pain or gastrointestinal problems, reflecting similar associations observed for the first-order mood disorders factor (Figure 5).

Genetic correlations underscored differences between disorder-level and transdiagnostic genetic effects (Figure 4). Although TUD Common was significantly positively genetically correlated with SCZ (r_g_ = 0.35, SE = 0.02, *p* < 0.001), TUD Independent was not (r_g_ = -0.05, SE = 0.03, *p* = 0.08), with a similar pattern observed for other thought/psychotic disorders. SCZ Common had a nominally weaker negative genetic correlation with cognitive performance (r_g_ = - 0.09, SE = 0.02, *p* < 0.001) and was significantly positively correlated with educational attainment (r_g_ = 0.11, SE = 0.02, *p* < 0.001), while SCZ Independent had consistently negative associations with both (r_g_ = -0.22, SE = 0.03, *p* < 0.001 and r_g_ = -0.09, SE = 0.02, *p* < 0.001, respectively). Among other traits, BD Common and Independent showed opposite patterns of associations with automobile speeding propensity and cognitive performance, both of which were negative for BD Common (r_g_ = -0.21, SE = 0.02, *p* < 0.001; and r_g_ = -0.24, SE = 0.02, *p* < 0.001, respectively) and positive for BD Independent (r_g_ = 0.13, SE = 0.03, *p* < 0.001; and r_g_ = 0.05, SE = 0.02, *p* = 0.03, respectively). SCZ and BIP Independent had significantly different associations with risk-taking (r_g_ = 0.25, SE = 0.03, *p* < 0.001 vs. r_g_ = -0.04, SE = 0.03, *p* = 0.14; Z = 6.84, *p* = 8.18 x 10^-12^) and MDD (r_g_ = 0.35, SE = 0.03, *p* < 0.001 vs. r_g_ = -0.04, SE = 0.03, *p* = 0.22; Z = 9.19, *p* = 3.84 x 10^-20^).

#### African Ancestry

The psychiatric disorders factor was genetically correlated with all 11 traits examined, and the SUD factor was significantly correlated with all except PTSD. There were minimal differences between the genetic correlations for the first-order factors, likely due to the large variance in estimates resulting from low statistical power (Supplementary Figure 31). The second-order SUD and psychiatric factor correlated significantly with all traits except PTSD, and the strongest correlations were with smoking trajectory, OUD, depression, and maximum alcohol consumption (Supplementary Figure 32).

#### Trans-ancestry

The EUR SUD factor was significantly genetically correlated with the AFR SUD factor (r_g_ = 0.730, SE = 0.094, *p* = 0.004). Similarly, the AFR psychiatric disorders factor was genetically correlated with both the EUR psychotic factor (r_g_ = 0.471, SE = 0.216, *p* = 0.014) and the EUR mood disorders factor (r_g_ = 0.571, SE = 0.204, *p* = 0.035).

**Figure 5.**
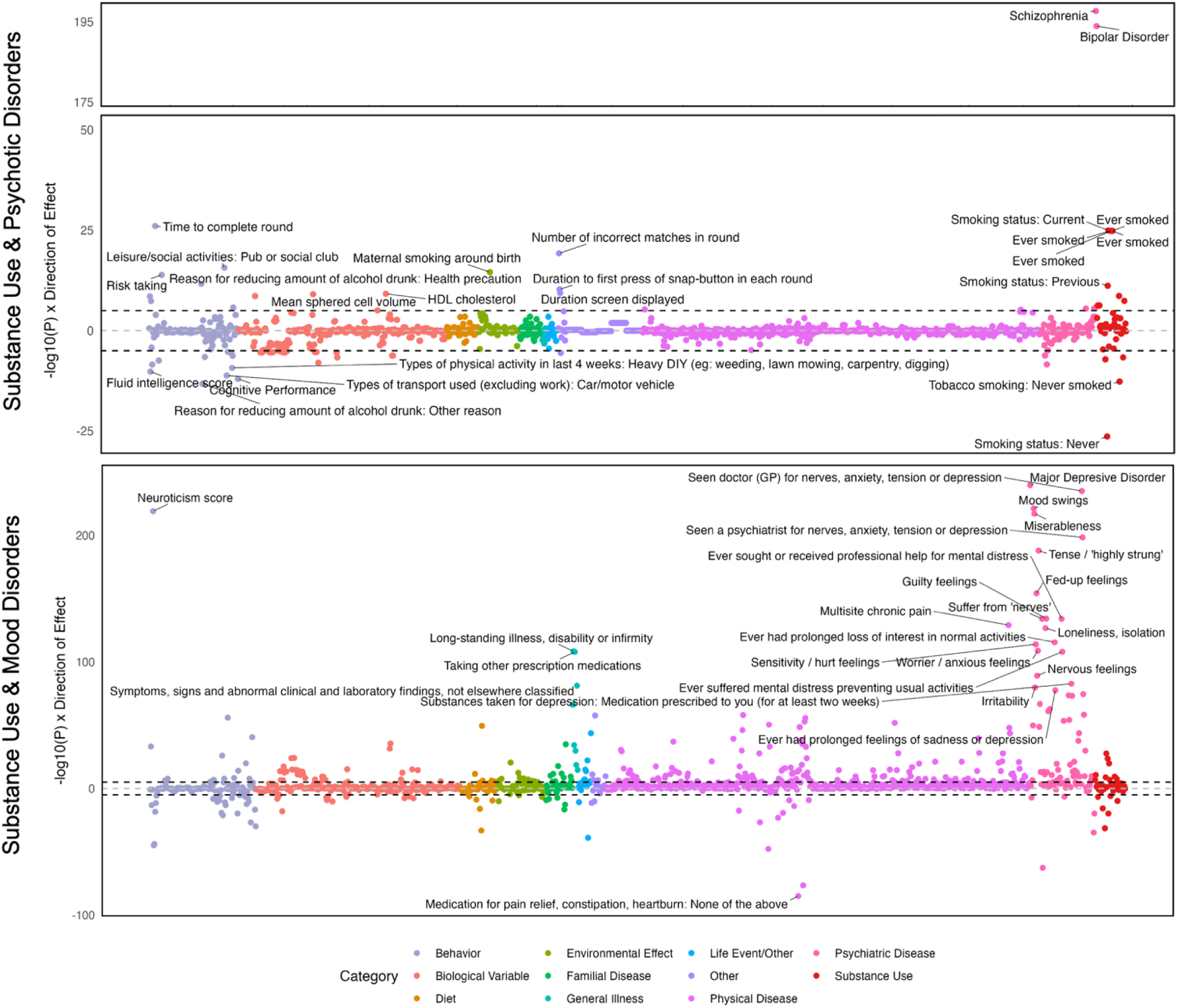
Genetic correlations for second-order common factors using Complex Trait Genetics Virtual Lab. The dashed line represents the log-transformed Bonferroni-corrected p-value across the 1,437 traits included in the analysis.

### PheWAS in Penn Medicine BioBank

#### European Ancestry

Among participants in PMBB, the SUD factor was associated with TUD, tobacco-related illnesses (lung and other respiratory system cancers and chronic airway obstruction), and mood/anxiety disorders (Supplementary Figure 33 and Supplementary Table 29). The psychotic disorders factor was significantly associated only with psychiatric traits, including BD and mood/anxiety disorders (Supplementary Figure 34 and Supplementary Table 30). In contrast, although the mood disorders factor was most strongly associated with the presence of various mood and anxiety disorders, it was also associated with physical health conditions like pain, sleep disorders, and obesity (Supplementary Figure 35 and Supplementary Table 31).

The second-order SUD and psychotic disorders factor was significantly associated with tobacco and alcohol-related disorders, with a nonsignificant association with BD (Supplementary Figure 36 and Supplementary Table 32). Although the PheWAS of the SUD and mood disorders factor revealed the strongest associations with substance use and psychiatric disorders, it also showed significant associations with pain, type 2 diabetes, ischemic heart disease, hypertension, and sleep disorders, amongst other physical health conditions (Supplementary Figure 37 and Supplementary Table 33).

PheWAS further highlighted the enhanced specificity of the GWAS-by-subtraction models. Although the TUD Common factor was associated with multiple SUDs, the TUD Independent factor demonstrated greater specificity, having the highest associations with TUD and related medical conditions, such as chronic airway obstruction (Supplementary Figure 38). PheWAS for SCZ Common factor showed broad associations with mood disorders, including BD, but there were no significant associations for SCZ Independent (Supplementary Figure 39). PheWAS of both the BD Common and Independent PRS showed significant associations with BD and other mood disorders, but the association with depression was only significant for BD Independent (Supplementary Figure 40).

#### African Ancestry

Although there were no statistically significant phenotypic associations among AFR individuals in PMBB, the top hits generally aligned with the factor being examined. For example, the SUD factor was most strongly associated with TUD, followed by SUDs broadly (Supplementary Figure 41 and Supplementary Table 34). Among the top associations for the psychiatric factor were generalized anxiety disorder and the “other mental disorder” phenotype (Supplementary Figure 42 and Supplementary Table 35). Similar results were seen for the second-order SUD and psychiatric factor, which included TUD, alcohol-related disorders, and mood disorders within the top associations (Supplementary Figure 43 and Supplementary Table 36).

## Discussion

Leveraging the largest available GWAS in European- and African-ancestry individuals, we combined complementary multivariate methodologies to examine the shared genetic architecture across SUDs, psychotic, mood, and anxiety disorders. We also examined genetic effects that operate independent of the shared genetic risk to influence disorders. By integrating transdiagnostic and disorder-level gSEM, we identified potential biological mechanisms that contribute to comorbidity across disorder classes and those that distinguish commonly co-occurring conditions. Our findings revealed both pervasive pleiotropy across SUDs and other psychiatric disorders *and* trait-specific associations, while also highlighting the need to increase representation of non-European ancestry individuals in genetic studies of mental health.

### Identification of SUDs, Psychotic, and Mood Disorder Factors

Consistent with other psychiatric genetic findings,^19,70^ we identified common genetic factors underlying disorders that exhibit shared features. As expected, MDD and anxiety, which are highly comorbid and share similar symptoms, loaded onto the same factor. Additionally, across ancestries, common genetic risk partially accounted for the shared features of BD and SCZ. Other gSEM studies have also showed that SCZ and BD load onto the same genetic factor,^18,70–72^ highlighting their strong shared etiology despite belonging to different diagnostic classes. Although smaller samples and greater genetic diversity limited statistical power to replicate the EUR factor structure in AFR individuals, there were significant commonalities across genetic ancestry groups. For example, in both AFR and EUR models, SUDs loaded onto a single factor that was highly genetically correlated with a previously identified genetic addiction factor.^19^ Additionally, trans-ancestry genetic correlations highlighted the consistency of the genetic influences on the AFR psychiatric disorders factor with both the EUR psychotic and mood disorders factors.

In AFR individuals, we identified a lead SNP (rs1944683) associated with SUDs that had not been previously GWS or in LD with GWS SNPs in AFR substance-related GWAS. This locus has, however, been previously implicated in alcohol, tobacco, cannabis, and opioid-related traits in EUR and cross-ancestry GWAS.^64–66^ Our ability to identify this lead SNP was facilitated by the use of gSEM, which allowed us to leverage power across multiple SUDs. In doing so, we were able to overcome some of the limitations posed by the relatively low statistical power of existing GWAS in AFR populations. However, despite statistical advancements, our study remained underpowered in AFR individuals.

In EUR individuals, we detected two significant loci for the mood and anxiety disorders factor that had not been previously significant in GWAS of mood or anxiety disorders. Performing chromatin interaction mapping on the lead SNPs for these loci implicated genes involved in immune and stress responses (*BTLA* and *NECTIN3*, respectively) and a gene related to hippocampal development (*FOXP1*).^58,60,62,73^ The identification of *BTLA*, which encodes the B- and T-lymphocyte attenuator protein, is consistent with other research highlighting the role of immune dysregulation in psychiatric pathogenesis.^74–76^ *NECTIN3* encodes a cell adhesion molecule that is involved in synaptic plasticity, enriched in hippocampal neurons, and implicated in stress-related disorders.^61,62,77^ FOXP1, a transcription factor that plays a crucial role in hippocampal development, has also been implicated in the regulation of synaptic plasticity.^58^ Further highlighting the potential role of immune functioning, these loci have previously been implicated in GWAS of lymphocyte and leukocyte counts, as well as autoimmune conditions, such as inflammatory bowel disease. Thus, dysfunctions in synaptic plasticity and immune regulation may underlie a range of psychiatric conditions, including MDD and anxiety.^78,79^ Targeting these common biological pathways could lead to the development of therapeutics with broader efficacy profiles.

In EUR individuals, both the SUD and psychotic disorder factors showed a significant association with genes expressed in brain tissue during prenatal development. Although no developmental stage reached significance for mood disorders or in AFR individuals, the top associations were consistently prenatal periods. This underscores the importance of early neurodevelopmental processes, including neurogenesis, synaptogenesis, and the formation of neurotransmitter systems, in shaping the susceptibility to SUDs and psychiatric disorders. Furthermore, prenatal development may represent a sensitive window during which genetic and environmental factors interact to influence long-term mental health outcomes.^80^ Epigenetic mechanisms, such as those modulated by maternal stress and other environmental conditions,^81,82^ may play a crucial role in mediating gene expression patterns and contributing to the developmental origins of psychiatric disorders. The specifics of these epigenetic mechanisms and other non-coding regulatory processes remain largely unclear. However, statistical and technological advances, such as next-generation sequencing and multi-omics analysis, show promise for furthering knowledge in this area.^83^

### SUDs Share Genetic Liability with Psychotic and Mood Disorders

Evaluating shared genetic variance across the common factors revealed higher-order dimensions of liability to psychopathology, including genetic risk shared between SUDs and psychotic disorders and between SUDs and mood/anxiety disorders. A PheWAS in the PMBB showed broad phenotypic manifestations of these dimensions of genetic liability. Notably, in EUR individuals, SUD and mood/anxiety disorders exhibited significant associations with various physical health conditions, including obesity, type 2 diabetes, chronic pain, chronic airway obstruction, heart disease, hypertension, and sleep disorders. Because the PheWAS was underpowered in AFR individuals, there were no significant associations, though the top associations included relevant phenotypes, such as alcohol-related disorders, TUD, and mood disorders.

Genetic correlations further underscored the pervasive impact of genetic risk for psychiatric comorbidities and SUDs. The second-order factors (i.e., SUD and mood disorders and SUD and psychotic disorders) were genetically correlated with adverse outcomes that included lower cognitive performance, elevated HDL cholesterol, chronic pain, long-standing illness, and miserableness. In AFR individuals, the second-order factor, representing shared genetic effects across SUDs and psychiatric disorders, was significantly genetically correlated with pain intensity and various substance use and psychiatric phenotypes. Thus, genetic risk for co-occurring SUDs and psychiatric disorders has far-reaching implications for both mental and physical health.

In addition to shared variance, in EUR individuals there was substantial unique genetic variance for both psychotic (0.63, SE = 0.05) and mood/anxiety disorders (0.56, SE = 0.04). The residual genetic variance in SUDs was also significant, albeit smaller, after accounting for genetic variance shared with psychotic and mood/anxiety disorders (0.19, SE = 0.04). In EUR individuals, more of the genetic variance in SUDs was shared with mood/anxiety than psychotic disorders. This suggests that there are different degrees of genetic convergence between SUDs and their common psychiatric comorbidities. Evaluating these patterns of genetic overlap and heterogeneity can help distinguish highly comorbid psychiatric disorders and pinpoint both shared and distinct etiological mechanisms.

### Specificity of Genetic Effects for Tobacco Use Disorder, Schizophrenia, and Bipolar Disorder

To evaluate transdiagnostic and disorder-level liability, SNPs for TUD, SCZ, and BD were parsed into effects that: (1) operated through their respective common factors or (2) were uncorrelated with the common factor. Thus, we were able to distinguish biological mechanisms and genetic correlates that contribute to comorbidity across disorders from those that show greater specificity. This hierarchical approach can yield genetic knowledge across levels of psychopathology and identify patterns of convergence and divergence across disorders.

As an example of the utility of this approach, GWAS-by-subtraction findings provided insights into the underlying structure of psychotic disorders. SCZ and BD shared a common genetic core, consistent with their shared psychotic features and in line with empirical nosological models.^84^ However, the genetic risk for each disorder that was independent of this common psychotic core showed different patterns of associations with other complex traits. For example, SCZ Independent was more negatively genetically correlated with measures of cognition and educational attainment than BD Independent, which was more strongly associated with risk-taking behaviors and affective disorders. Our results are highly consistent with the expanded psychosis continuum hypothesis, which proposes that although SCZ and BD share a psychotic core, cognitive and affective domains differentiate the disorders.^85^ Specifically, SCZ is characterized by greater cognitive impairments and BD by greater affective impairments. Our findings lend genetic support to this hypothesis, previously investigated using only psychological and neural evidence.^85^

PheWAS results further showcased the enhanced specificity of findings when independent PGS for TUD, SCZ, and BD were compared with transdiagnostic liability. For example, TUD Independent exhibited several associations not observed for the TUD Common factor, including ischemic heart disease, atherosclerosis, obesity, and skin conditions. This may reflect a combination of unhealthy lifestyle factors, direct physiological effects of tobacco use, and shared biological pathways underlying these conditions. This finding is also consistent with epidemiologic and genetic research supporting tobacco use as one of the strongest risk factors for cardiovascular disease.^86–88^ Differential patterns also emerged for SCZ and BD, with SCZ Common and BD Independent showing broader associations with mood disorders than their respective components. Recent research comparing transdiagnostic and disorder-specific genetic effects across 11 psychiatric disorders similarly observed substantial differences in genetic associations,^89^ as has research parsing alcohol-specific risk from broader externalizing liability.^90^ Thus, hierarchical genetic approaches can facilitate a more nuanced understanding of patterns of comorbidity and heterogeneity across co-occurring conditions.

These findings also pave the way for developing more refined PGS than are currently available. For example, depression PGS show little specificity, accounting for a similar amount of variance in mood disorders, anxiety disorders, ADHD, and SUDs.^91^ Similarly, an evaluation of 16 PGS for psychiatric phenotypes found that most were associated with general psychopathology rather than the specific domain for which they purported to measure risk.^92^ Although PGS have shown clinical utility in predicting some non-psychiatric disorders,^93,94^ prediction performance for psychiatric phenotypes remains limited.^95^ This may be due in part to a lack of specificity of currently available psychiatric PGS. Existing PGS are useful for providing a broad overview of genetic predispositions to psychiatric disorders. However, when the focus shifts to exploring associations of a given psychiatric disorder without the confounding influence of co-occurring conditions, a more granular PGS is needed. Efforts to develop more precise PGS, including via the combined application of transdiagnostic and disorder-level genetic methods, may ultimately enhance their clinical utility.

## Conclusions

Using a combination of multivariate approaches across multiple ancestral groups, we examined the common and independent genetic effects on SUDs, psychotic, mood, and anxiety disorders. In doing so, we identified potential biological mechanisms contributing to comorbidity both within and across classes of disorders, while also identifying pathways that may distinguish commonly co-occurring conditions. Isolating transdiagnostic genetic risk factors from those exhibiting greater specificity may aid in enhancing the precision of genetic prediction, as we observed when comparing genetic correlations and phenotypic associations of genetic risk factors. Thus, integrating transdiagnostic and disorder-level genetic models both clarifies the biological underpinnings of psychiatric comorbidity and identifies distinct biological pathways that contribute to heterogeneity within classes of psychiatric conditions.

In addition to advancing our understanding of the genetic architecture of SUDs and other psychiatric disorders, our study also highlights the importance of inclusivity in genetic research. Previous gSEM studies have been limited to European-ancestry individuals,^16,96,97^ but as gSEM has shown the potential to uncover novel genetic associations and provide greater insights into the etiology of complex traits and diseases, it is imperative that these advances be made available to individuals of all ancestral backgrounds. Unfortunately, in combination with smaller samples, lower LD and higher genetic diversity among African-ancestry individuals present statistical challenges for many existing analyses.^98^ This is further complicated by the high degree of admixture present among individuals of non-European ancestries within the United States.^99^ Accurate consideration of population substructure via the use of large, representative reference panels is essential for advancing genetic discovery. We provide details (Supplementary Materials) on the efforts we took to ensure the inclusion of African-ancestry individuals despite present limitations in the hopes that this may aid researchers faced with similar decisions. Advancing genetic discovery across diverse populations will be key in shaping our understanding of psychiatric etiology.

## Data Availability

GWAS Summary Statistics used for the present study can be accessed at the following locations: UK Biobank (http://www.nealelab.is/uk-biobank/), dbGaP accession phs001672 for Million Veteran Program (https://www.ncbi.nlm.nih.gov/projects/gap/cgi-bin/study.cgi?study_id=phs001672), iPSYCH (https://ipsych.dk/en/research/downloads/), Psychiatric Genetics Consortium (https://pgc.unc.edu/for-researchers/download-results/), the Social Science Genetic Association Consortium (https://thessgac.com/), GWAS catalog (https://www.ebi.ac.uk/gwas/publications/28979981; https://www.ebi.ac.uk/gwas/publications/33532862), Gelernter Lab website (https://medicine.yale.edu/lab/gelernter/stats/), Global Biobank Meta-analysis Initiative (https://www.globalbiobankmeta.org/resources), and the Diabetes Genetics Replication and Meta-Analysis Consortium (https://diagram-consortium.org/downloads.html). Once accepted for publication, summary statistics for the first-order, second-order, and GWAS-by-subtraction analyses will be made publicly available.

## Acknowledgements

This study is supported by NIH grants K01AA028292 (R.L.K.), R01MH125938 (R.E.P.), P50AA022537 (R.E.P.), T32 HG009495 (K.L.F.), the Brain & Behavior Research Foundation NARSAD grant 28632 PS Fund (R.E.P.), the California Tobacco-Related Disease Research Program (TRDRP; T32IR5226) (S.S-R), the Department of VA Office of Research and Development grants IK2 CX002336 (E.E.H.), I01 BX003341 (H.R.K.) and the Veterans Integrated Service Network 4 Mental Illness Research, Education and Clinical Center. This publication does not represent the views of the Department of VA or the United States Government. We acknowledge the Penn Medicine BioBank (PMBB) for providing data and thank the patient-participants of Penn Medicine who consented to participate in this research program. We would also like to thank the Penn Medicine BioBank team and Regeneron Genetics Center for providing genetic variant data for analysis. The PMBB is approved under IRB protocol #813913 and supported by the Perelman School of Medicine at the University of Pennsylvania, a gift from the Smilow family, and the National Center for Advancing Translational Sciences of the National Institutes of Health under CTSA award number UL1TR001878.

