## Supplementary Materials for "Combining Transdiagnostic and Disorder-Level GWAS Enhances Precision of Psychiatric Genetic Risk Profiles in a Multi-Ancestry Sample"

#### MTAG

Using Multi-Trait Analysis of GWAS (MTAG),[1] we leveraged the genetic effects from a study of Lifetime Anxiety Disorder[2] and a study of GAD-2 questionnaire scores[3] to enhance the statistical power of a GWAS for a broad spectrum of anxiety disorders[4] in European ancestry individuals. We chose to enhance the power of the summary statistics from Otowa, et al., 2016, because they included the most diverse array of anxiety disorders among the three anxiety GWAS. This choice was supported by the strong genetic correlations between the generalized anxiety GWAS[4] and both GWAS of lifetime anxiety disorder ($r_g$ = 0.7429) and GAD-2 scores ($r_g$ = 0.7309)

Effective sample sizes were calculated as the sum of 4/(1/*n*_case + 1/*n*_control) for each cohort in each of the two case-control GWAS. For the GAD-2 score GWAS, the total sample size was used as the input for MTAG because GAD-2 is a continuous trait. As quality control measures included in the MTAG software, SNPs with MAF<0.01 were excluded from analysis, along with duplicate SNPs and those with missing values. Following MTAG analysis, the effective sample size for follow-up analyses was calculated using the formula described by Turley, et al., 2018.[1]

#### Procedures for Summary Statistics in GenomicSEM

All summary statistics and analyses were conducted on the NCBI hg19/GRCh37 genome assembly. For traits with continuous outcomes (i.e., GAD-2 score), the total sample size was used for LDSC and GenomicSEM computations. For traits with a binary outcome (i.e., case-control), the effective sample size column contained within the GWAS summary statistics was used. When no effective sample size column was present, effective sample size was calculated for each set of summary statistics using the formula described by Grotzinger, et al, 2023:[5]

$$N_{eff} = \sum_{k=1}^{N} 4 * v_k * (1 - v_k) * n_k$$

Where v and n are the sample prevalence and sample total, respectively, for the $k^{th}$ cohort of a GWAS meta-analysis of N cohorts. Summary statistics were then prepared for GWAS using the following options in GenomicSEM: The “se.logit” flag was set to “TRUE” when the standard error column reflected the standard error of a logistic beta, the “OLS” flag was set to “TRUE” when the phenotype reflected a continuous outcome, and the “linprob” flag was set to “TRUE” when the phenotype was of a binary outcome but with only Z-statistics present as a measure of effect in the GWAS summary statistics. SNPs were then filtered based on MAF>0.01 and 0.6. Following preparation of summary statistics, 2,083,079 SNPs remained for analysis in the European-ancestry subset, and 6,350,709 SNPs remained for analysis in the African-ancestry subset.

**African Ancestry Reference Panels**

To determine the optimal linkage disequilibrium (LD) score reference panel for use in the African ancestry gSEM models, we compared three sets of references: (1) 1000 Genomes Phase 3, (2) PanUKB, and (3) Million Veteran Program (MVP). We used publicly available 1000 Genomes[11] and PanUKB[12] LD scores. MVP LD scores were generated from 1000 randomly selected African ancestry MVP participants using covariate-adjusted LD score regression (cov-LDSC),[13] which is a method that has shown improved performance among admixed populations, such as African Americans. As recommended to further account for population stratification, the first ten ancestry-specific principal components (PCs) were computed within the sample and included as covariates when generating LD scores.

To ensure the accuracy of and prevent bias in estimates derived from the LD scores, we restricted LD score regression (LDSC) analyses to well-imputed, biallelic autosomal SNPs that are outside of the MHC region. The set of SNPs meeting this criteria varied for each LD reference panel. For the 1000 Genomes Phase 3 panel, we used the list of 1,217,312 HapMap3 SNPs provided in the reference files prepared by Finucane et al. (2015)[14] for LDSC. For the PanUKB reference, we retained all 1,190,983 SNPs, as LD scores were computed only for SNPs that met the aforementioned criteria and passed additional quality control, including having imputation quality ($R^2$) > 0.90 and minor allele frequency > 0.01 (see https://pan-dev.ukbb.broadinstitute.org/docs/ld/index.html). For the MVP reference, we restricted our analyses to SNPs that met the same criteria as those used by the Broad Institute to prepare the PanUKB reference files. Thus, a total of 2,388 SNPs were removed due to low MAF, and 8,707 were removed due to low imputation quality, leaving 1,516,281 SNPs in MVP.

In comparing the performance of the three sets of reference panels, we evaluated: (1) the number of SNPs retained following filtering and munging the input summary statistics, (2) liability scale SNP-based heritability, (3) confounding evidenced by inflated values on the LDSC intercept, and (4) the length and distribution of resulting LD blocks. Results are presented below:

| 1000G reference, 1000G SNPlist | | | | |
|---|---|---|---|---|
| **trait** | **# snps** | **heritability** | **SE** | **intercept** |
| AUD | 423441 | 0.0806 | 0.0133 | 1.0304 |
| TUD | 691706 | 0.0445 | 0.008 | 1.0257 |
| OUD | 250024 | 0.0668 | 0.0229 | 1.0236 |
| CanUD | 604363 | 0.0616 | 0.0116 | 1.0306 |
| GAD2 | 869992 | 0.0282 | 0.0365 | 1.0076 |
| MDD | 869312 | 0.0415 | 0.0188 | 1.0184 |
| SCZ | 891719 | 0.1204 | 0.0294 | 1.0587 |
| BIP | 891833 | 0.1417 | 0.0642 | 1.0344 |

| MVP reference, MVP SNPlist | | | | |
|---|---|---|---|---|
| **trait** | **# snps** | **heritability** | **SE** | **intercept** |
| AUD | 1508956 | 0.0427 | 0.0062 | 1.0615 |
| TUD | 1515026 | 0.0434 | 0.0065 | 1.0631 |
| OUD | 1512615 | 0.0218 | 0.0092 | 1.0347 |
| CanUD | 1507988 | 0.021 | 0.0056 | 1.0464 |
| GAD2 | 1476669 | 0.0391 | 0.0217 | 1.0056 |
| MDD | 1480787 | 1.00E-03 | 0.0082 | 1.0297 |
| SCZ | 1512163 | 0.0496 | 0.0155 | 1.0649 |
| BIP | 1511435 | 0.0554 | 0.0328 | 1.0324 |

| UKBB reference, Pan-UKBB SNPlist | | | | |
|---|---|---|---|---|
| **trait** | **# snps** | **heritability** | **SE** | **intercept** |
| AUD | 613531 | 0.0885 | 0.0153 | 1.0352 |
| TUD | 979504 | 0.064 | 0.0083 | 1.0235 |
| OUD | 429964 | 0.0662 | 0.0206 | 1.034 |
| CanUD | 897996 | 0.068 | 0.0116 | 1.0289 |
| GAD2 | 1152886 | 0.0619 | 0.0394 | 1.0026 |
| MDD | 1144306 | 0.0397 | 0.017 | 1.0193 |
| SCZ | 1184566 | 0.1661 | 0.0278 | 1.05 |
| BIP | 1183486 | 0.2238 | 0.0631 | 1.0254 |

Using the 1000 Genomes LD reference panel and SNP list resulted in the fewest number of SNPs remaining after performing LDSC on the input summary statistics, including as few as 250,024 SNPs for OUD. As LDSC accuracy decreases as the number of SNPs decreases,[15] we chose not to progress with the 1000 Genomes reference panels due to the potential for unreliable genetic correlations upon which gSEM models are based. On the other hand, the reference panels generated in MVP resulted in the largest number of remaining SNPs but tended to produce lower heritability estimates than the other reference panels, including a non-significant heritability estimate for MDD. MVP also consistently had the highest inflation in test statistics based on the LDSC intercept. Finally, examining the distribution of the LD scores, MVP LD scores were consistently lower than those using PanUKB. As PanUKB reference panels resulted in an adequate number of SNPs available for analyses, produced significant heritability estimates, showed low inflation in test statistics, and had a broader distribution of LD scores compared to MVP (see below), we conducted African ancestry analyses using PanUKB references.

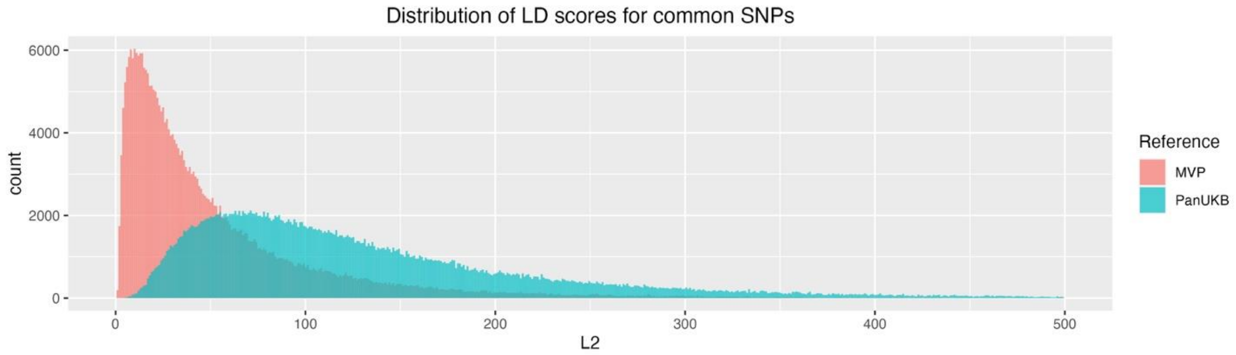

| Comparison across ALL SNPs in both sets | | |
|---|---|---|
| **Statistic** | **MVP L2** | **PanUKB L2** |
| Min | 0.202 | 2.564 |
| 25th percentile | 24.679 | 53.958 |
| Median | 48.104 | 86.065 |
| Mean | 77.451 | 110.042 |
| 75th percentile | 92.463 | 136.970 |
| Max | 3672.834 | 3193.00 |

#### LD Clumping & Identification of Novel Lead SNPs

Following common factor GWAS and GWAS-by-subtraction, LD clumping of summary statistics results was performed using PLINK 1.9[16] with ancestry-matched 1000 Genomes Phase 3 (for European) or PanUKB (for African) reference panels, a significance threshold of $5*10^{-8}$ for index SNPs, $r^2$ threshold of 0.10, and physical distance threshold of 3000kb. For common factor GWAS, SNPs were considered not have been identified by any input GWAS if they were not located within +-1000kb of any lead SNP from any input study for the corresponding common factor. Lead SNPs from input studies were obtained from the supplementary materials for each input GWAS.

To determine if a lead SNP from common factor GWAS had previously been associated with any of the input traits by any previous study, a review of GWAS Catalog[17] was conducted. First, common factor GWAS lead SNP chromosome and base-pair information was lifted over from NCBI assembly hg19/GRCh37 to hg38/GRCh38 using the UCSC Genome Browser's LiftOver tool.[18] Then, for each lead SNP, a query of GWAS Catalog was conducted of all GWAS reporting significant SNPs in the range of +-1000kb of the lead SNP's position. The list of trait associations was subsequently reviewed for any terms corresponding to any input traits for the common factor GWAS. If there were no matches, then the SNP was considered novel in that it had not been previously associated with any previous GWAS of the input traits for a common factor at the time the search was conducted.

**SNP-Level PheWAS**

For any novel SNPs that were identified in GWAS, we performed a SNP-level PheWAS using GWAS Atlas.[19] Analyses examined 4,756 publicly available GWASs and used a Bonferroni corrected p-value of $1.05*10^{-5}$ to identify significant associations.

### Supplementary Figures

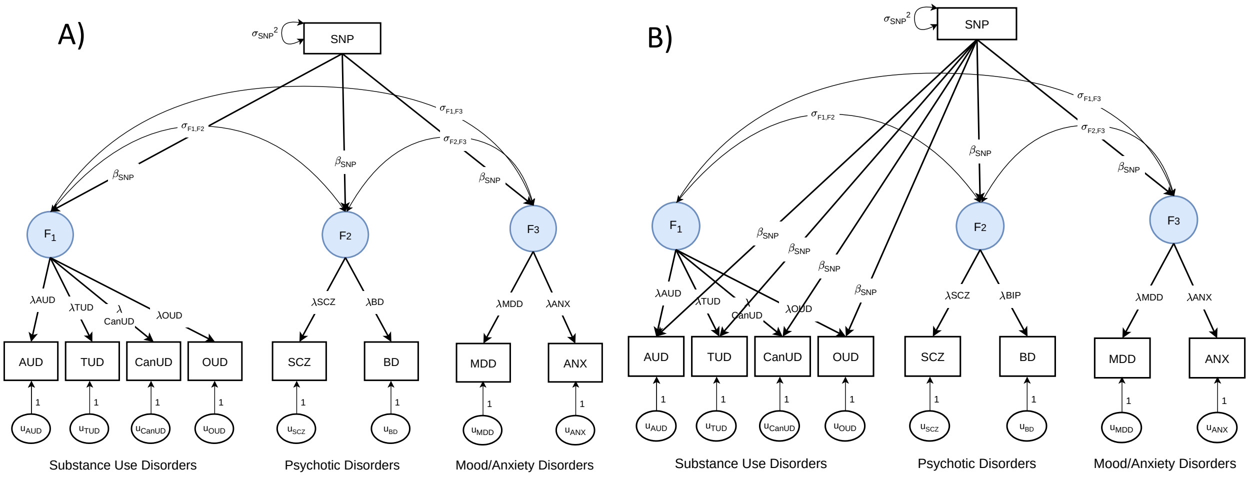

**Supplementary Figure 1. Common and independent pathway models to identify factor specific $Q_{SNPs}$**

Panel A depicts the common pathway model where a given SNP's effects operate through the factors. Panel B depicts the independent pathway model for Factor 1. In this model, each SNP predicts the indicators of Factor 1, as well as the other two factors. A $\chi^2$ difference test was performed for the two models to determine if the SNP's effects could be explained by its association with the factor or, instead, by its association with specific indicators. Follow-up independent pathway models (as shown in Panel B) were run for the each of the other two first-order factors to identify their factor-specific $Q_{SNPs}$. An analogous approach was applied for the second-order factors and for African ancestry models. SNPs whose $\chi^2$ p-value was < $5*10^{-8}$ were removed from summary statistics prior to performing downstream analyses.

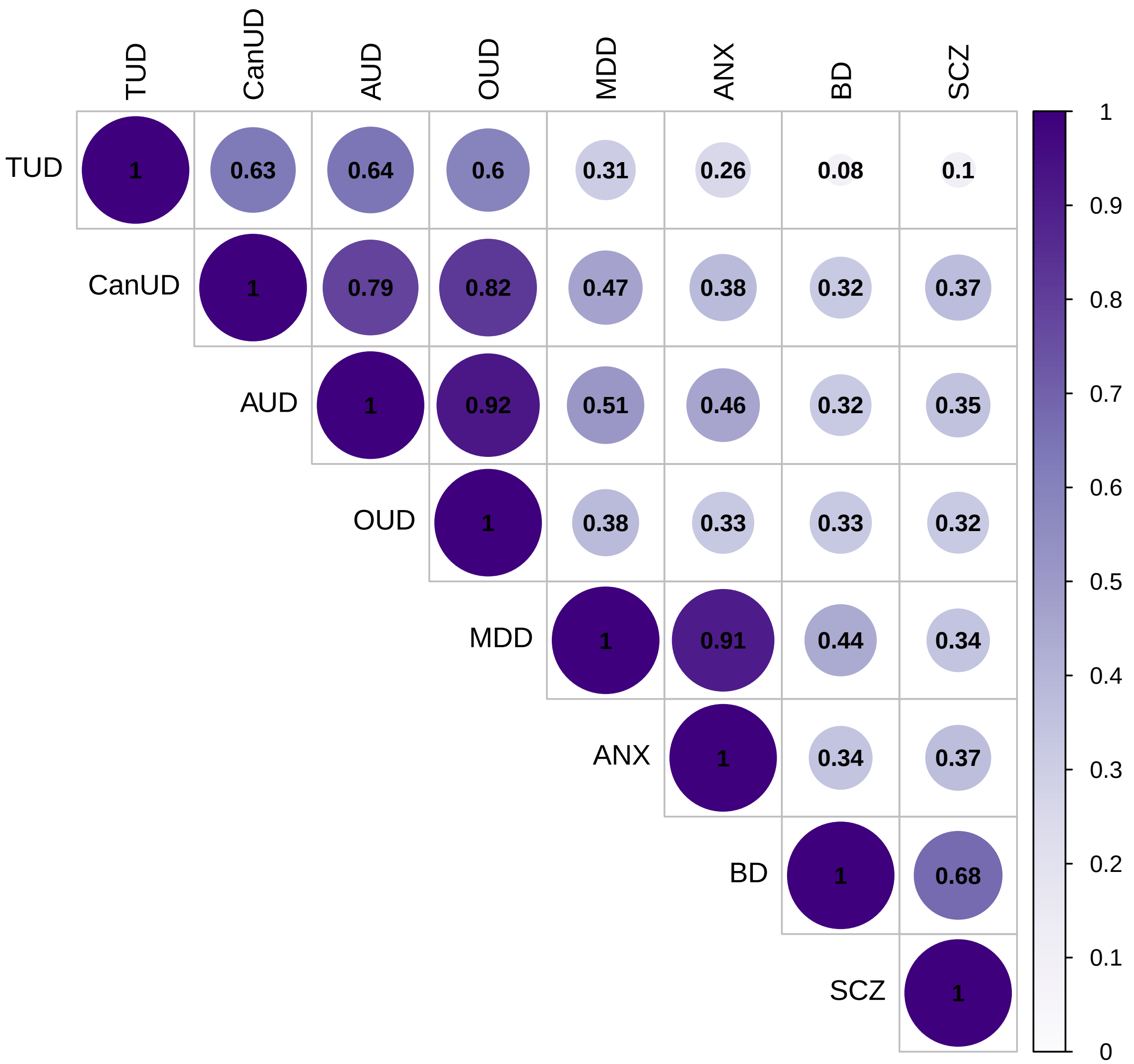

**Supplementary Figure 2. Genetic correlations of input GWAS in European ancestry individuals**

AUD = alcohol use disorder, CanUD = cannabis use disorder, TUD = tobacco use disorder, OUD = opioid use disorder, MDD = major depressive disorder, BD = bipolar disorder, ANX = anxiety disorders, SCZ = schizophrenia. Traits are ordered based on hierarchical clustering.

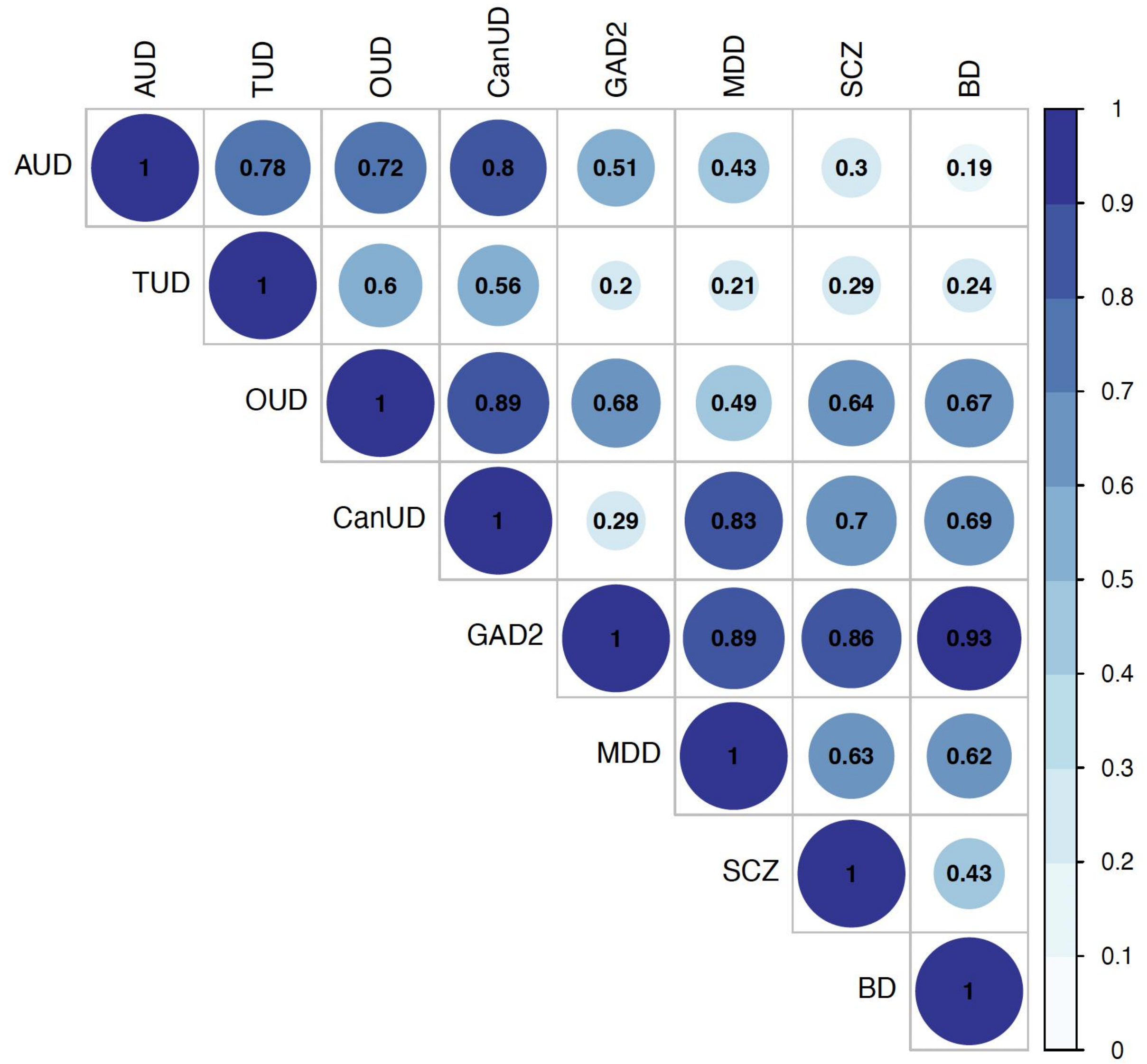

**Supplementary Figure 3. Genetic correlations of input GWAS in African ancestry individuals**

MDD = major depressive disorder, BD = bipolar disorder, GAD-2 = Generalized Anxiety Disorder-2 scores, SCZ = schizophrenia, AUD = alcohol use disorder, TUD = tobacco use disorder, CanUD = cannabis use disorder, OUD = opioid use disorder. Traits are ordered based on hierarchical clustering.

rs75174029

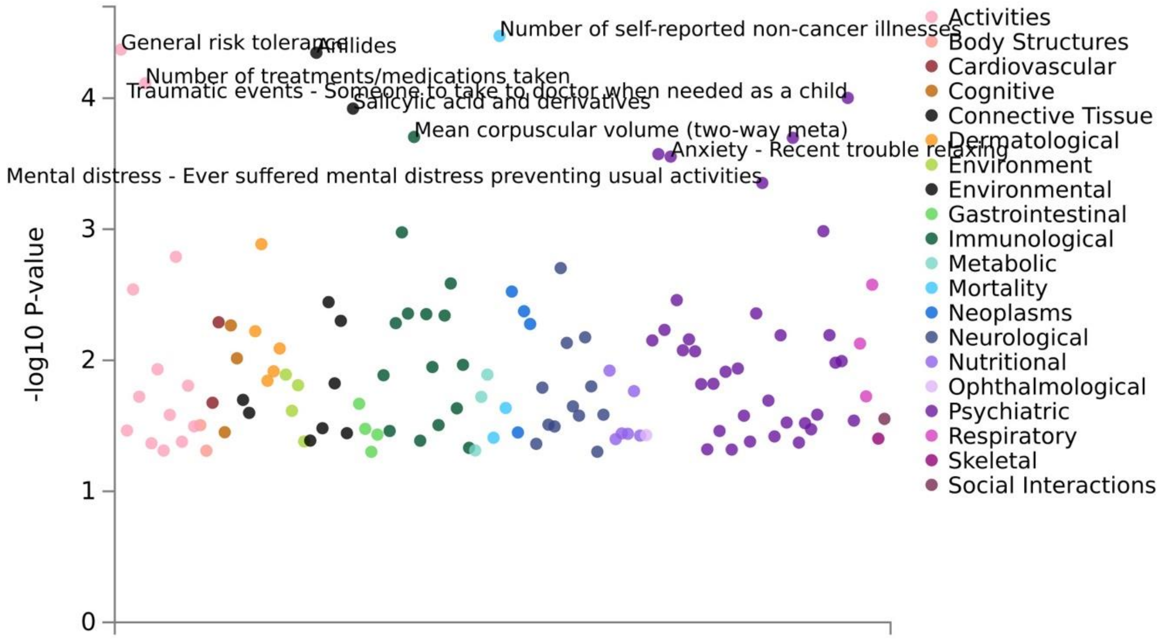

rs7652704

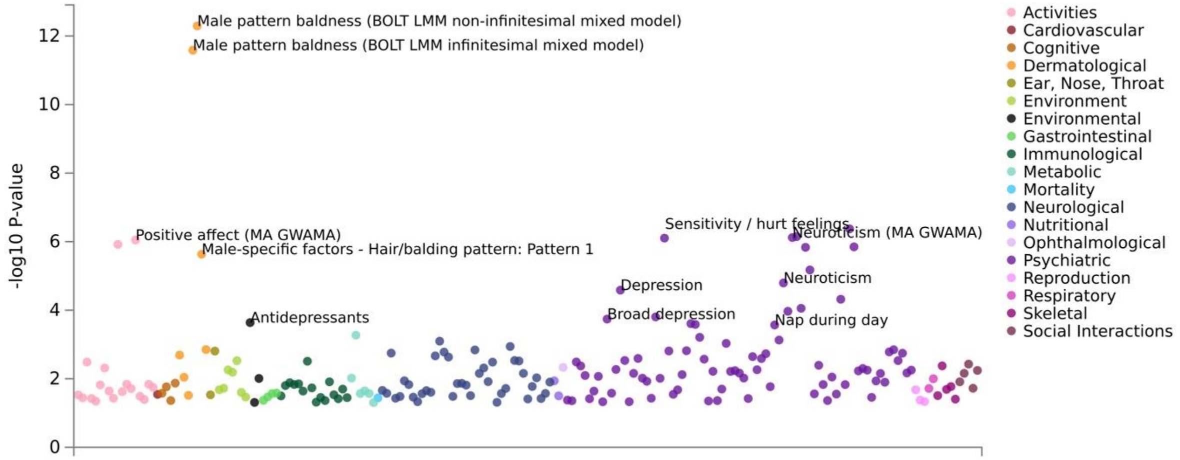

**Supplementary Figure 4. PheWAS plots of novel SNPs for the mood disorders common factor**

PheWAS plots were produced using GWAS Atlas.

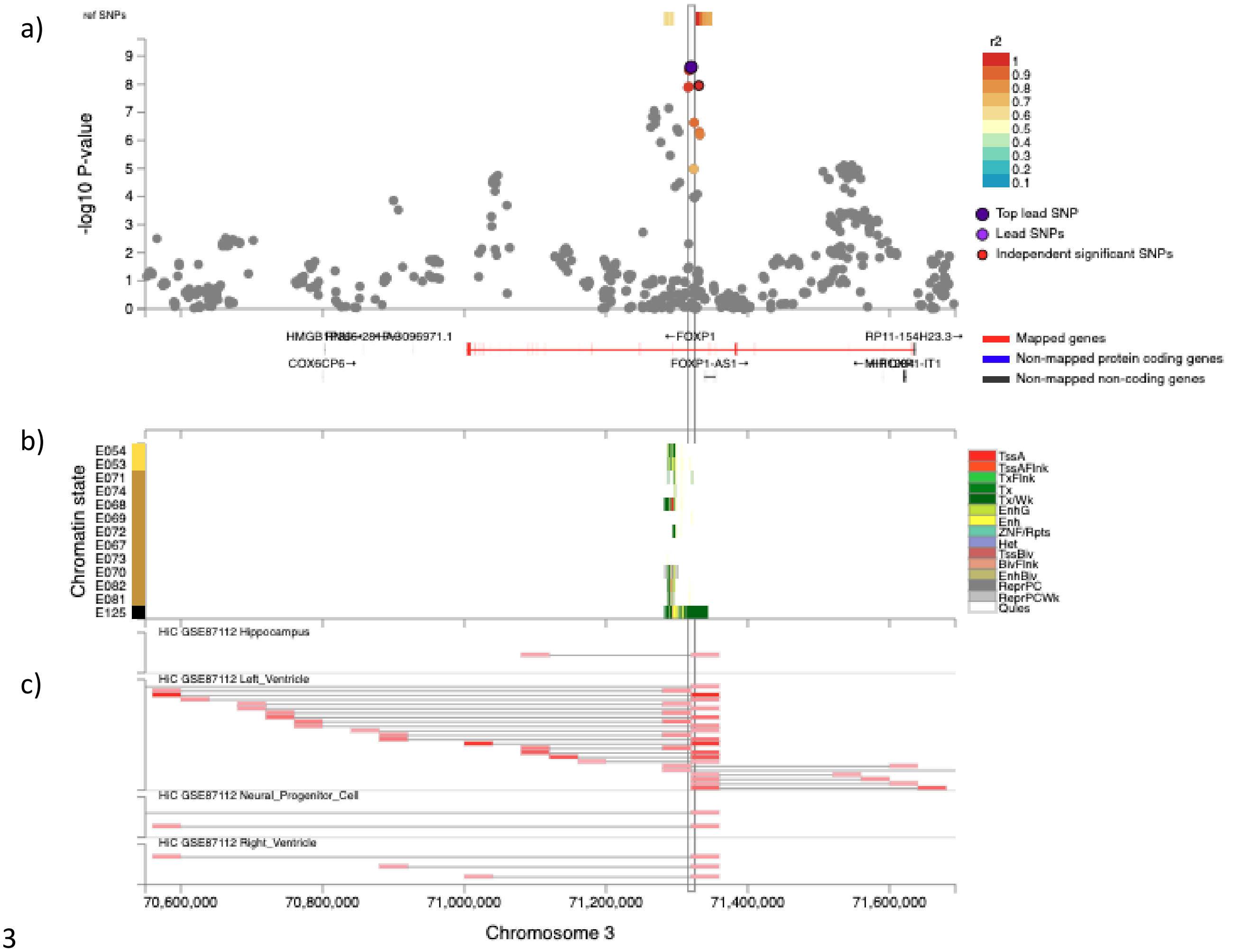

**Supplementary Figure 5. Regional annotation plot for rs75174029, a novel SNP identified by the European ancestry mood/anxiety disorders GWAS.**
**(a)** rs75174029 (in purple), its linked SNPs, and their position relative to genes. rs75174029's predicted genomic target *FOXP1* is shown in red. **(b)** Colocalization of rs75174029 with ROADMAP 15 core chromatin states (right-hand key) in 15 brain tissues (left hand key). E054 = ganglion eminence-derived neurospheres, E053 = cortex-derived neurospheres, E071 = hippocampus, E074 = substantia nigra, E068 = anterior caudate, E069 = cingulate gyrus, E072 = inferior temporal lobe, E067 = angular gyrus, E073 = dorsolateral prefrontal cortex, E070 = germinal matrix, E082 = female fetal brain, E081 = fetal male brain, E125 = NH-A astrocytes. TssA = Active Transcription Start Site, TsAFlnk = flanking active TSS, TxFlnk = transcribed at gene 5' and 3', Tx = strong transcription, TxWk = weak transcription, EnhG = genic enhancers, Enh = enhancers, ZNF/Rpts = ZNF genes and repeats, Het = heterochromatin, TssBiv = bivalent/poised TSS, BivFlnk = Flanking bivalent TSS/Enh, EnhBiv = bivalent enhancer, ReprPC = repressed PolyComb, PreprPCWk = weak repressed PolyComb, Quies = quiescent/low. **(c)** Colocalization with Hi-C signal in brain tissues. Each line represents an interaction, with the two red regions representing the loci which make contact.

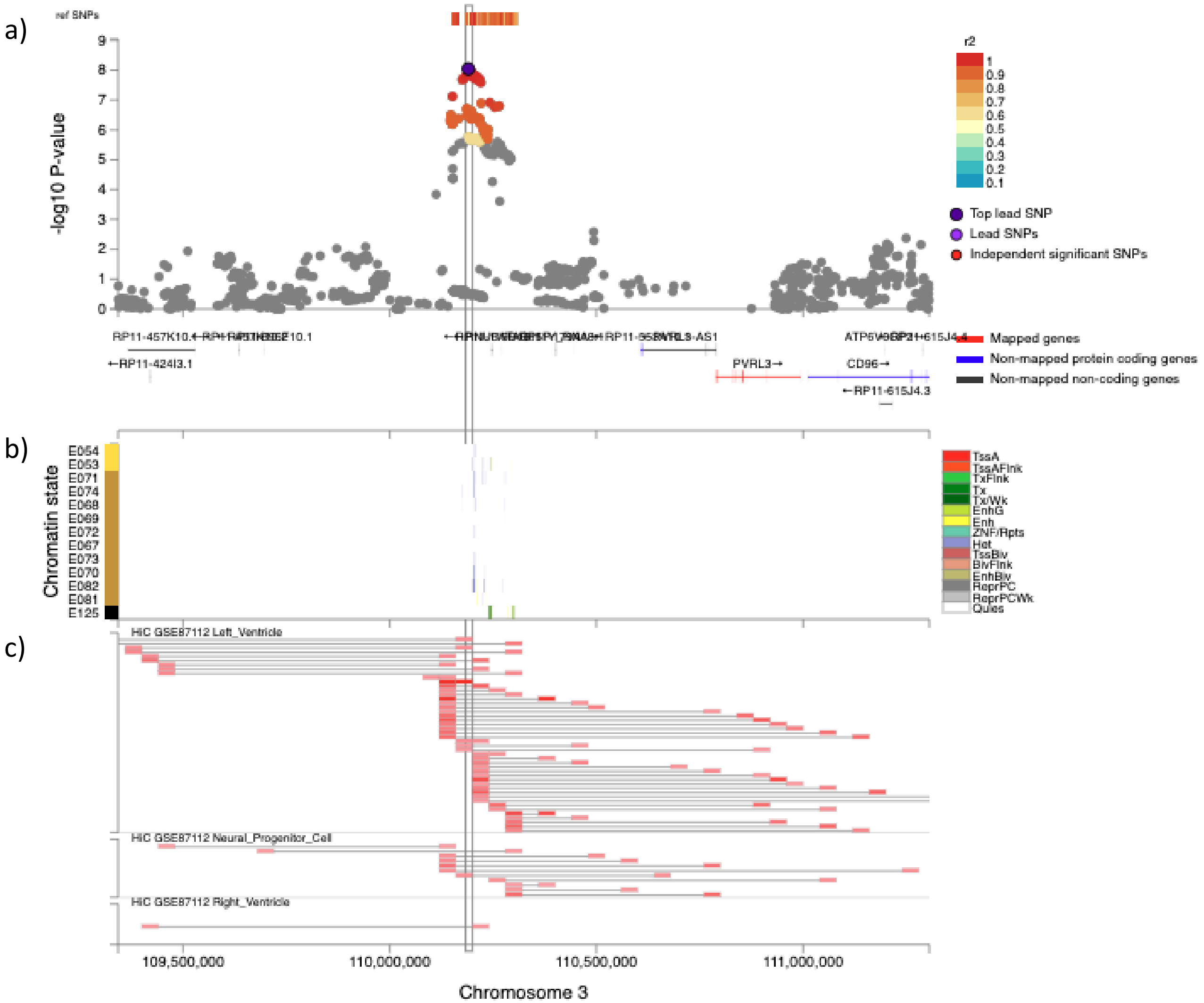

**Supplementary Figure 6. Regional annotation plot for rs7652704, a novel SNP identified by the European ancestry mood/anxiety disorders GWAS.** **(a)** rs7652704 (in purple), its linked SNPs, and their position relative to genes. rs7652704's predicted genomic target *PVRL3* (*NECTIN3*) is shown in red. **(b)** Colocalization of rs7652704 with ROADMAP 15 core chromatin states (right-hand key) in 15 brain tissues (left hand key). E054 = ganglion eminence-derived neurospheres, E053 = cortex-derived neurospheres, E071 = hippocampus, E074 = substantia nigra, E068 = anterior caudate, E069 = cingulate gyrus, E072 = inferior temporal lobe, E067 = angular gyrus, E073 = dorsolateral prefrontal cortex, E070 = germinal matrix, E082 = female fetal brain, E081 = fetal male brain, E125 = NH-A astrocytes. TssA = Active Transcription Start Site, TsAFlnk = flanking active TSS, TxFlnk = transcribed at gene 5' and 3', Tx = strong transcription, TxWk = weak transcription, EnhG = genic enhancers, Enh = enhancers, ZNF/Rpts = ZNF genes and repeats, Het = heterochromatin, TssBiv = bivalent/poised TSS, BivFlnk = Flanking bivalent TSS/Enh, EnhBiv = bivalent enhancer, ReprPC = repressed PolyComb, PreprPCWk = weak repressed PolyComb, Quies = quiescent/low. **(c)** Colocalization with Hi-C signal in brain tissues. Each line represents an interaction, with the two red regions representing the loci which make contact.

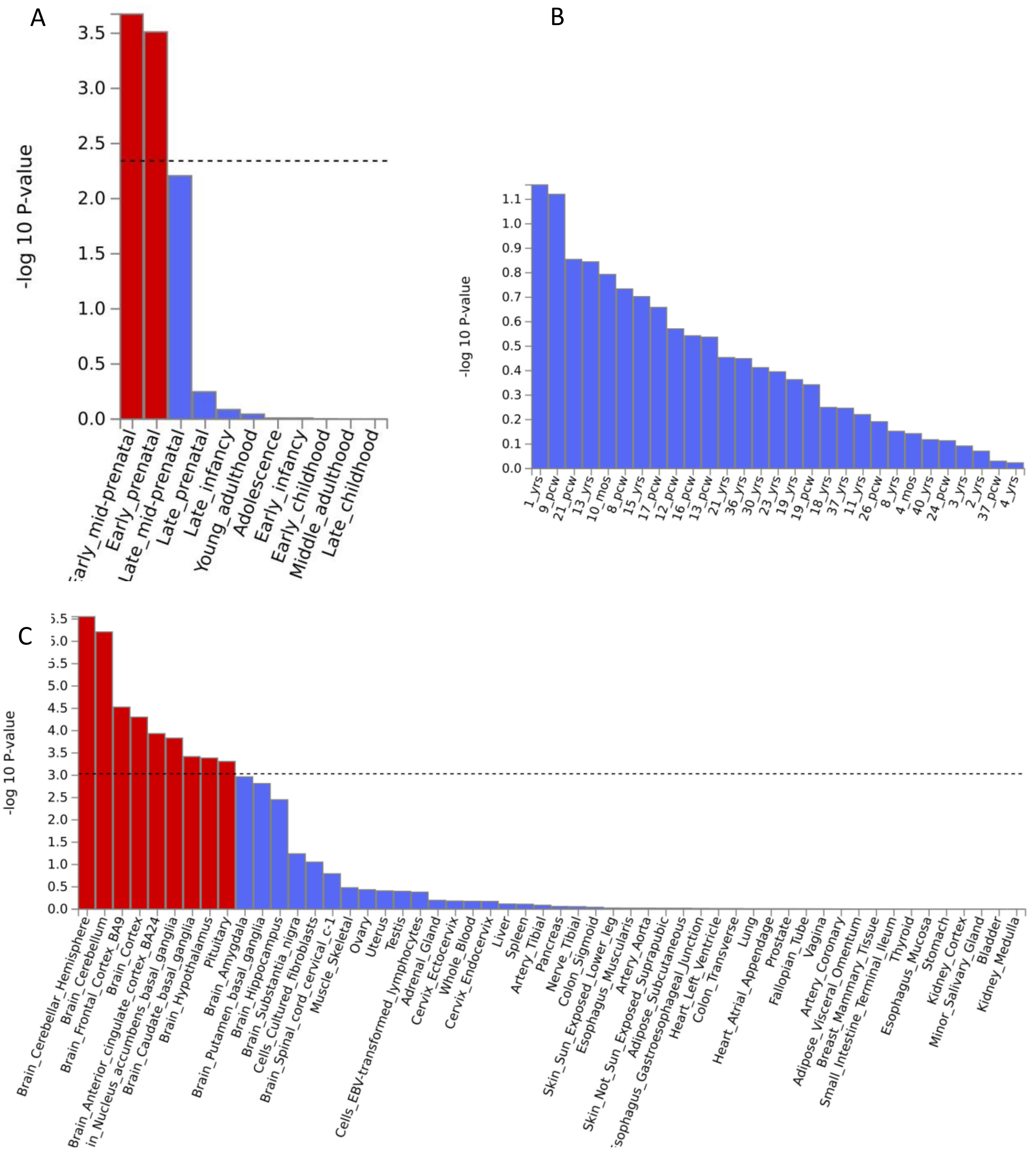

**Supplementary Figure 7. Results of MAGMA tissue expression analysis of EUR substance use disorders factor**

Results for the BrainSpan database are shown in panels A and B, and results for GTEx v8 are shown in Panel C. Dashed line represents significance threshold.

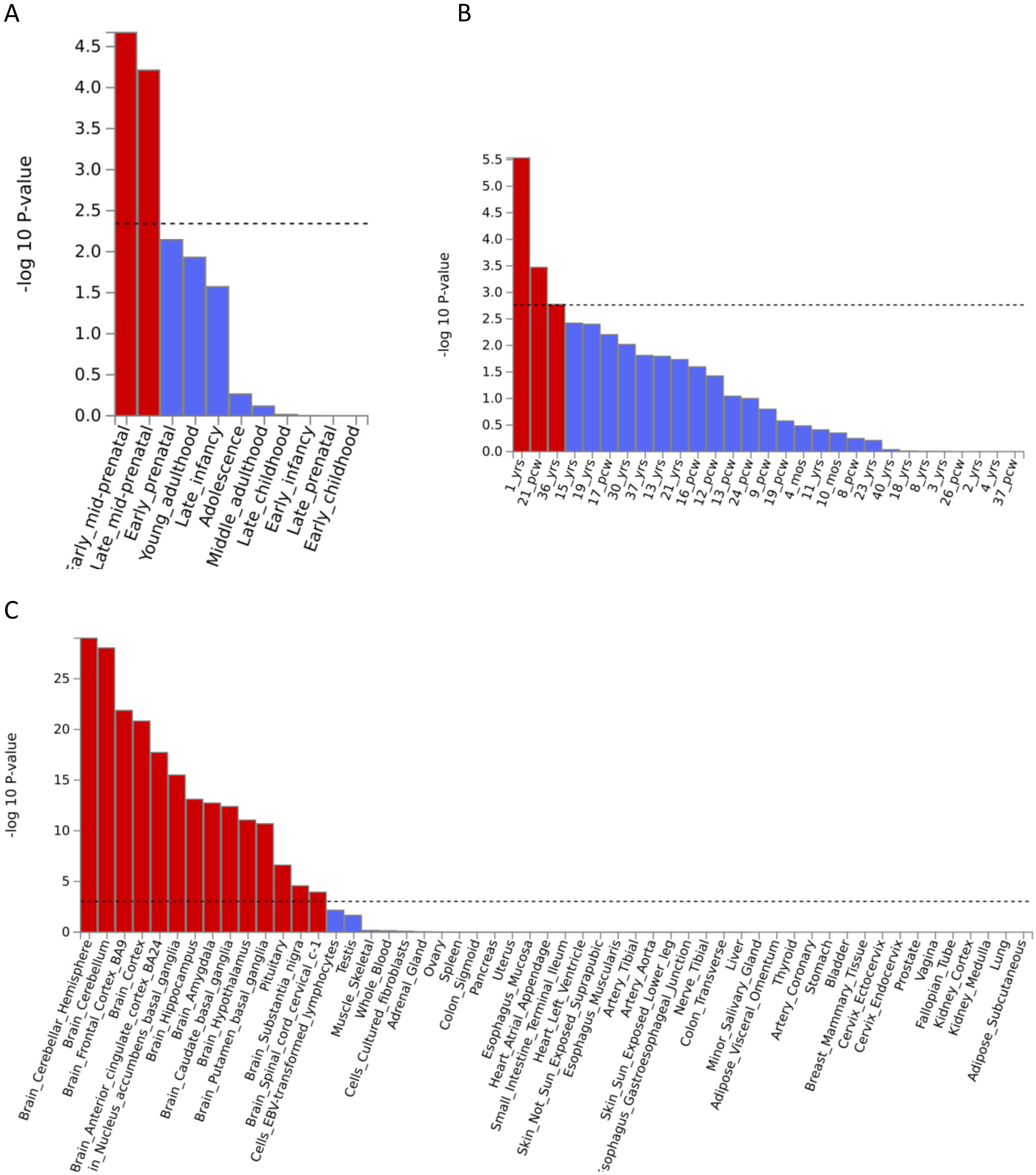

**Supplementary Figure 8. Results of MAGMA tissue expression analysis of EUR psychotic disorders factor**

Results for the BrainSpan database are shown in panels A and B, and results for GTEx v8 are shown in Panel C. Dashed line represents significance threshold.

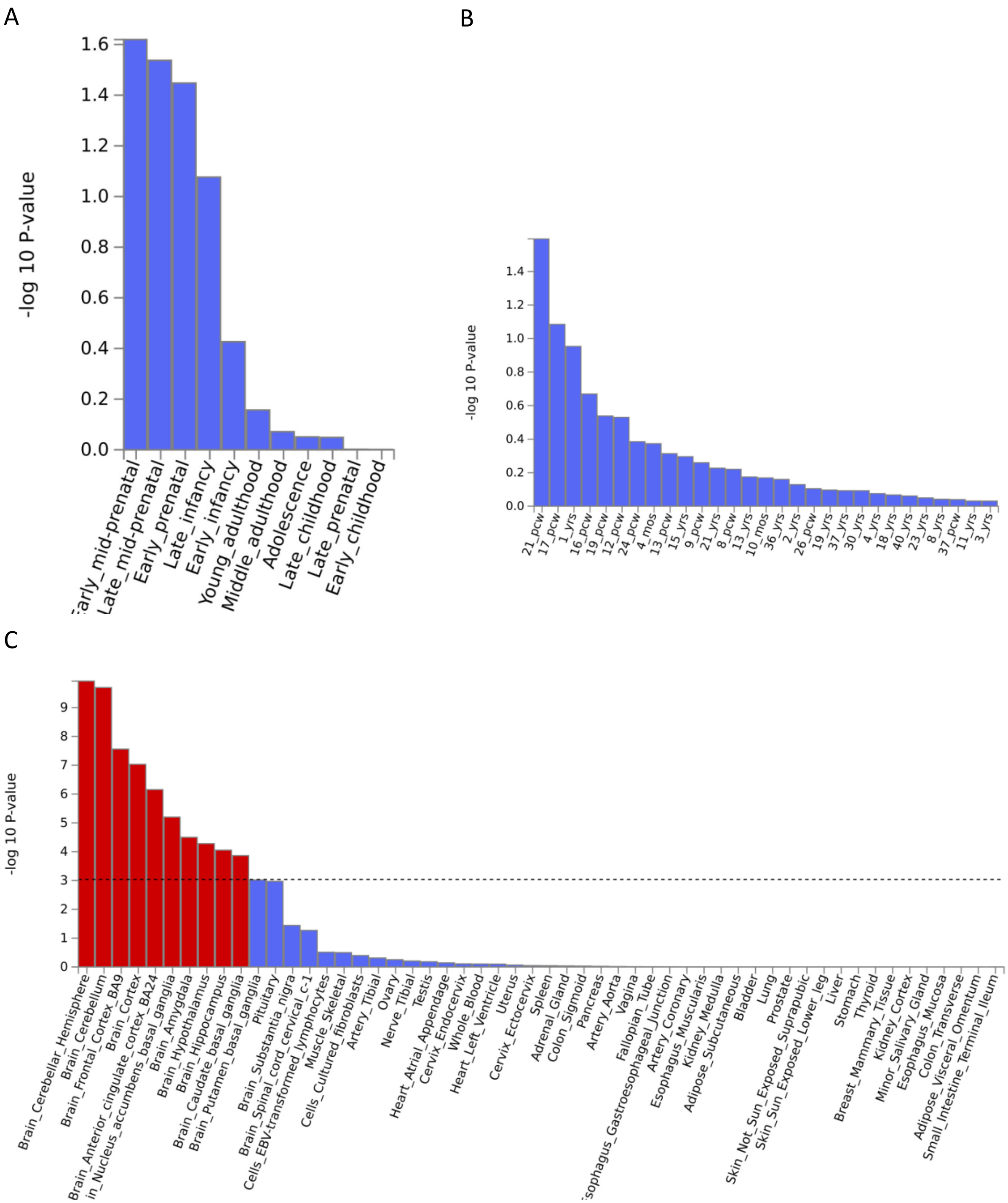

**Supplementary Figure 9. Results of MAGMA tissue expression analysis of EUR mood disorders factor**

Results for the BrainSpan database are shown in panels A and B, and results for GTEx v8 are shown in Panel C. Dashed line indicates significance threshold.

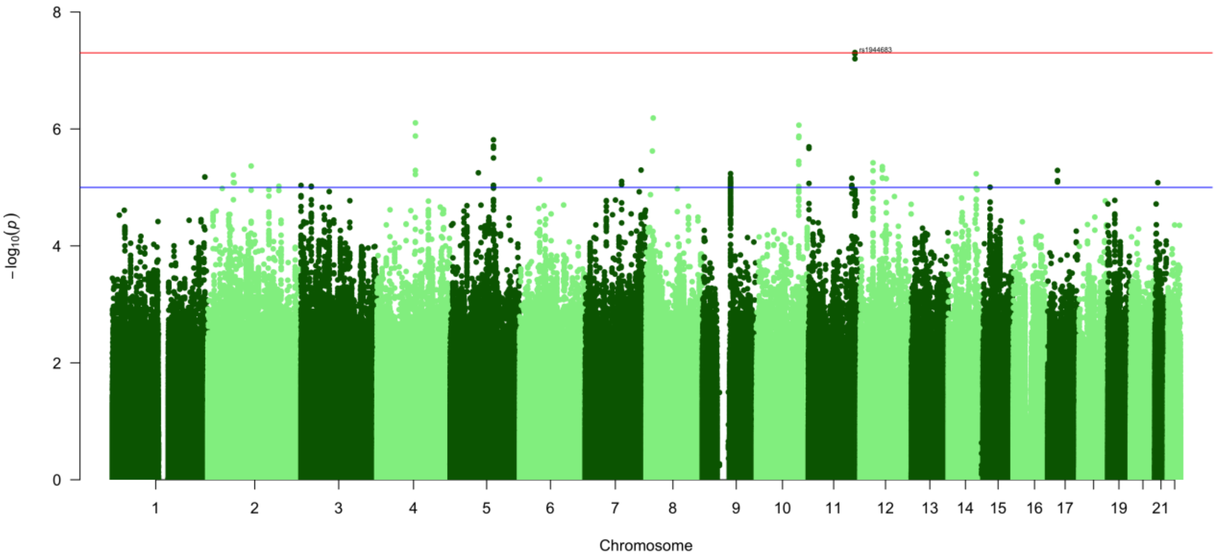

**Supplementary Figure 10. Manhattan plot for substance use disorders factor in AFR ancestry individuals**

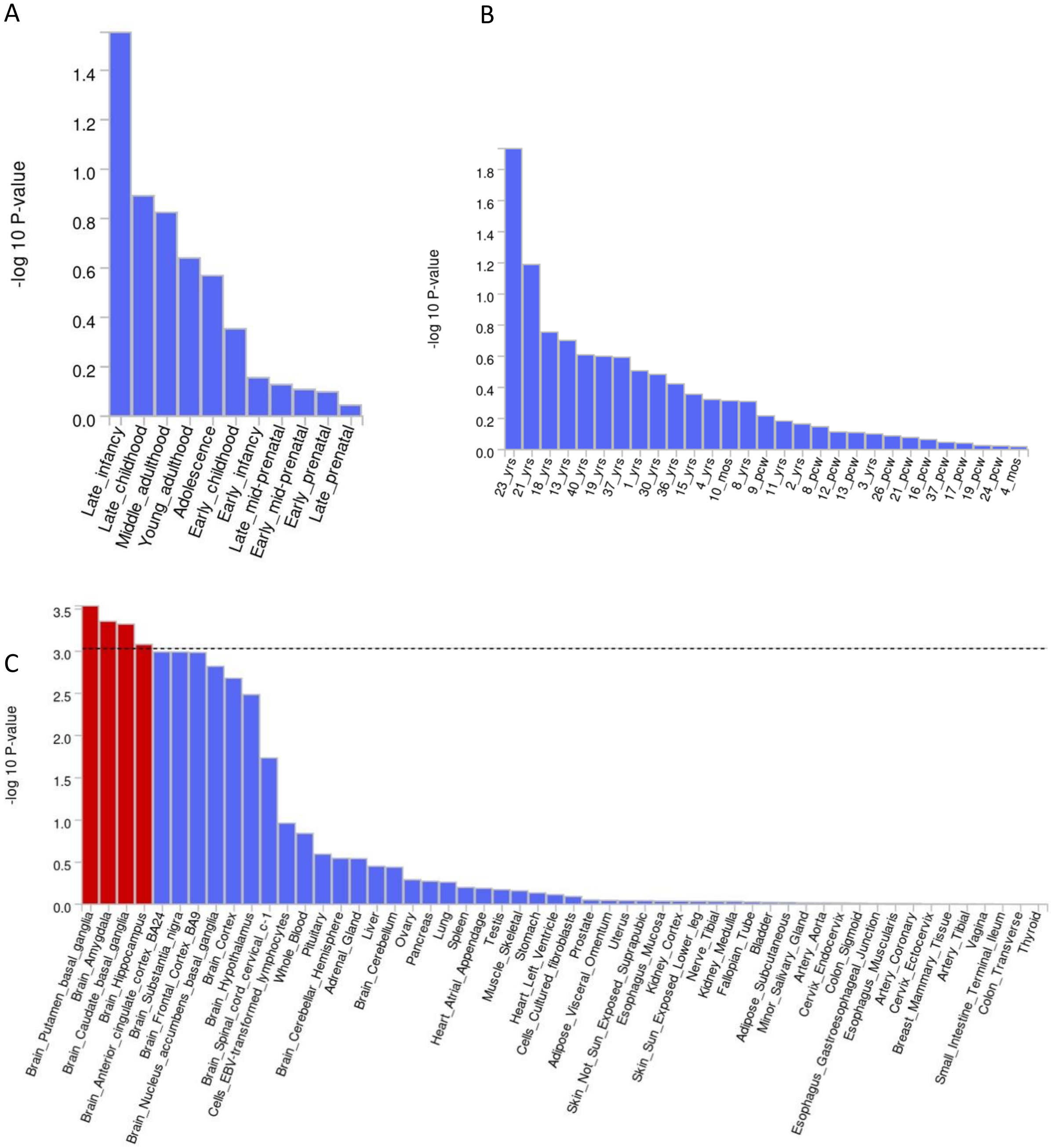

**Supplementary Figure 11. Results of MAGMA tissue expression analysis of AFR ancestry substance use disorders factor**
Results for the BrainSpan database are shown in panels A and B, and results for GTEx v8 are shown in Panel C. Dashed line indicates significance threshold.

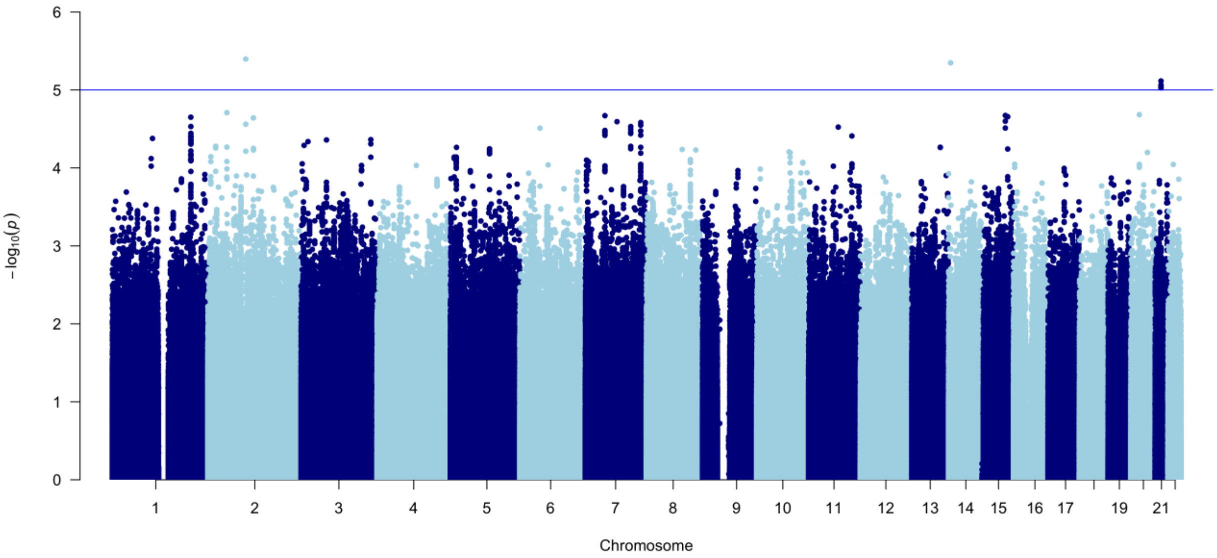

**Supplementary Figure 12. Manhattan plot for psychiatric disorders factor in AFR ancestry individuals**

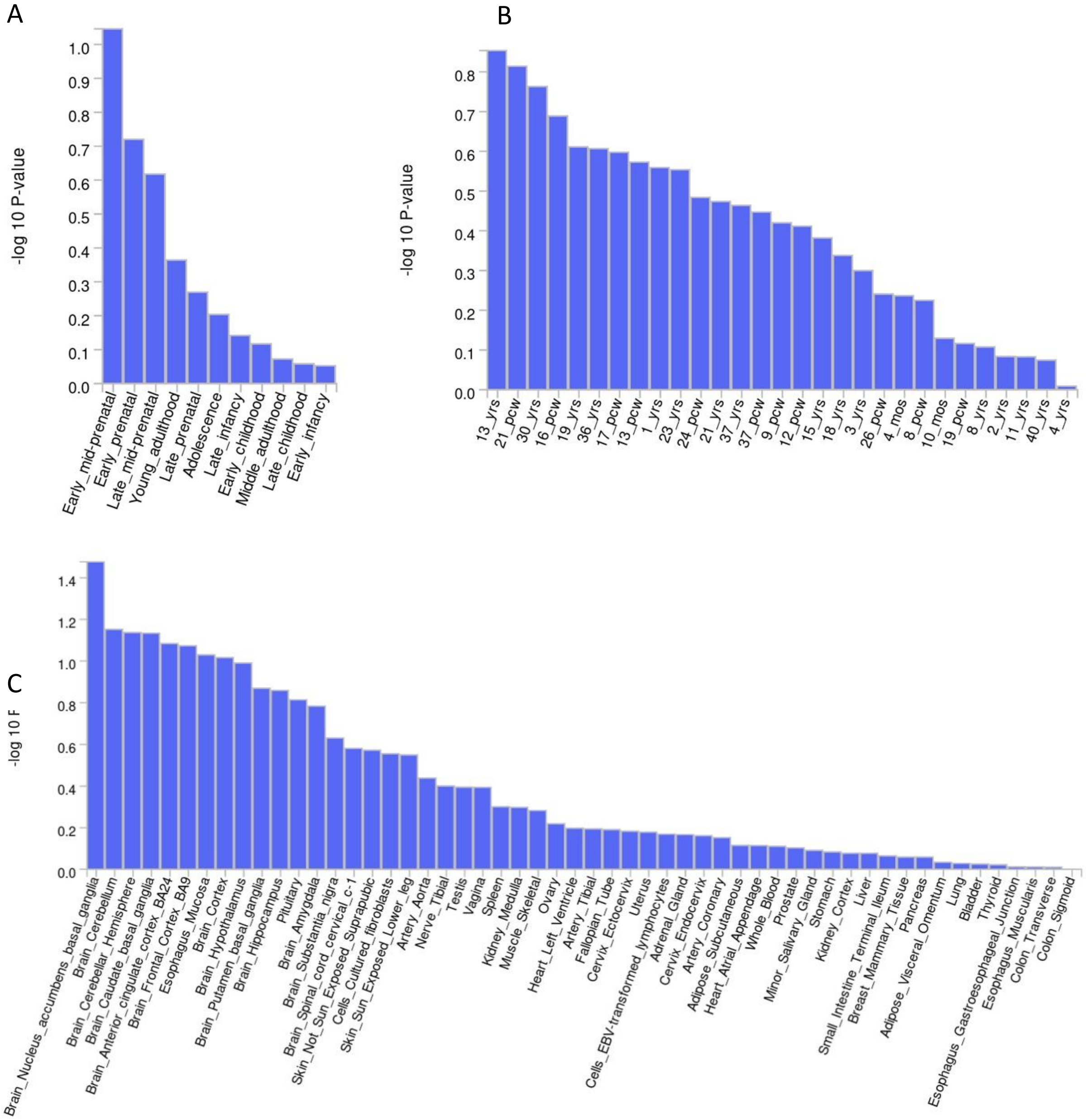

**Supplementary Figure 13. Results of MAGMA tissue expression analysis of AFR ancestry psychiatric disorders factor**

Results for the BrainSpan database are shown in panels A and B, and results for GTEx v8 are shown in Panel C.

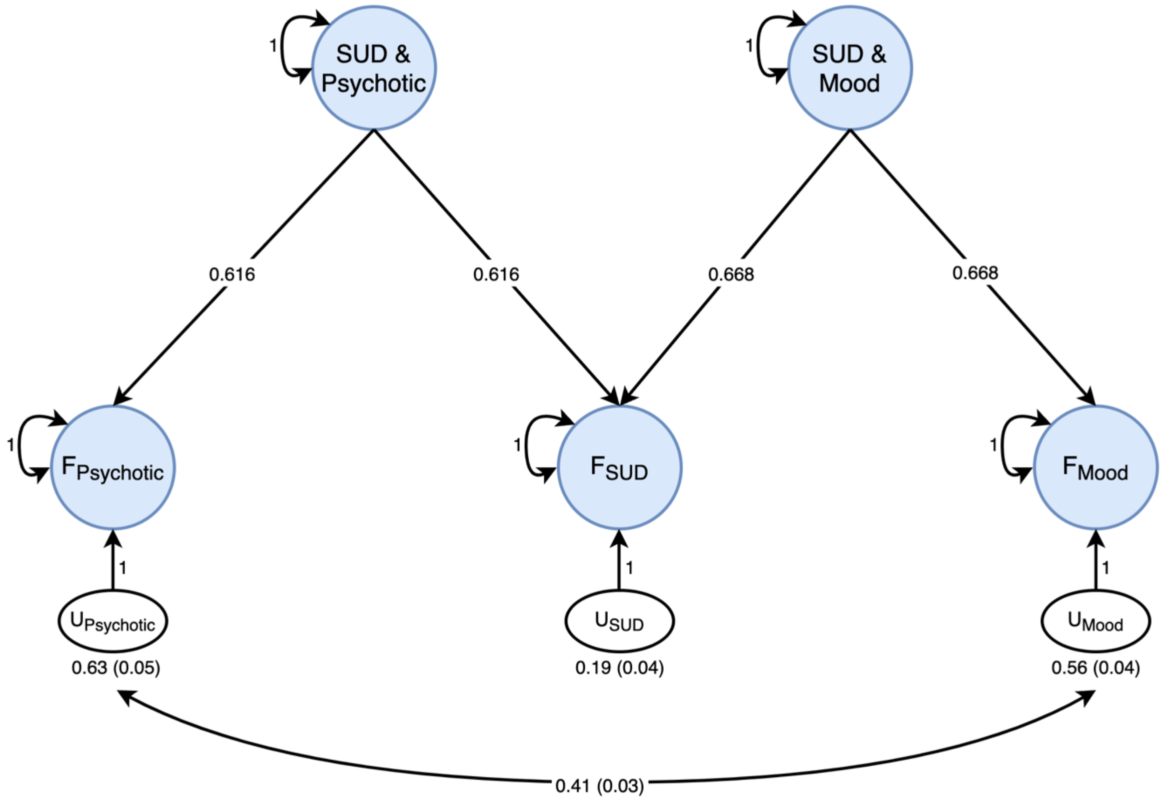

**Supplementary Figure 14. EUR ancestry second order common factor model**

Model fit statistics: $\chi 2(2) = 57.61$, p = $3.09*10^{-13}$, AIC = 65.61, CFI = 0.91, SRMR = 0.07.

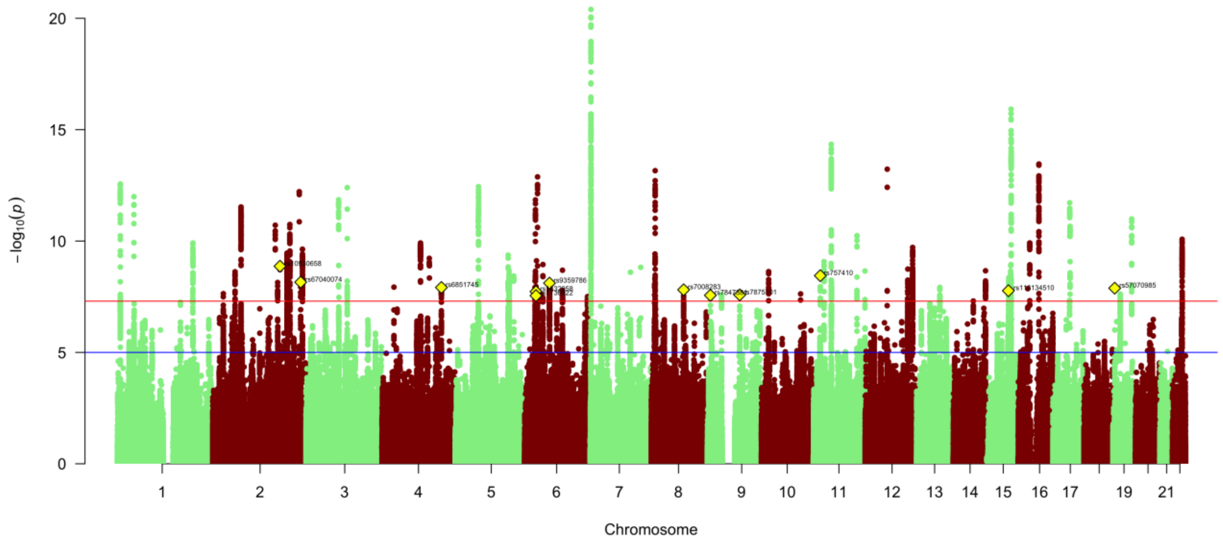

**Supplementary Figure 15. Manhattan plot for second-order common factor representing overlap between substance use and psychotic disorders in EUR ancestry individuals**
GWAS identified 76 lead SNPs, 12 of which were not in any of the input GWAS.

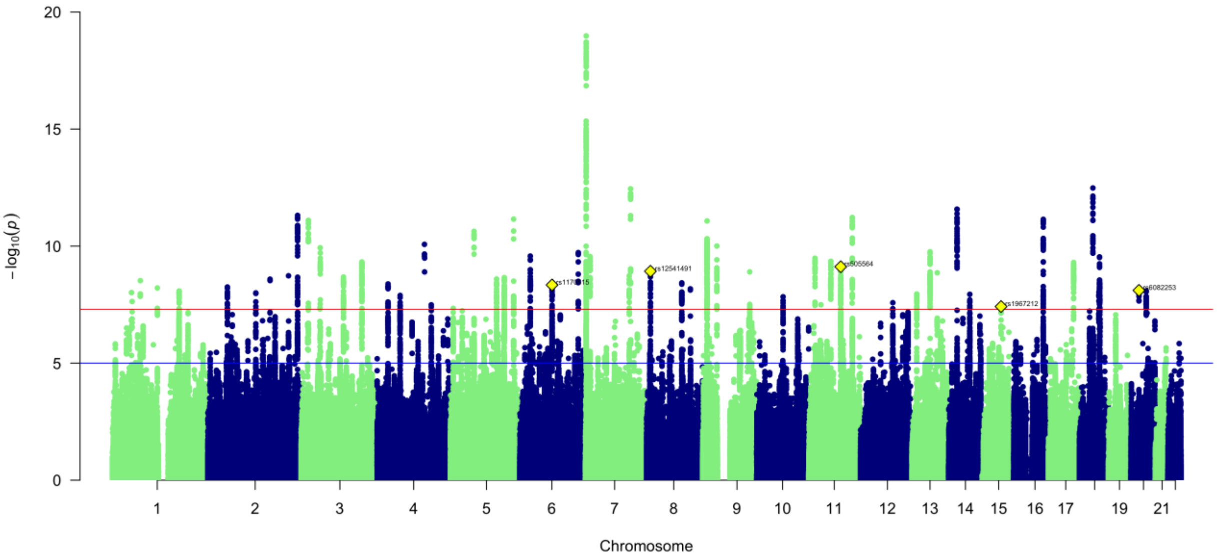

**Supplementary Figure 16. Manhattan plot for second-order common factor representing overlap between substance use and mood/anxiety disorders in EUR ancestry individuals**
GWAS identified 63 lead SNPs, 5 of which were not in any of the input GWAS.

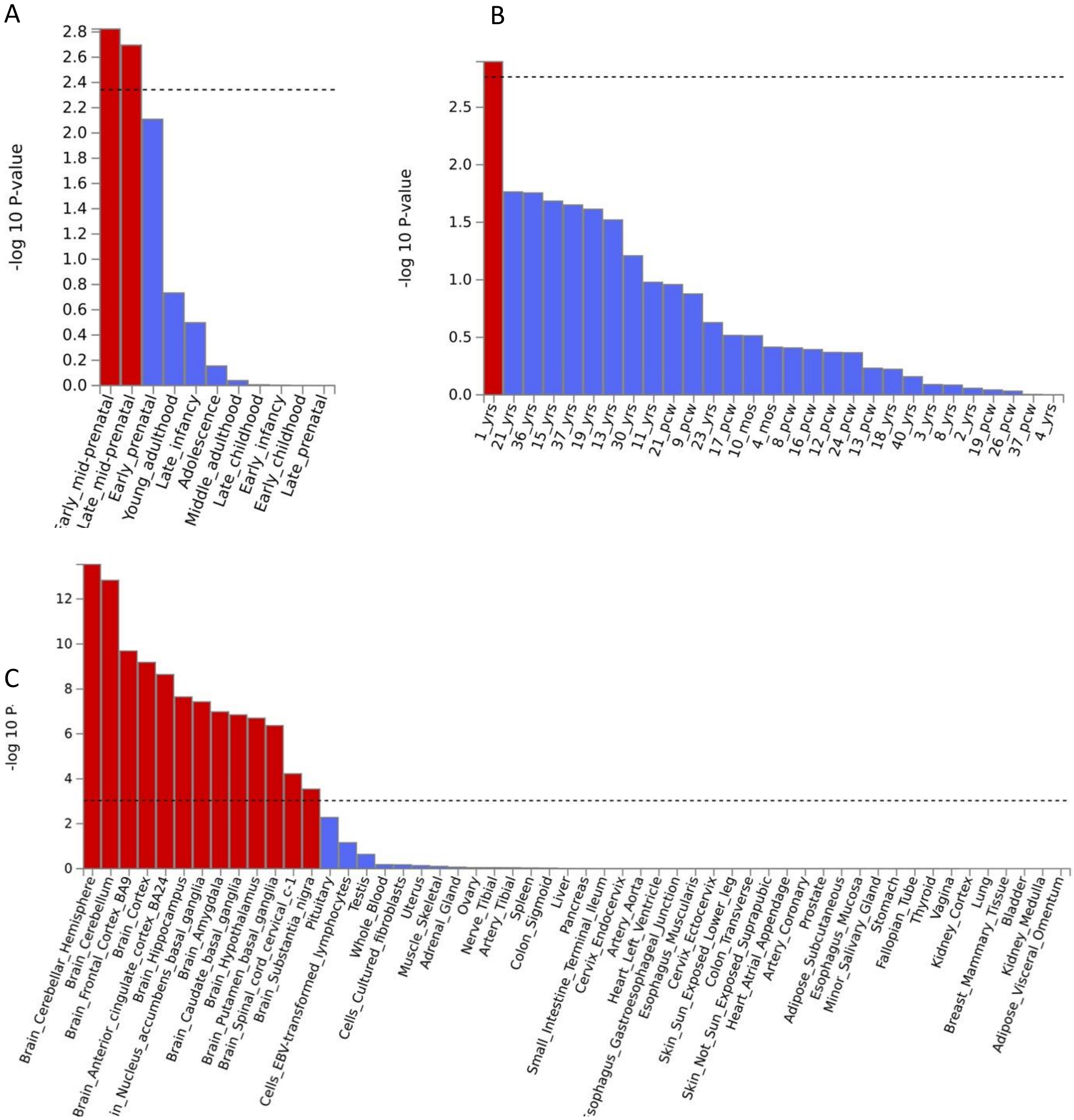

**Supplementary Figure 17. Results of MAGMA tissue expression analysis of EUR ancestry second-order substance use and psychotic disorders factor**

Results for the BrainSpan database are shown in panels A and B, and results for GTEx v8 are shown in Panel C. Dashed line indicates significance threshold.

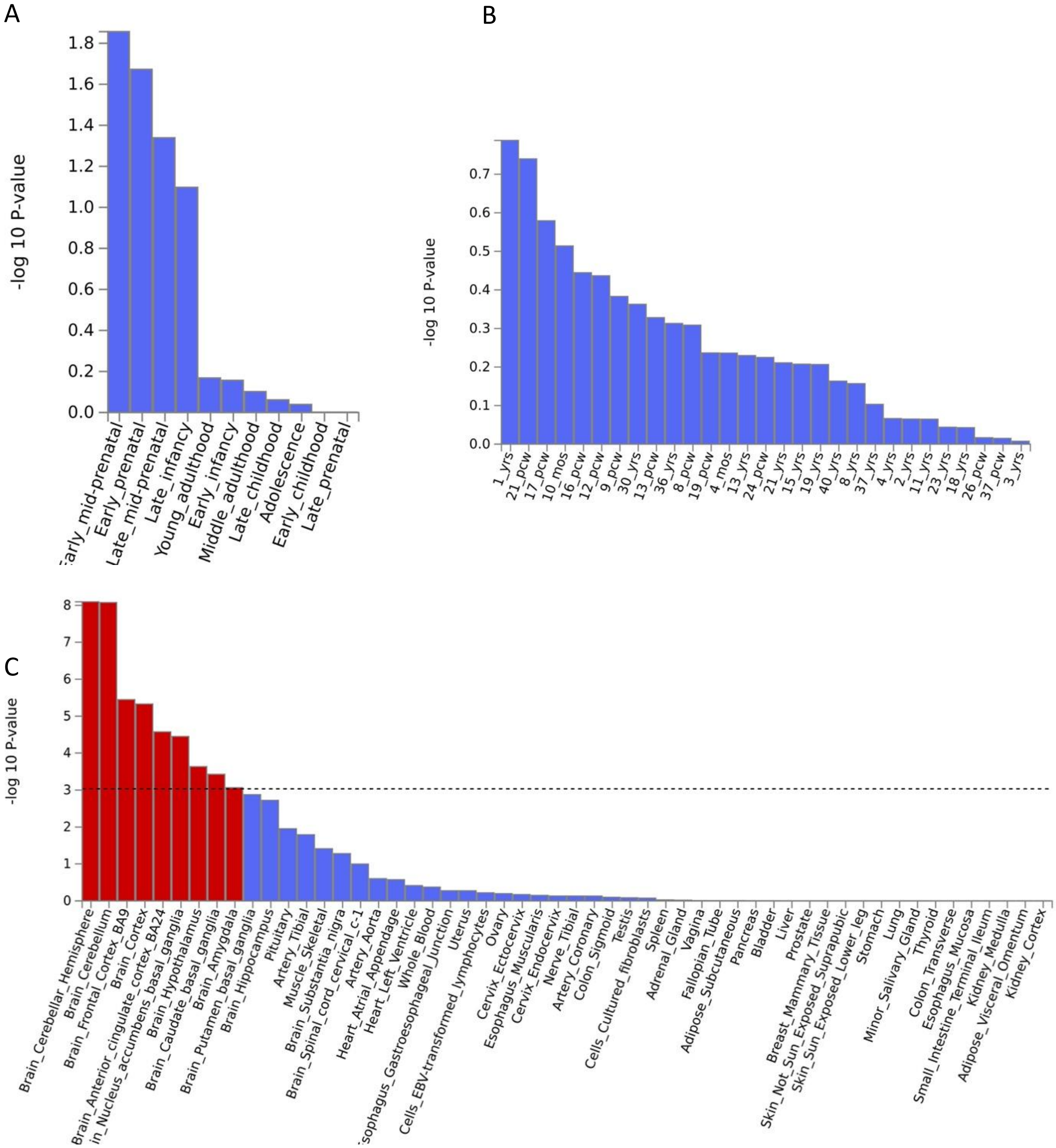

**Supplementary Figure 18. Results of MAGMA tissue expression analysis of EUR ancestry second-order substance use and mood disorders factor**

Results for the BrainSpan database are shown in panels A and B, and results for GTEx v8 are shown in panel C. Dashed line indicates significance threshold.

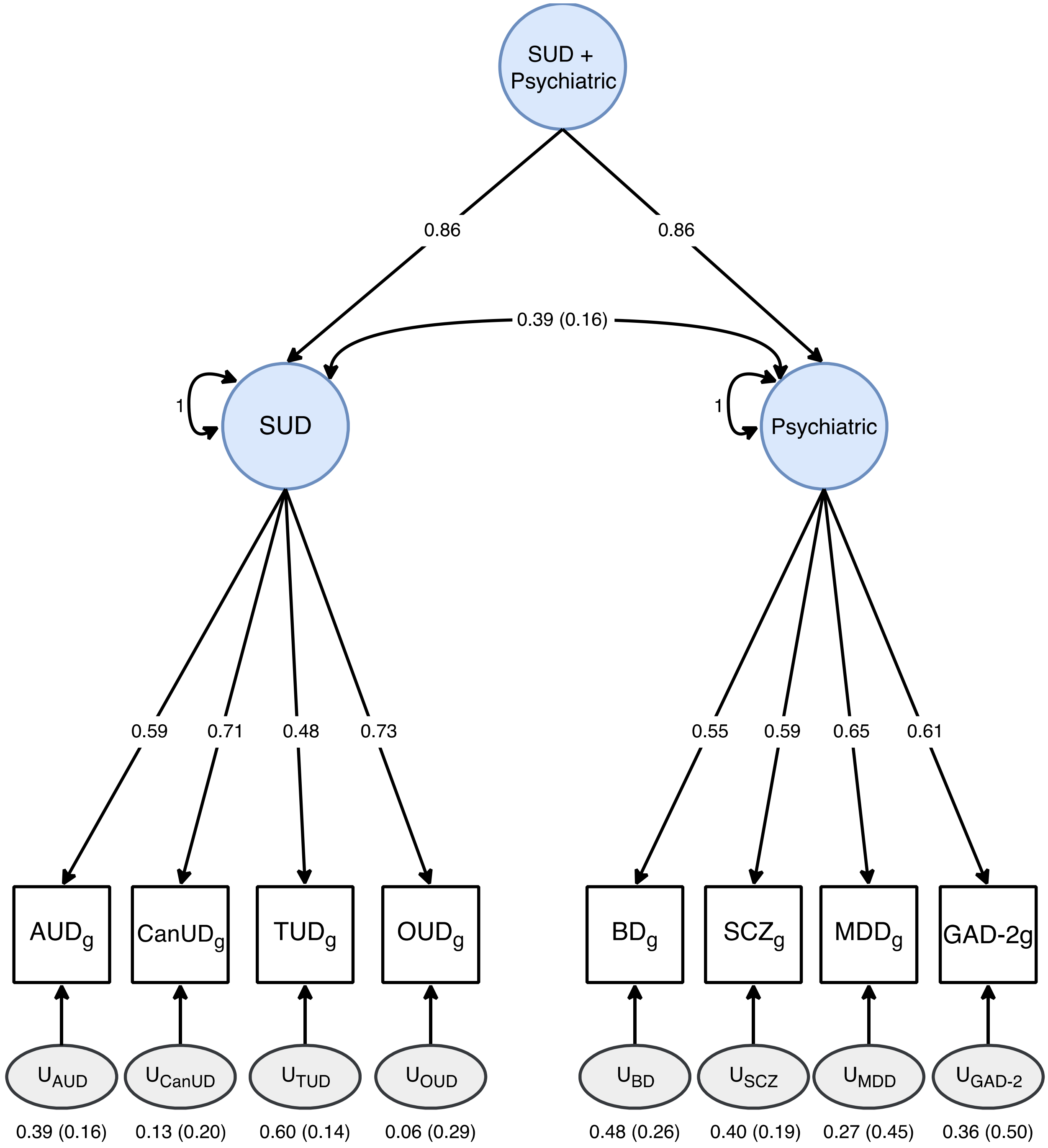

**Supplementary Figure 19. AFR ancestry second order common factor model**
Model fit statistics: $\chi 2(19) = 21.49$, $p = 0.31$, AIC = 55.49, CFI = 0.99, SRMR = 0.10.

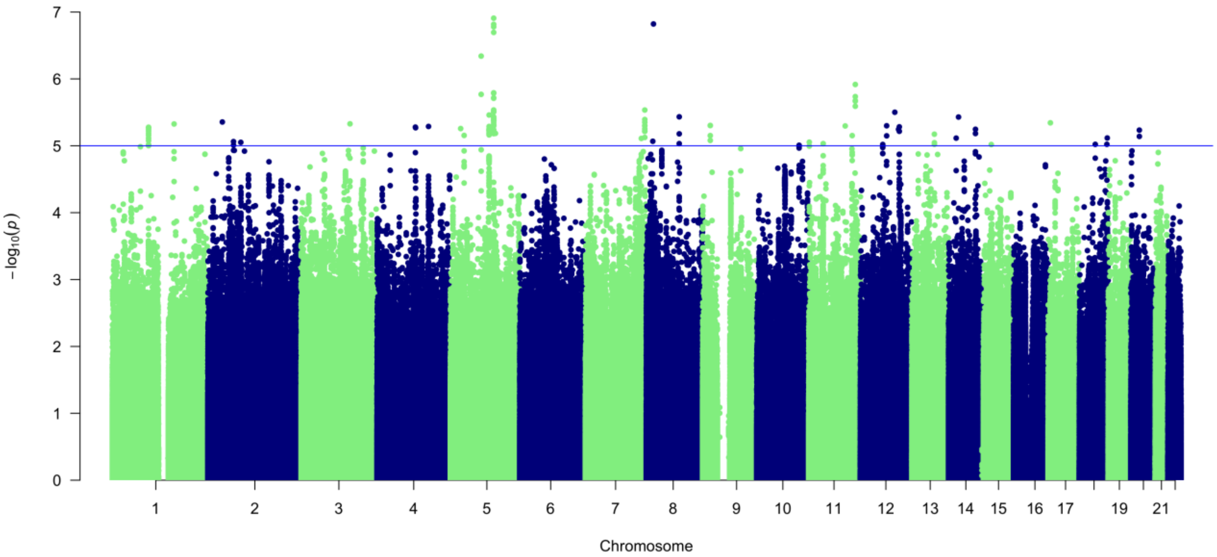

**Supplementary Figure 20. Manhattan plot for second-order common factor representing overlap between substance use and psychiatric disorders in AFR ancestry individuals**

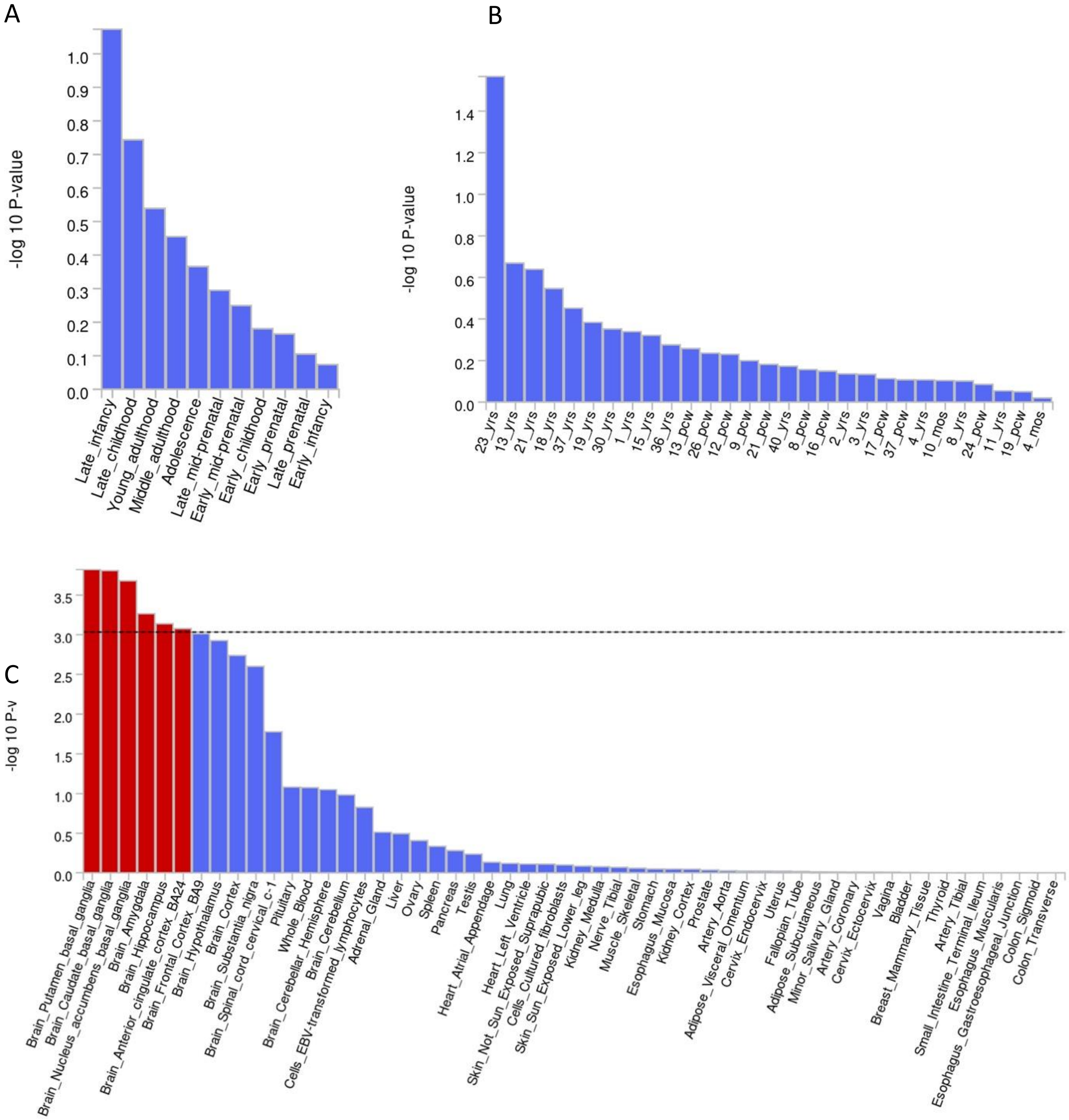

**Supplementary Figure 21. Results of MAGMA tissue expression analysis of AFR ancestry second-order substance use and psychiatric disorders factor**

Results for the BrainSpan database are shown in panels A and B, and results for GTEx v8 are shown in panel C. Dashed line indicates significance threshold.

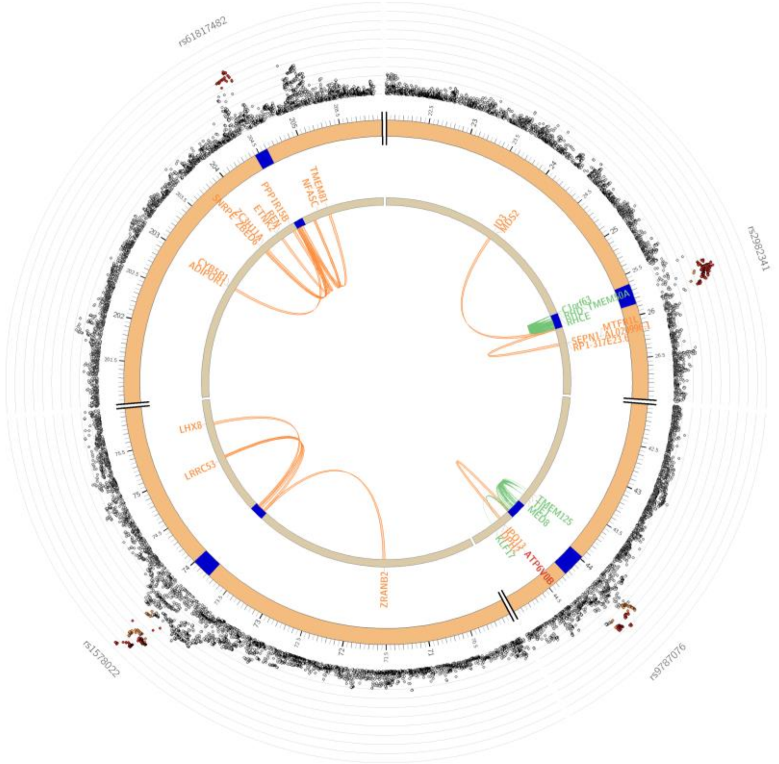

Chromosome 1

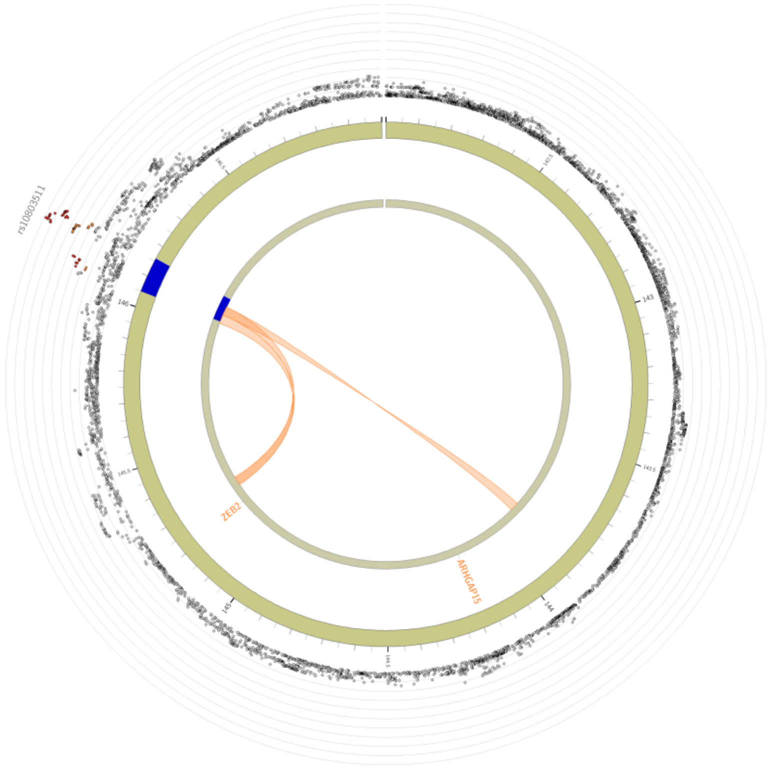

Chromosome 2

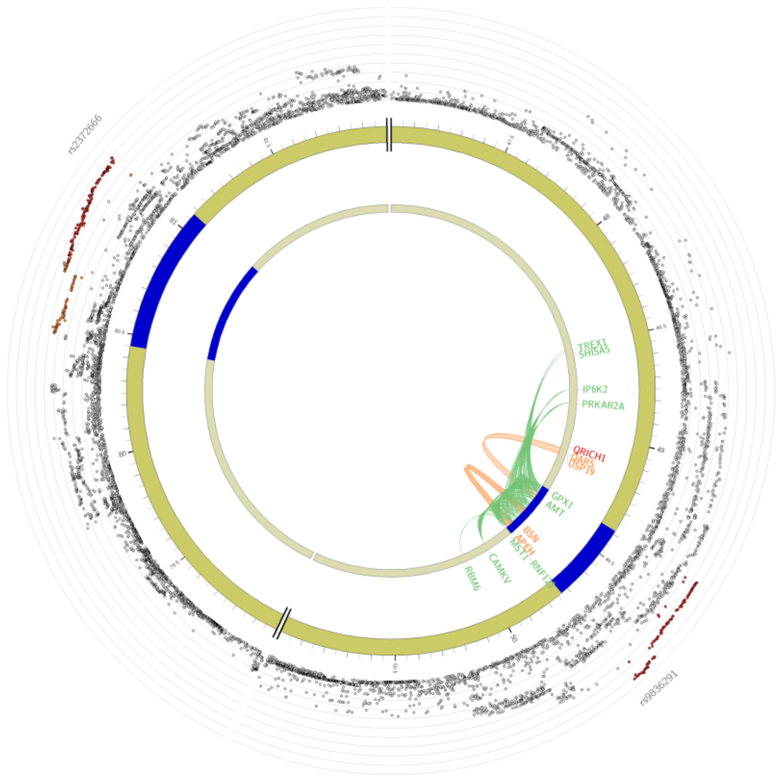

Chromosome 3

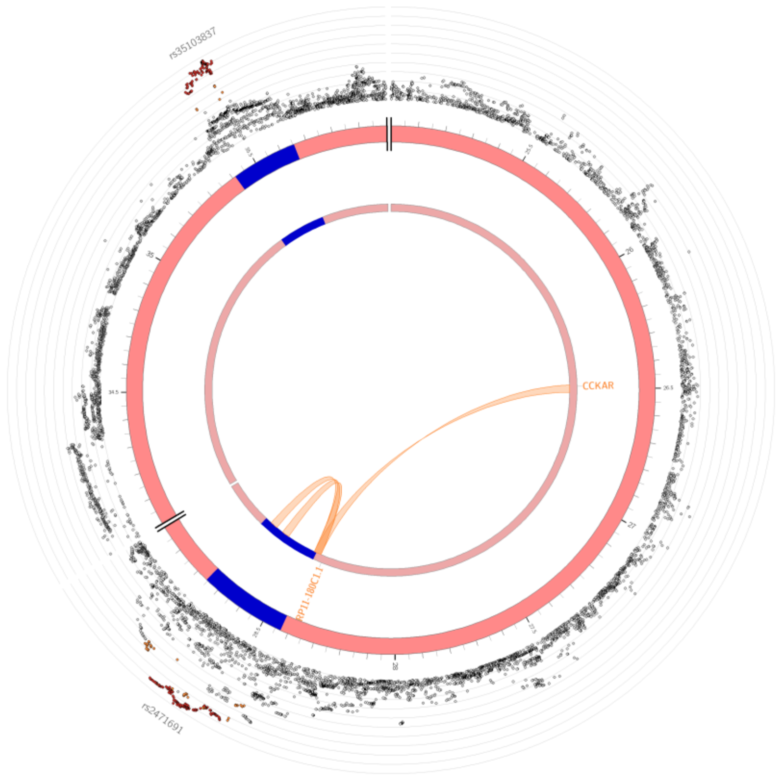

Chromosome 4

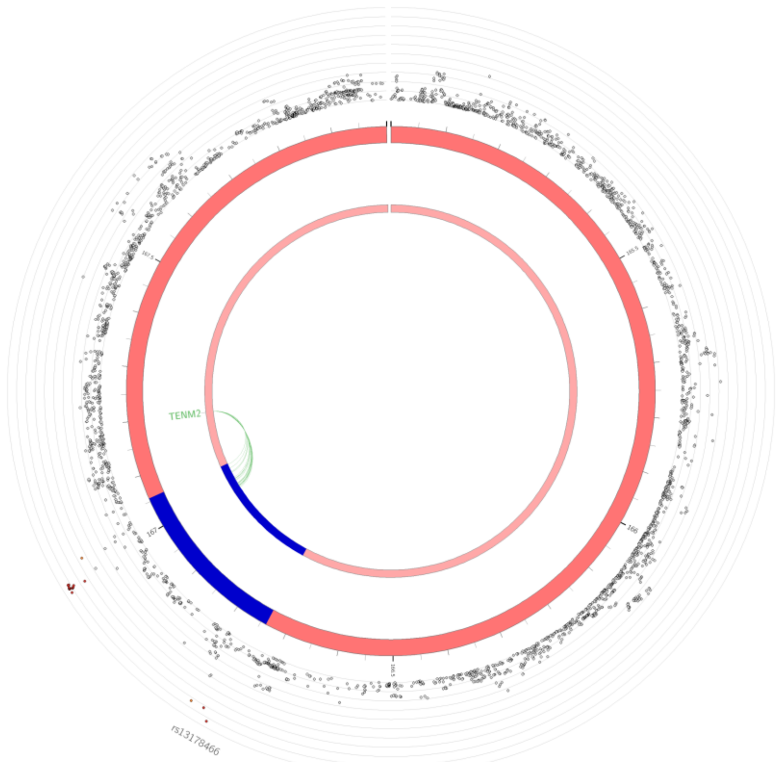

Chromosome 5

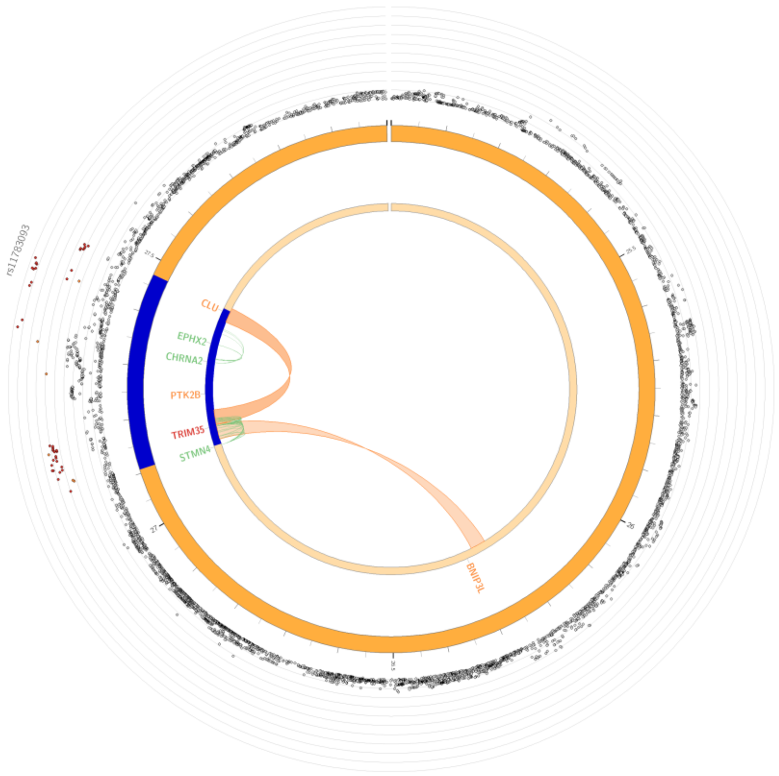

Chromosome 8

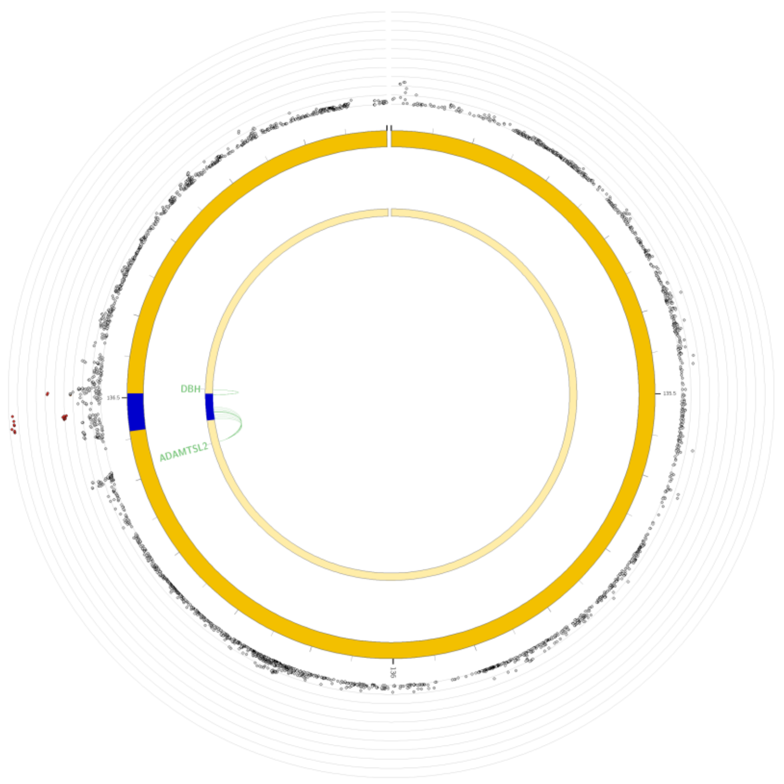

Chromosome 9

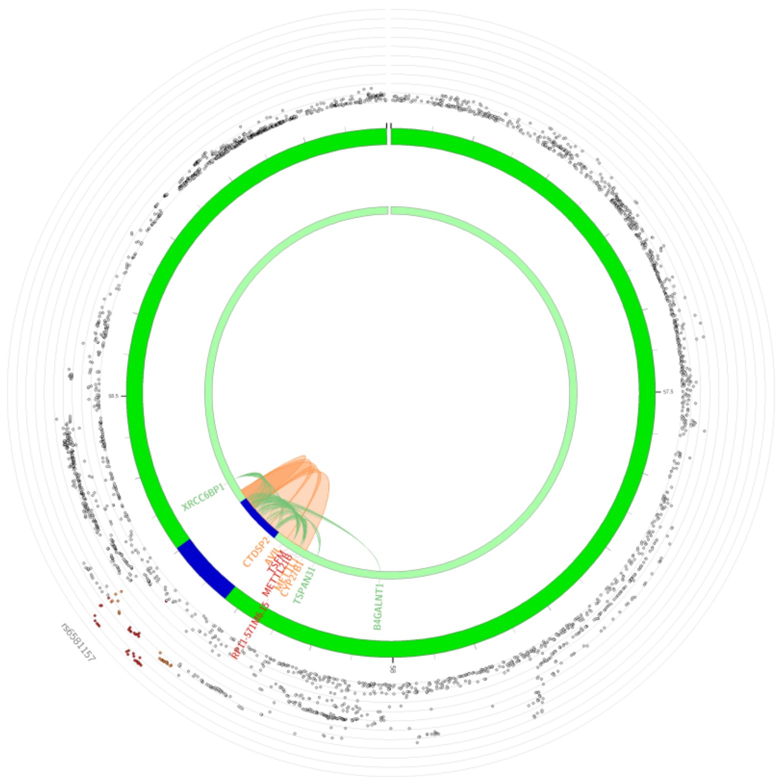

Chromosome 12

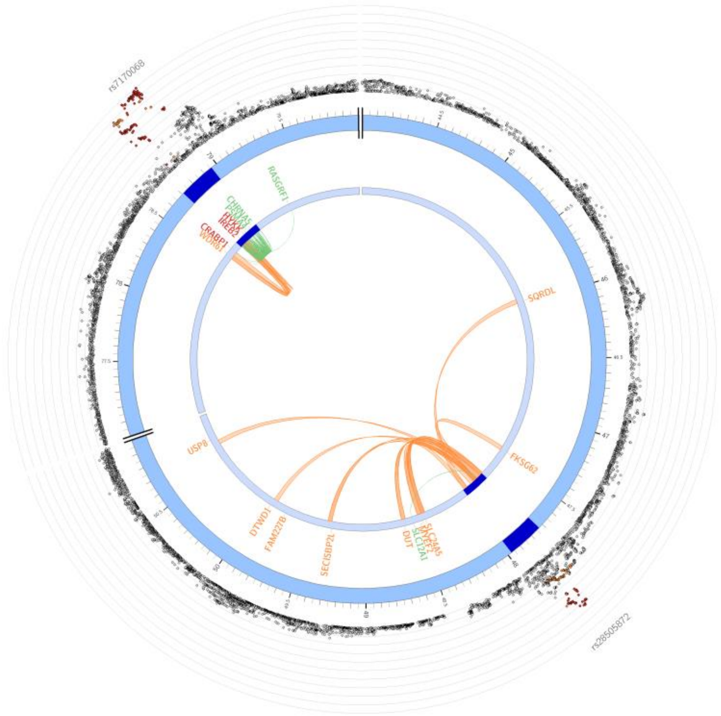

Chromosome 15

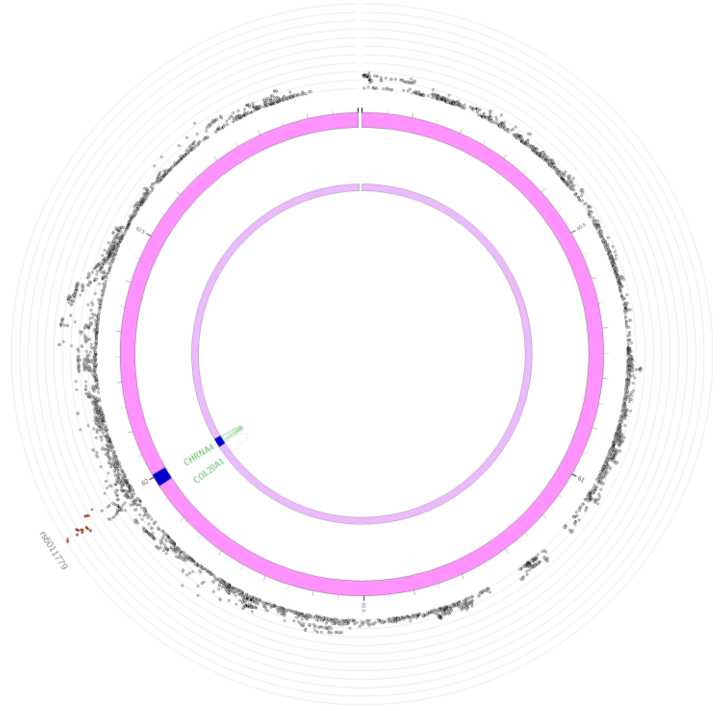

Chromosome 20

**Supplementary Figure 22. Significant SNPs identified in TUD Independent GWAS**
The chromosome is depicted as a circle with SNPs plotted by their -log10(p-value), and lead SNPs annotated. Orange links indicate chromatin contact, green links indicate cis-eQTLs, and red links indicate both.

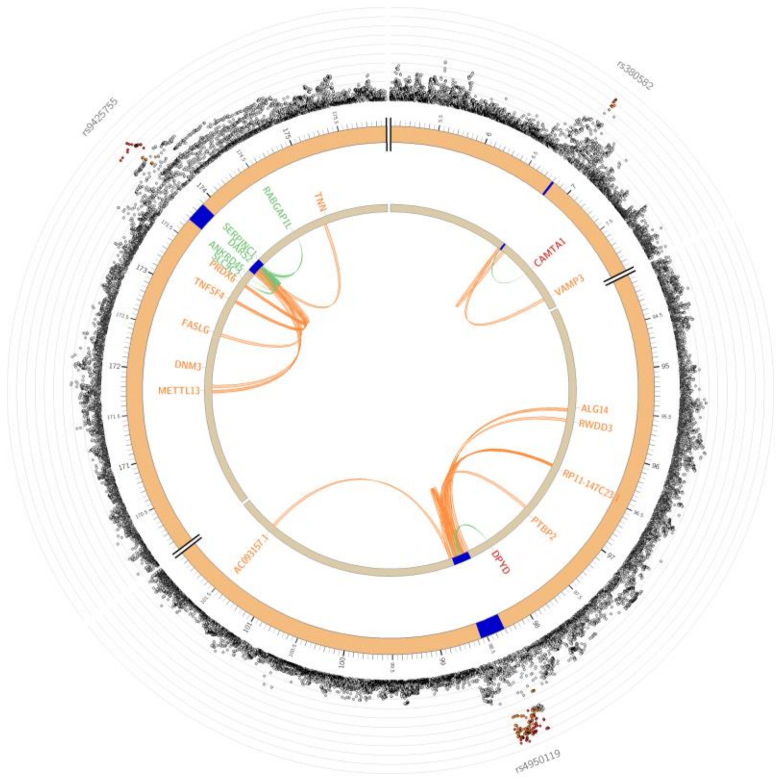

Chromosome 1

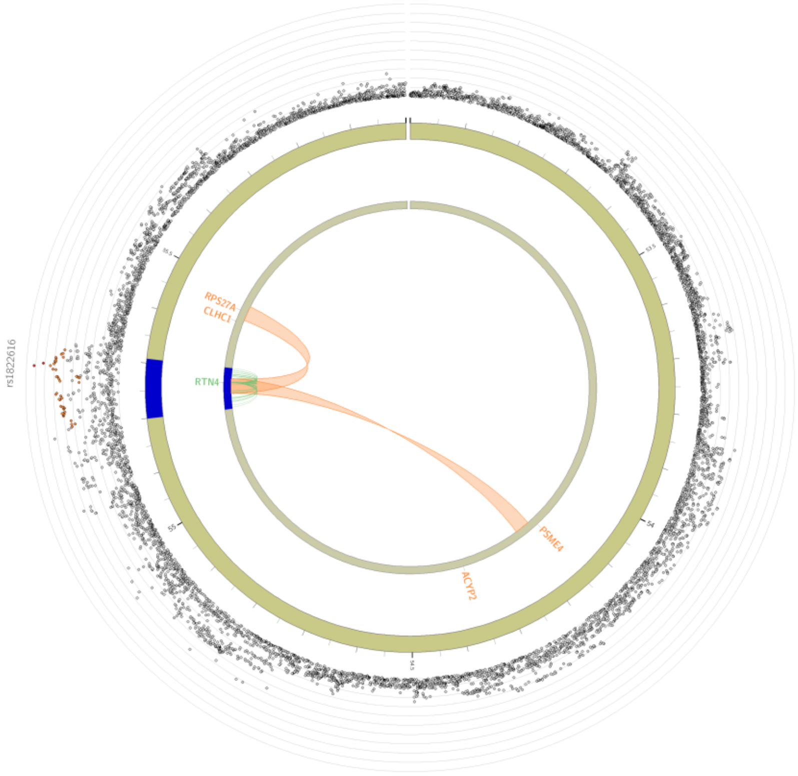

Chromosome 2

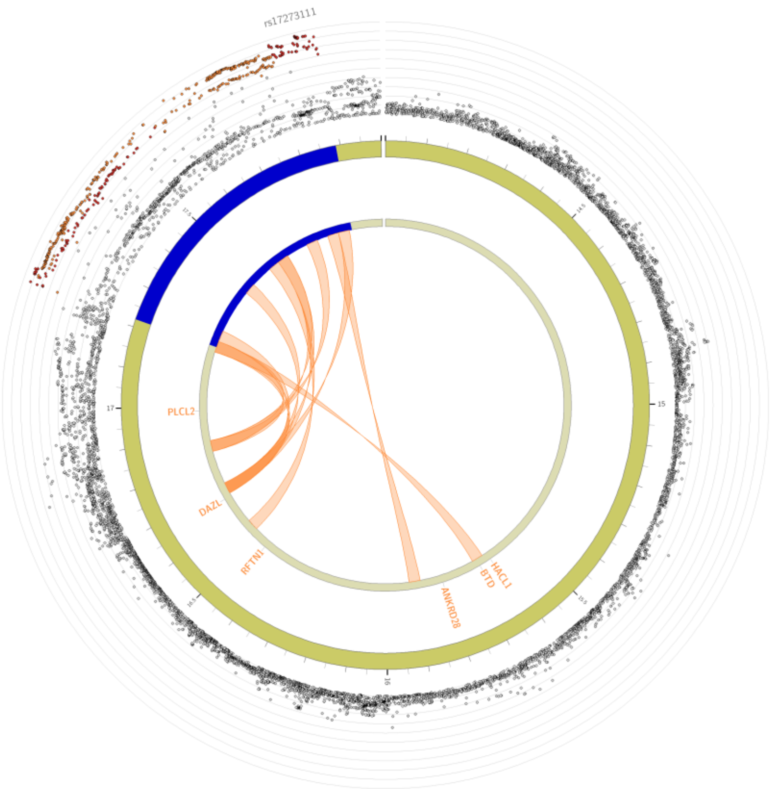

Chromosome 3

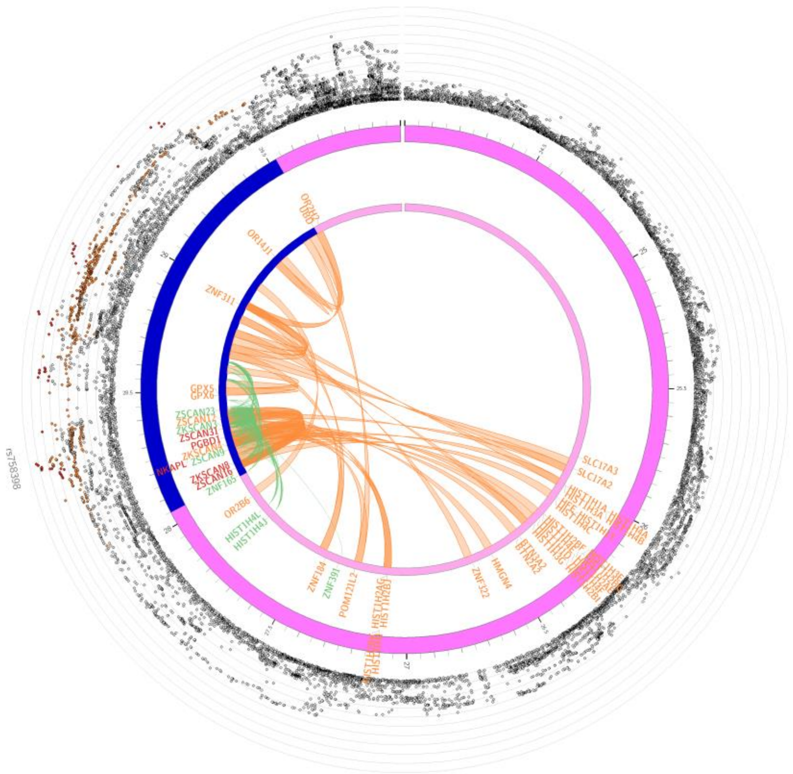

Chromosome 6

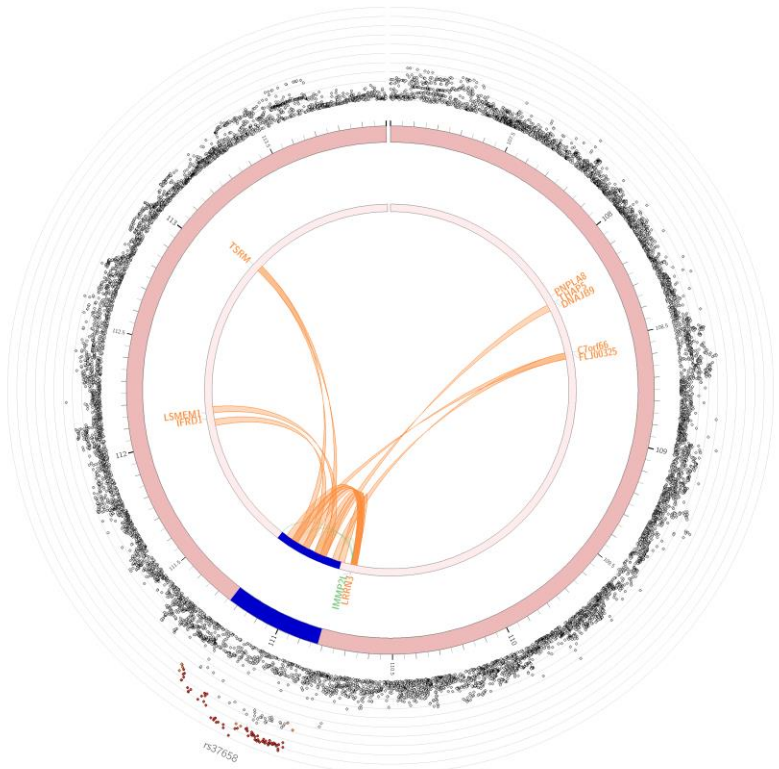

Chromosome 7

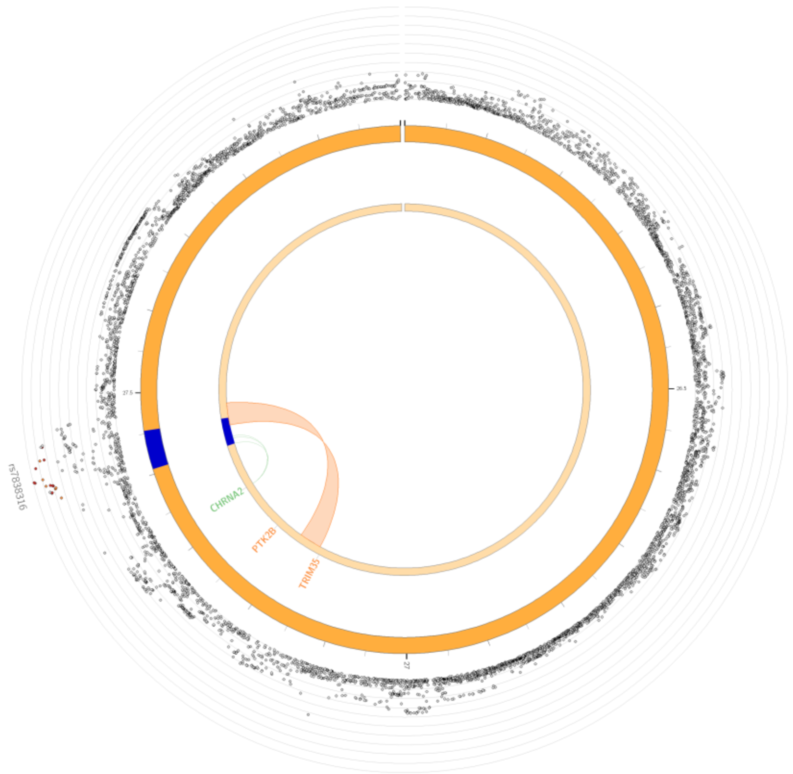

Chromosome 8

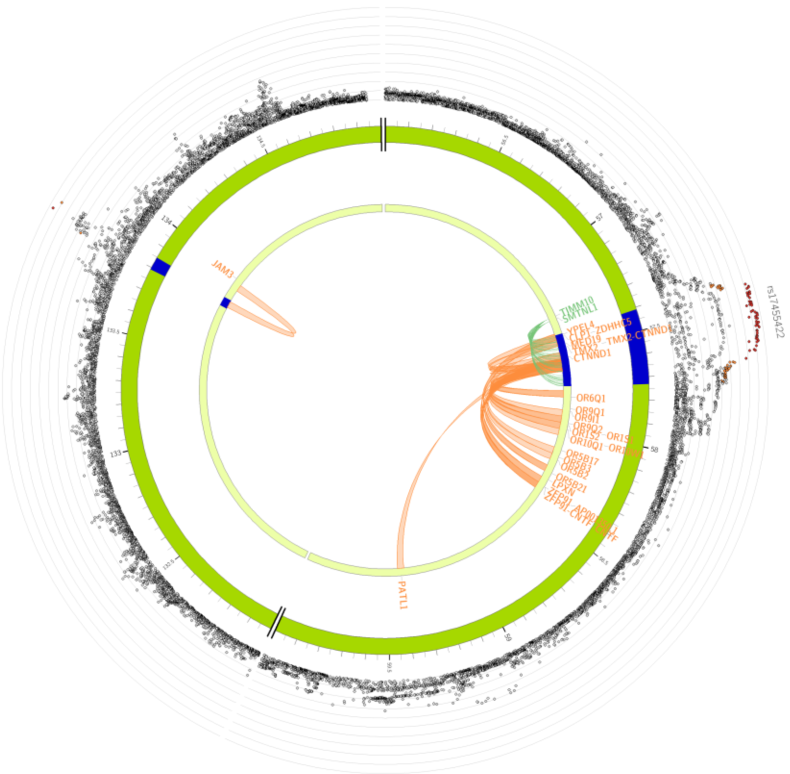

Chromosome 11

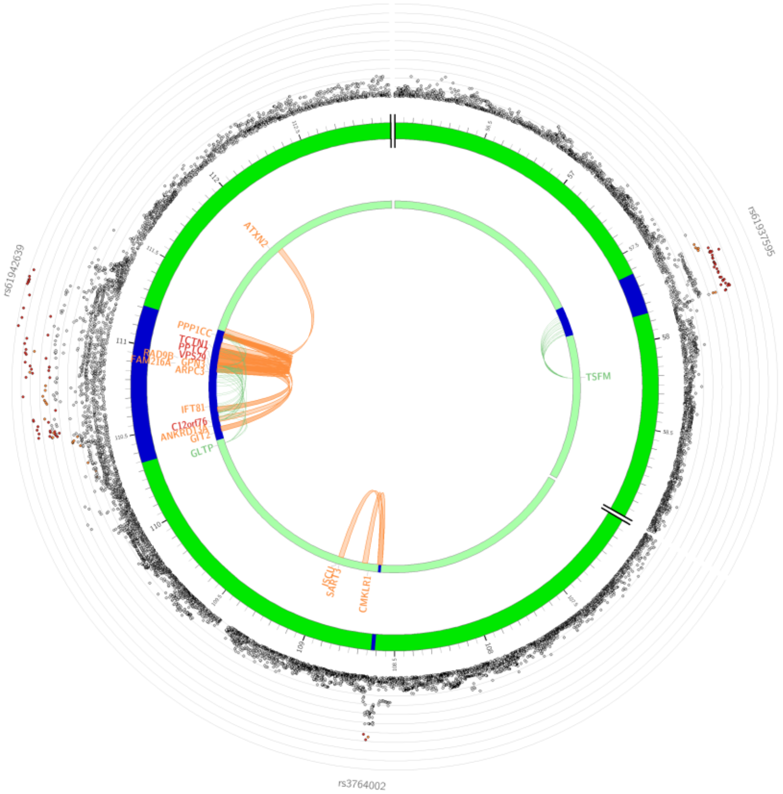

Chromosome 12

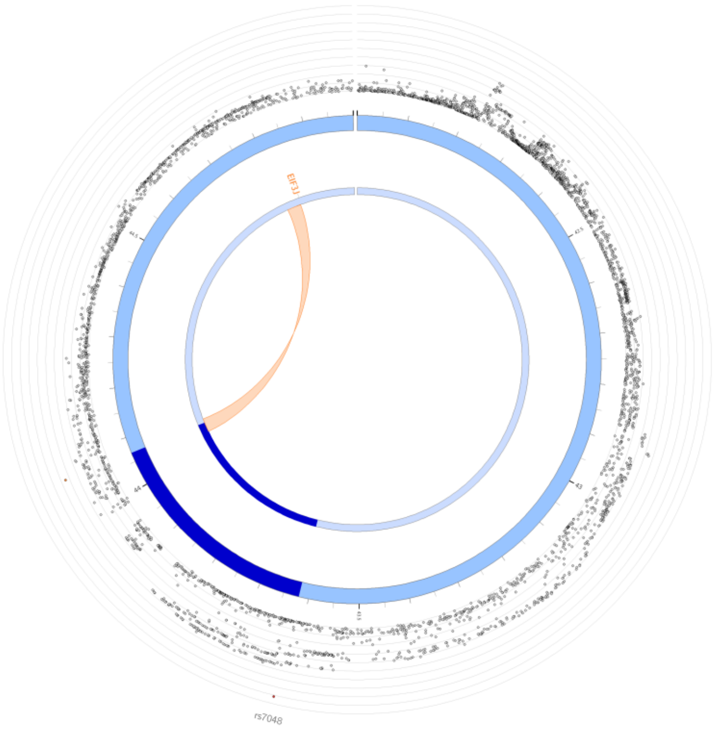

Chromosome 15

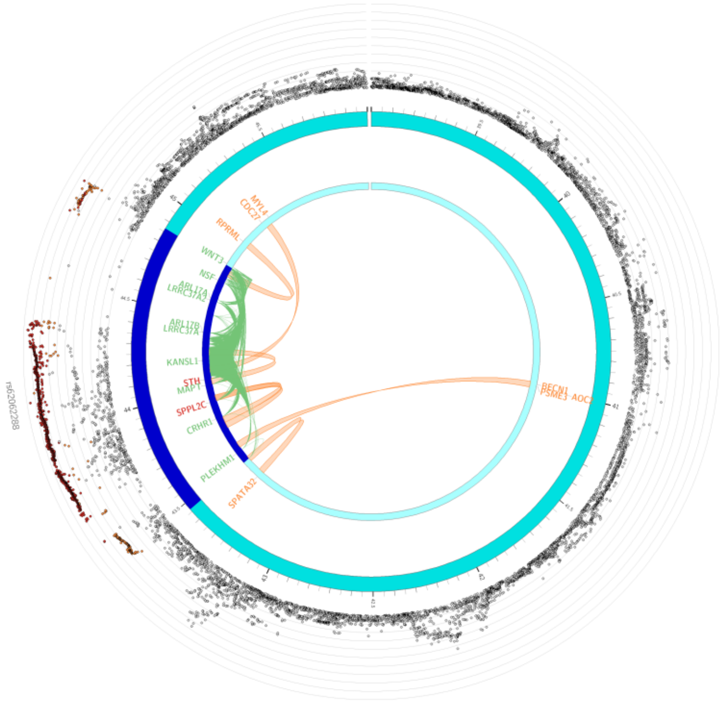

Chromosome 17

**Supplementary Figure 23. Significant SNPs identified in SCZ Independent GWAS**
The chromosome is depicted as a circle with SNPs plotted by their -log10(p-value), and lead SNPs annotated. Orange links indicate chromatin contact, green links indicate cis-eQTLs, and red links indicate both.

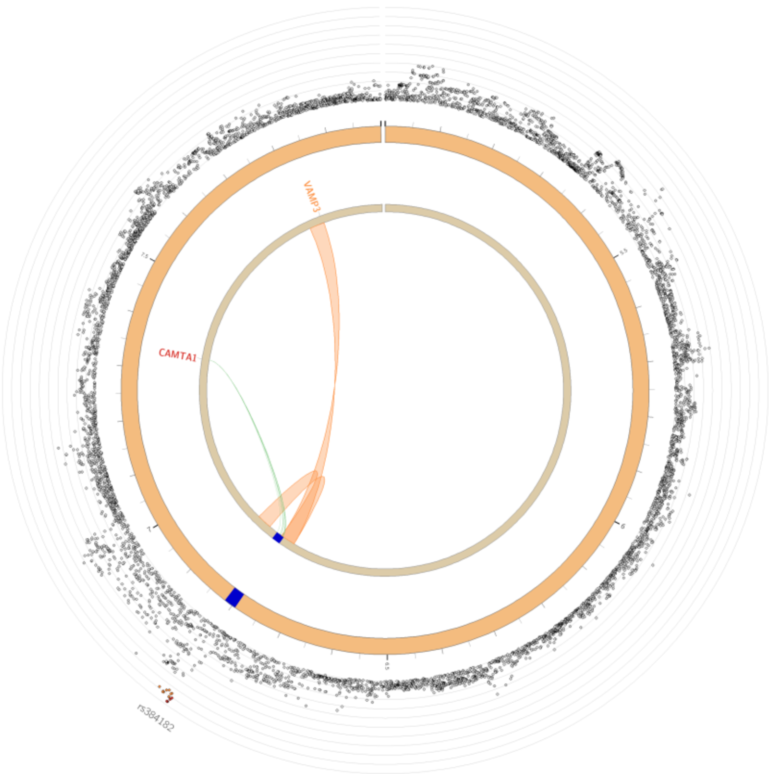

Chromosome 1

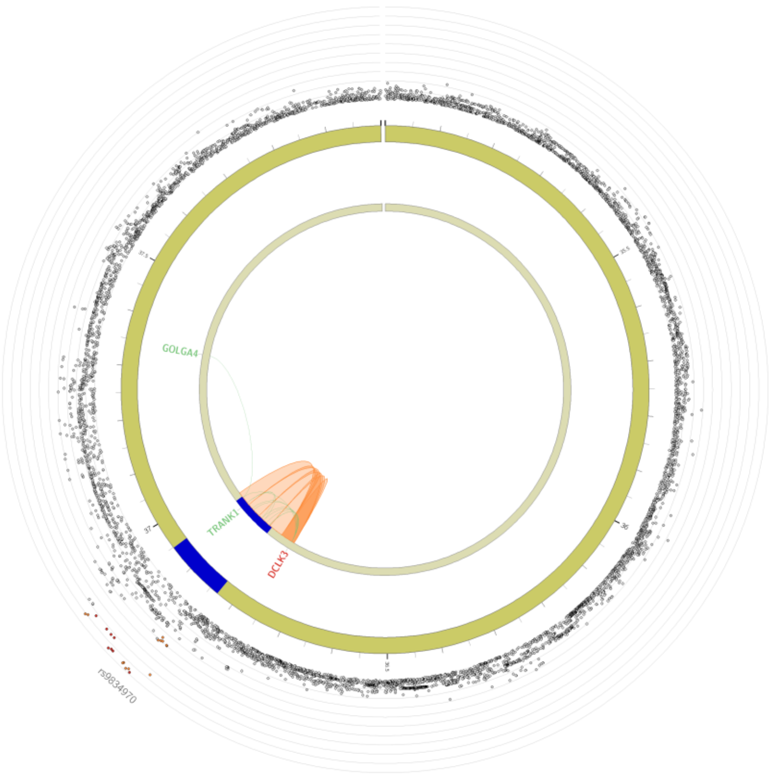

Chromosome 3

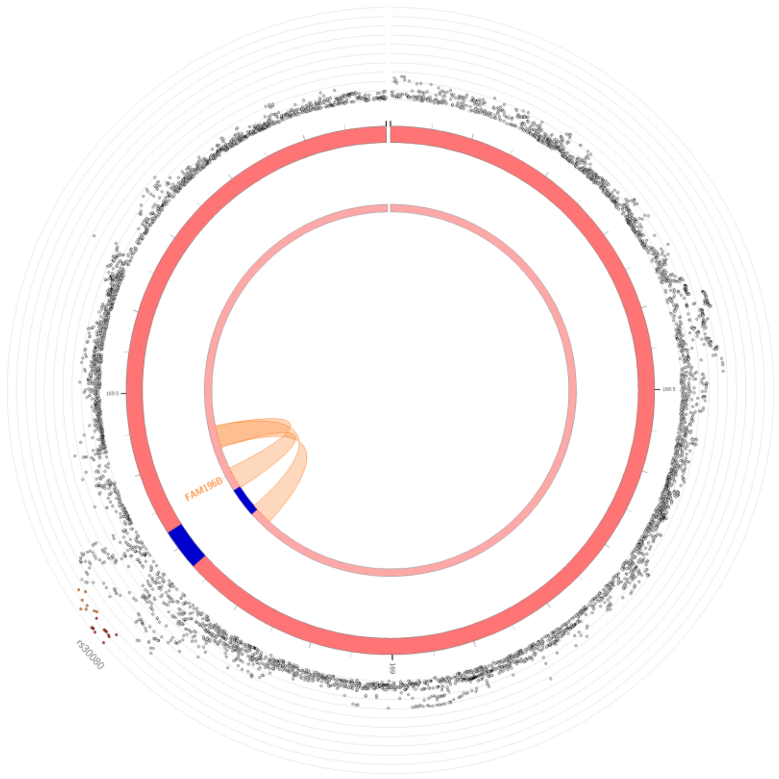

Chromosome 5

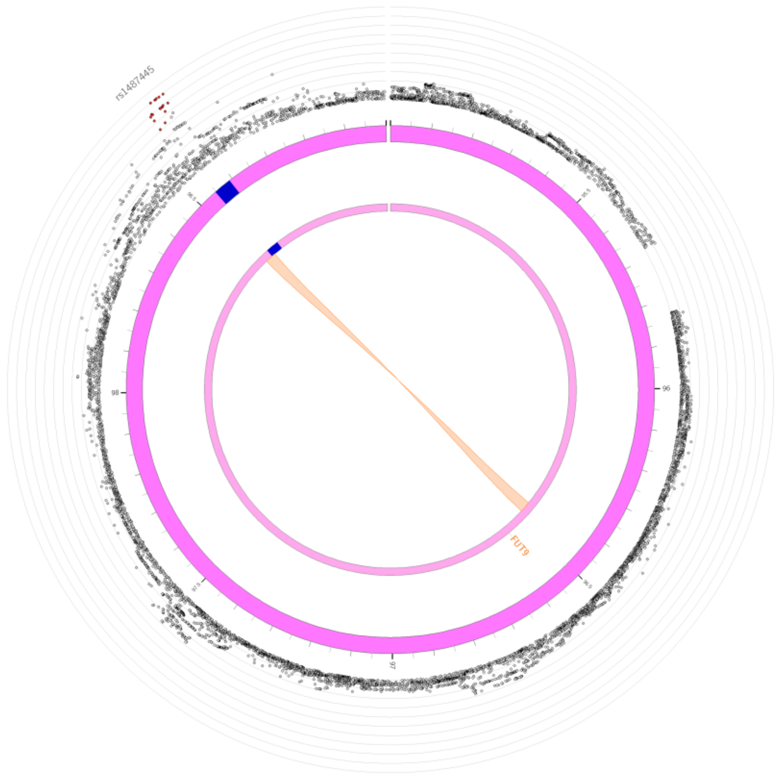

Chromosome 6

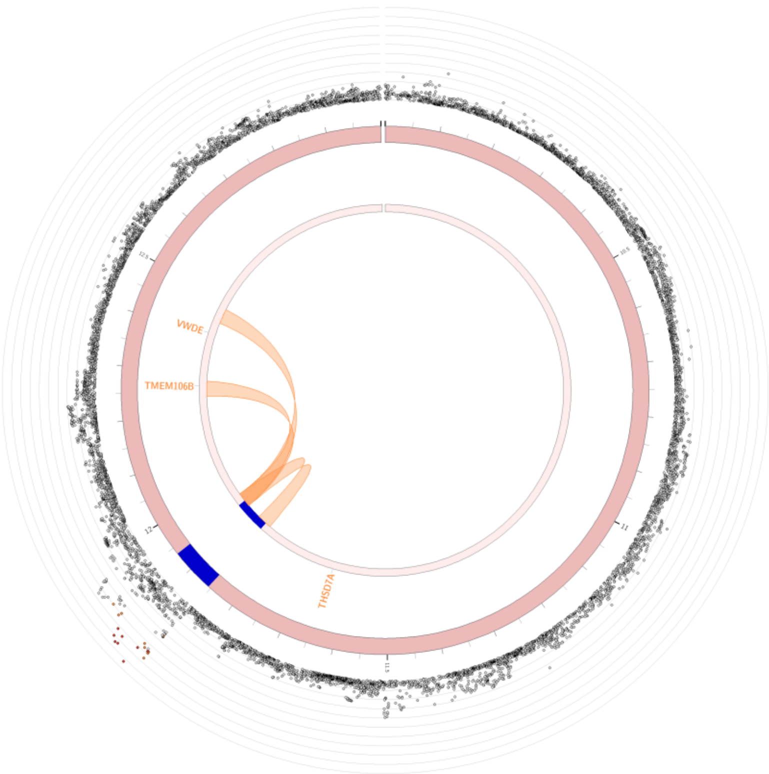

Chromosome 7

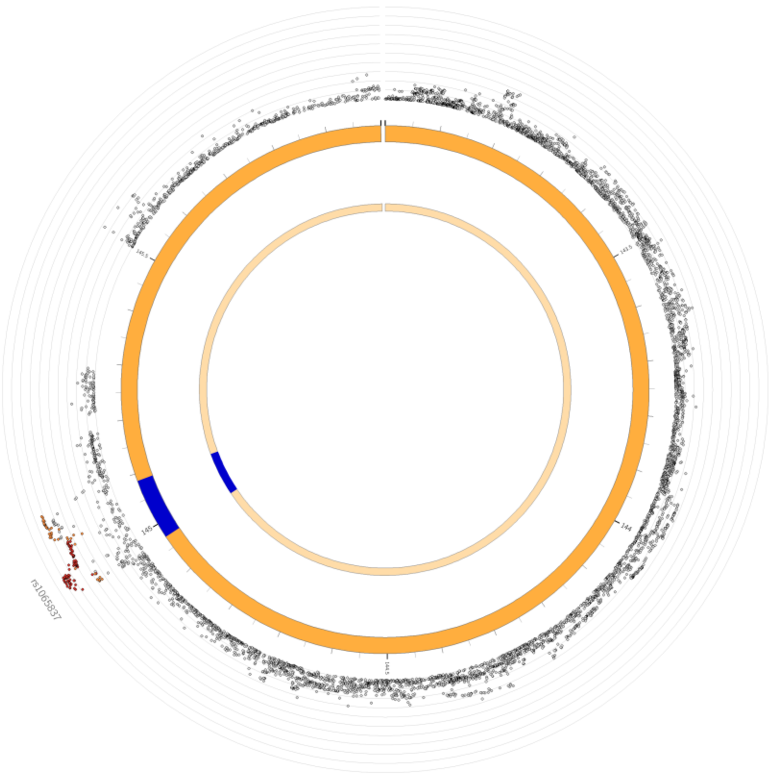

Chromosome 8

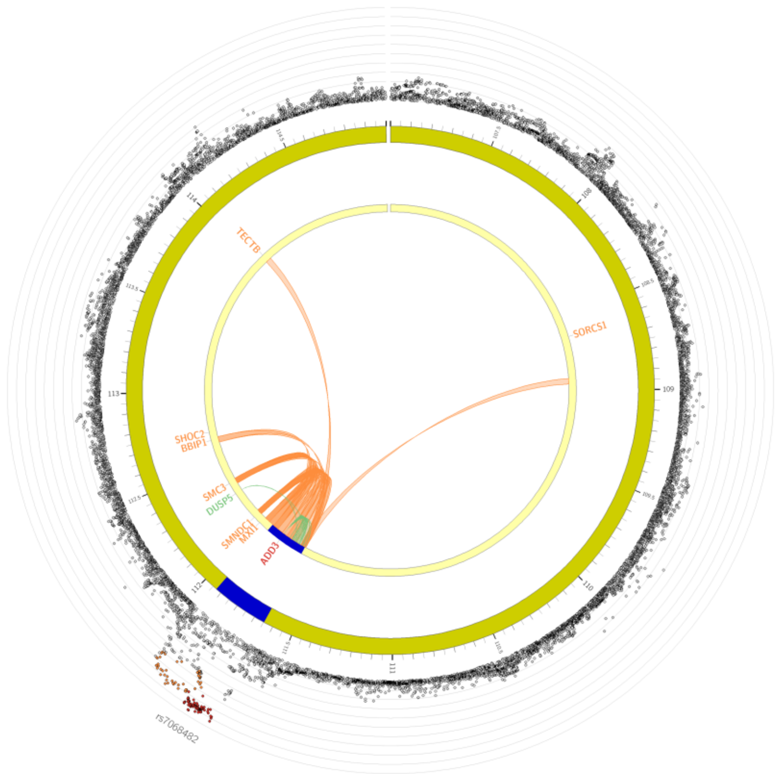

Chromosome 10

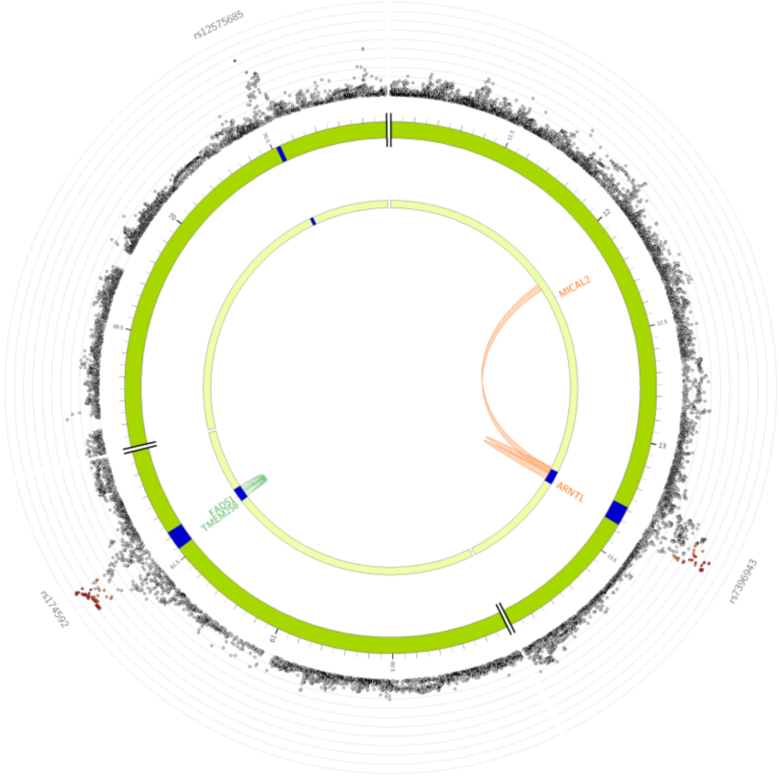

Chromosome 11

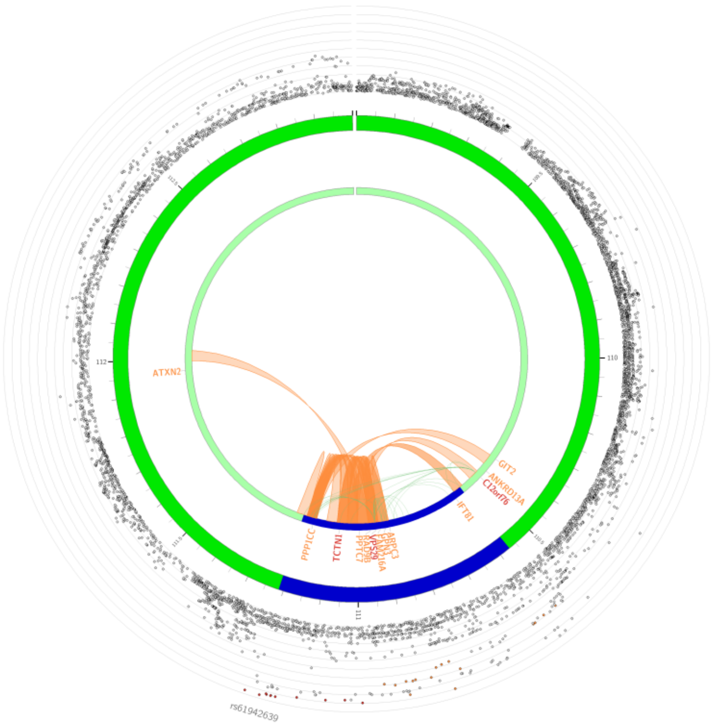

Chromosome 12

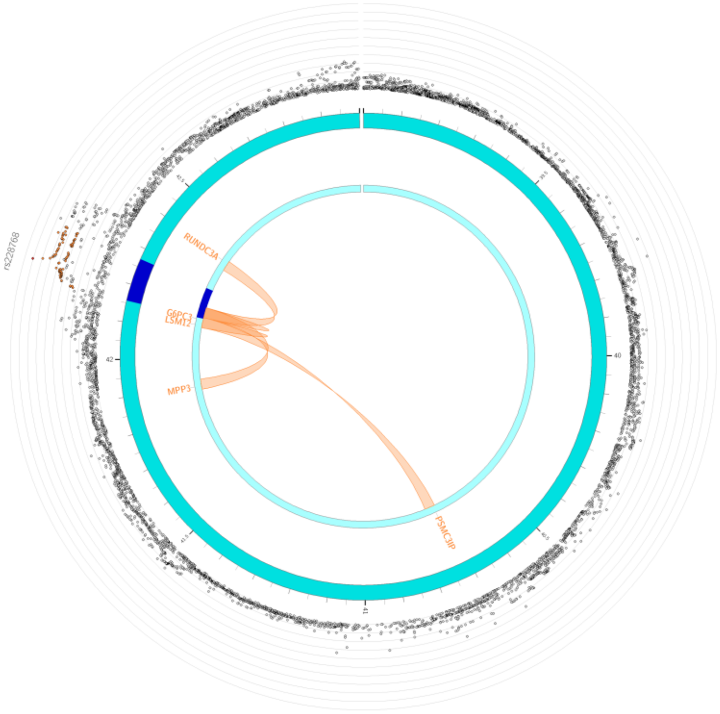

Chromosome 17

**Supplementary Figure 24. Significant SNPs identified in BD Independent GWAS**
The chromosome is depicted as a circle with SNPs plotted by their -log10(p-value), and lead SNPs annotated. Orange links indicate chromatin contact, green links indicate cis-eQTLs, and red links indicate both.

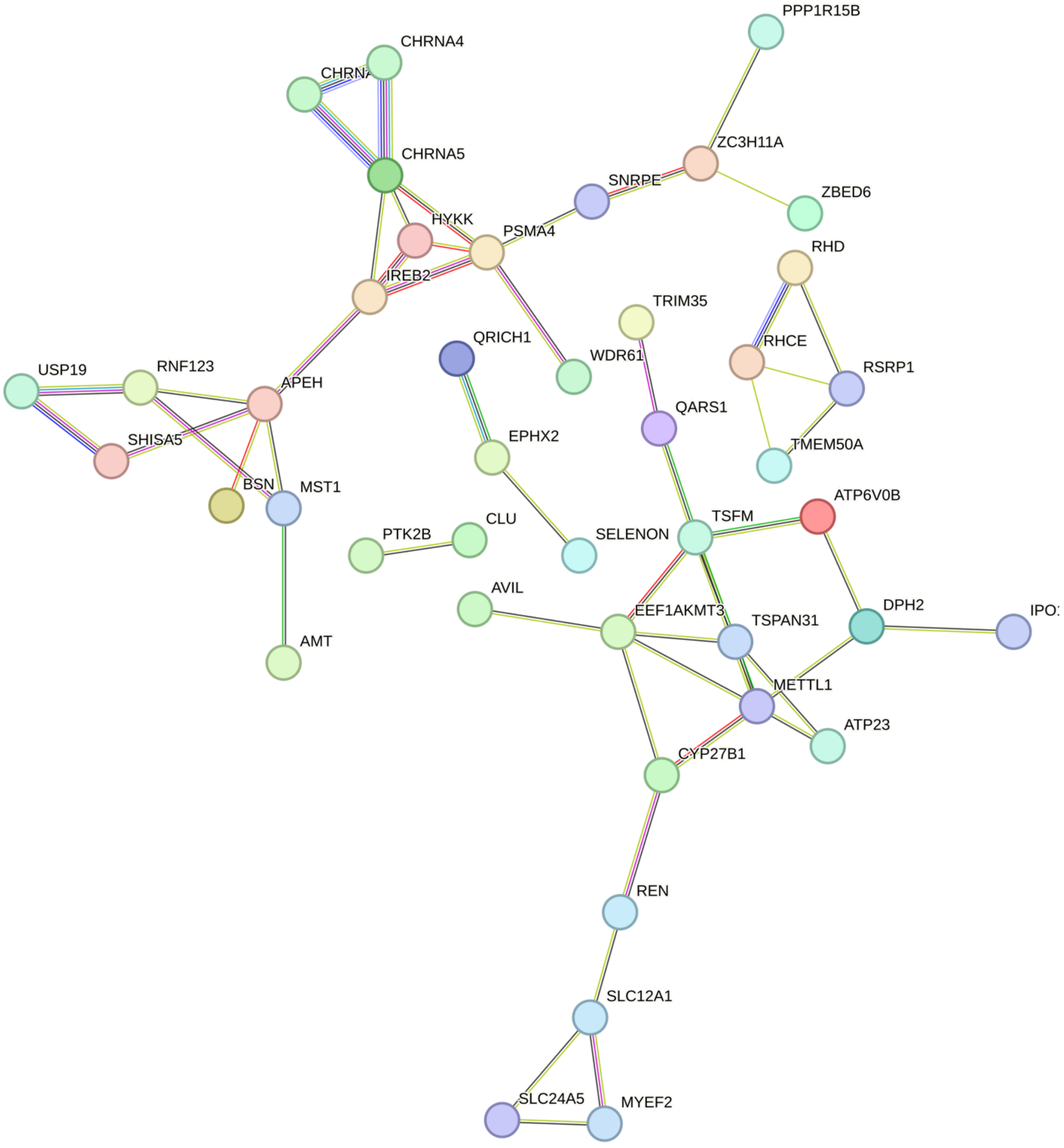

**Supplementary Figure 25. Protein-protein interaction network plot for TUD Independent.** Network plot was generated using STRING database v12.0. Nodes represent proteins, and edges represent protein-protein associations. Blue and pink edges represent known interactions, while green, red, and blue represent predicted interactions.

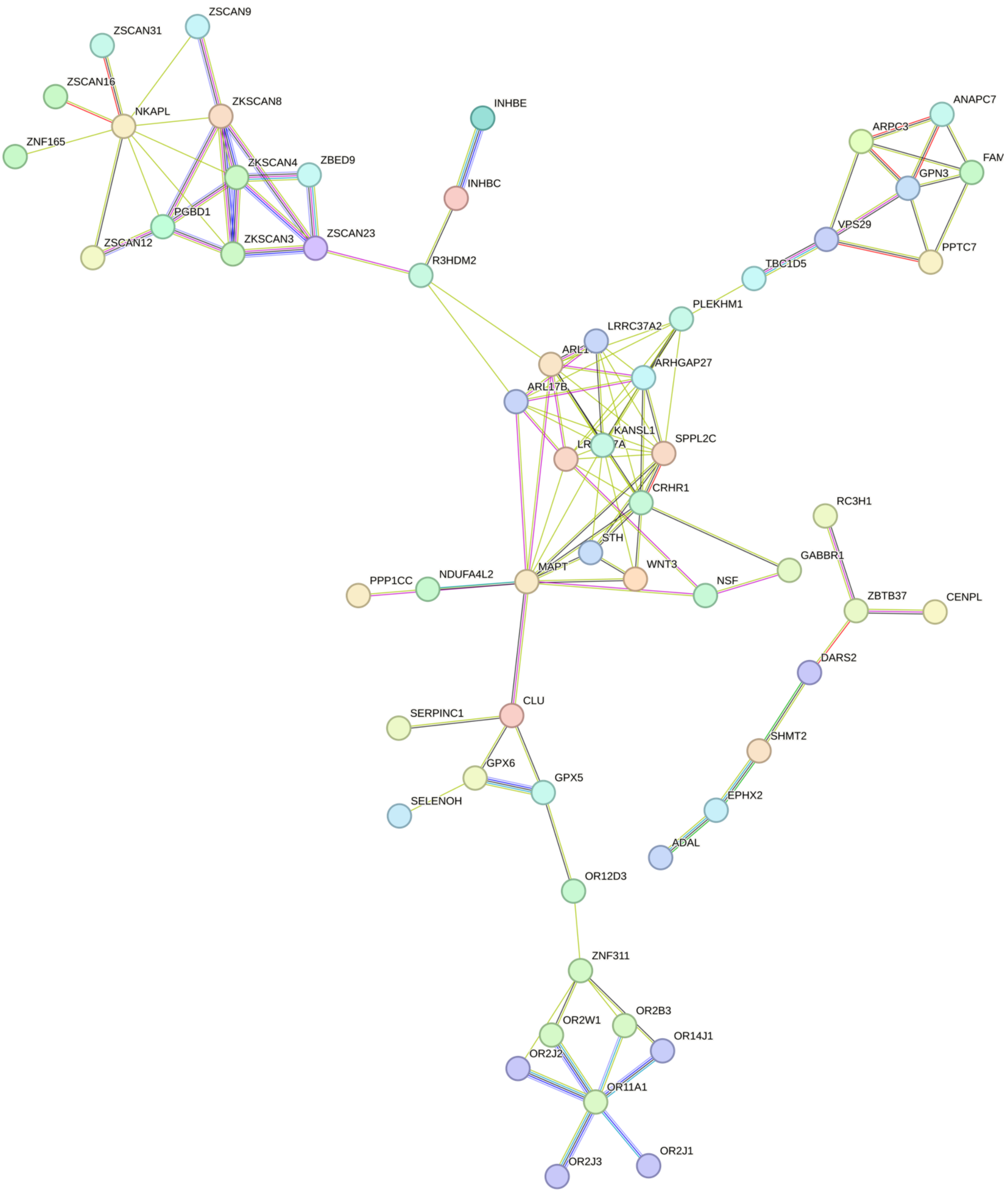

**Supplementary Figure 26. Protein-protein interaction network plot for SCZ Independent.** Network plot was generated using STRING database v12.0. Nodes represent proteins, and edges represent protein-protein associations. Blue and pink edges represent known interactions, while green, red, and blue represent predicted interactions.

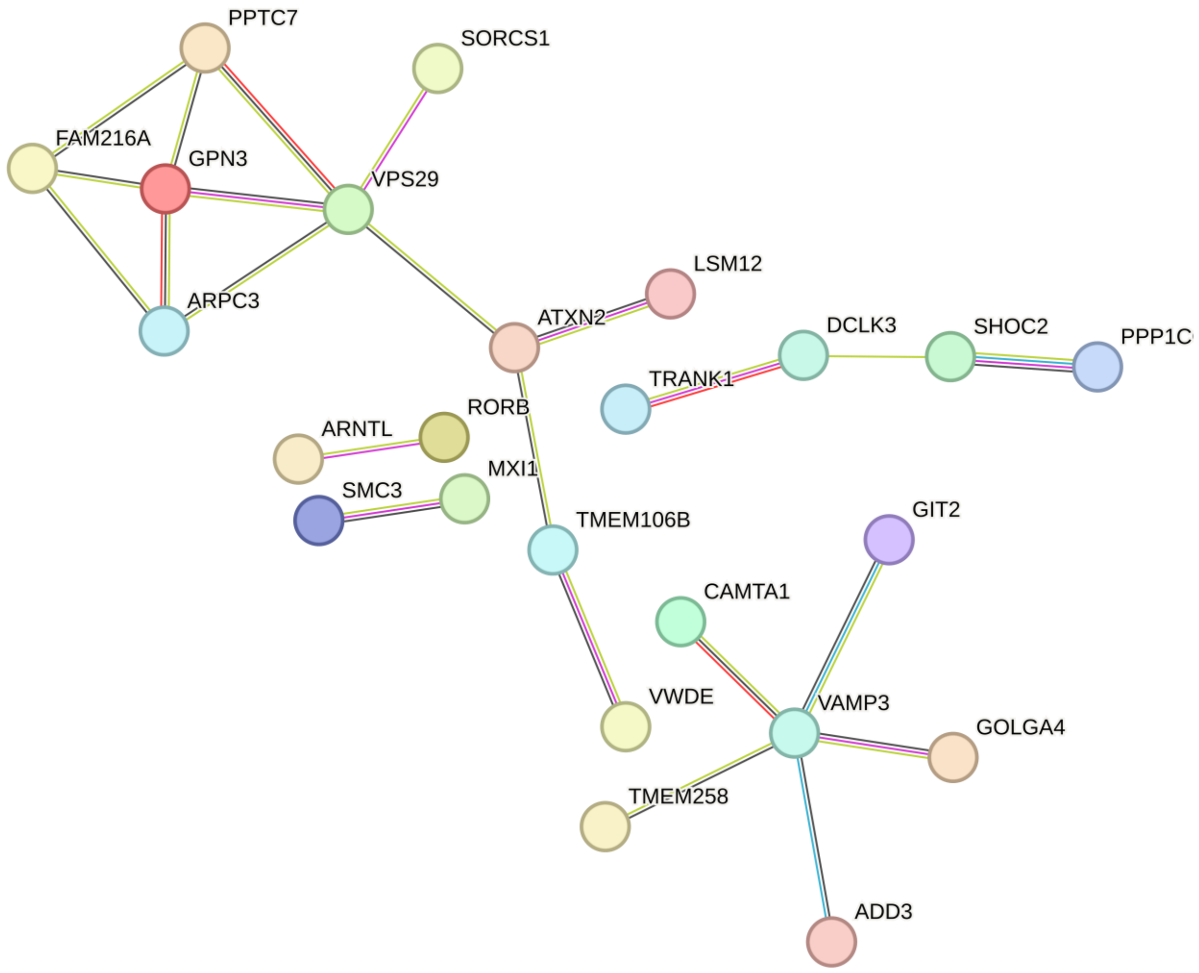

**Supplementary Figure 27. Protein-protein interaction network plot for BD Independent.** Network plot was generated using STRING database v12.0. Nodes represent proteins, and edges represent protein-protein associations. Blue and pink edges represent known interactions, while green, red, and blue represent predicted interactions.

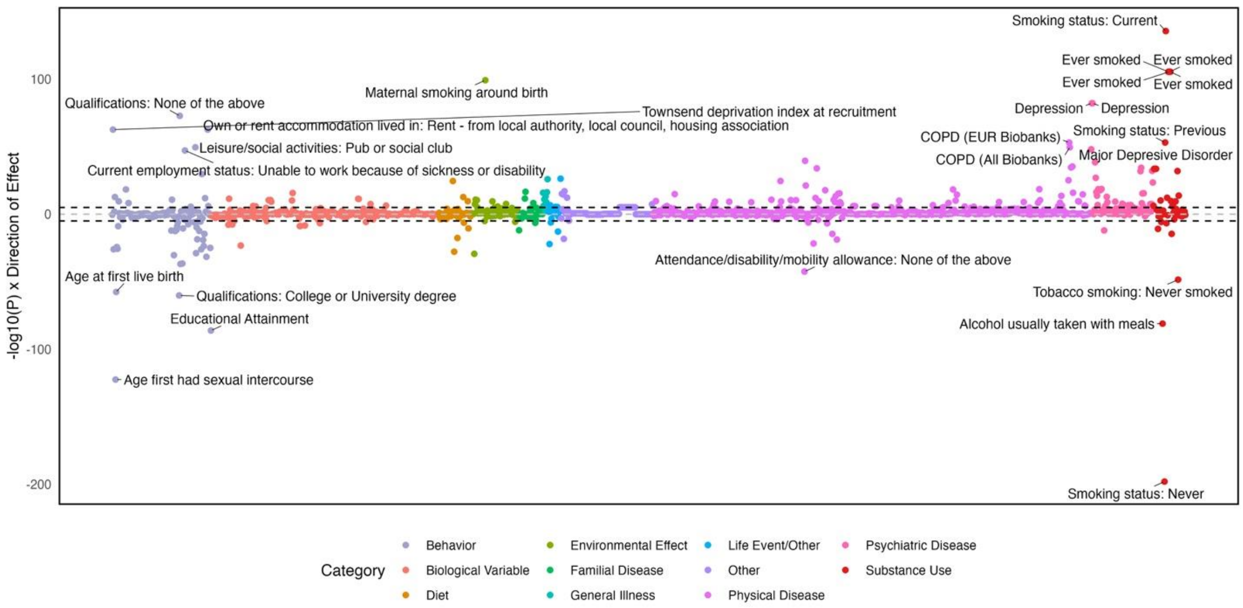

**Supplementary Figure 28. Genetic correlation results for the EUR substance use disorders factor**

The top 25 associations are shown. Assocation analyses were performed using the MASSIVE pipeline.

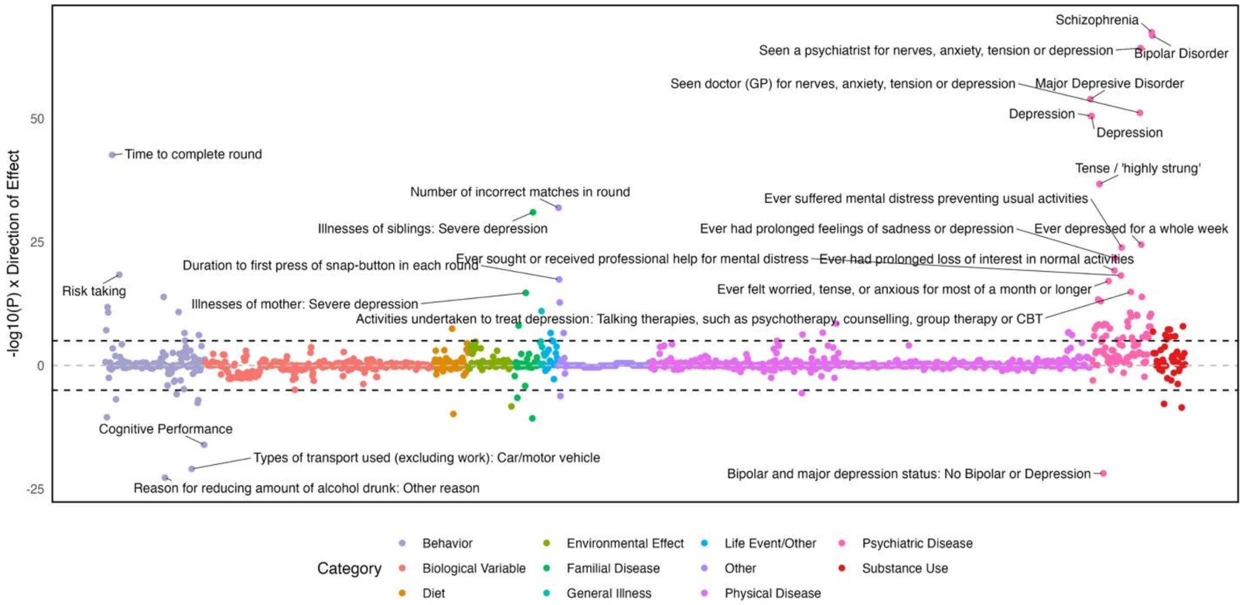

**Supplementary Figure 29. Genetic correlation results for the EUR psychotic disorders factor**

The top 25 associations are shown. Assocation analyses were performed using the MASSIVE pipeline.

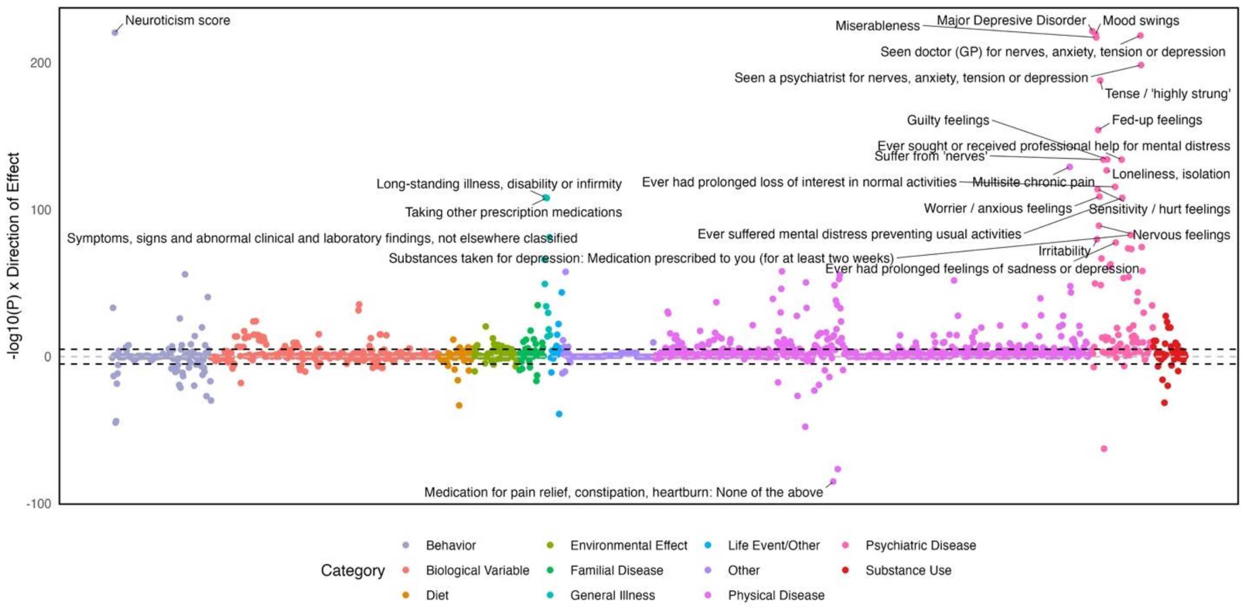

**Supplementary Figure 30. Genetic correlation results for the EUR mood disorders factor**
The top 25 associations are shown. Assocation analyses were performed using the MASSIVE pipeline.

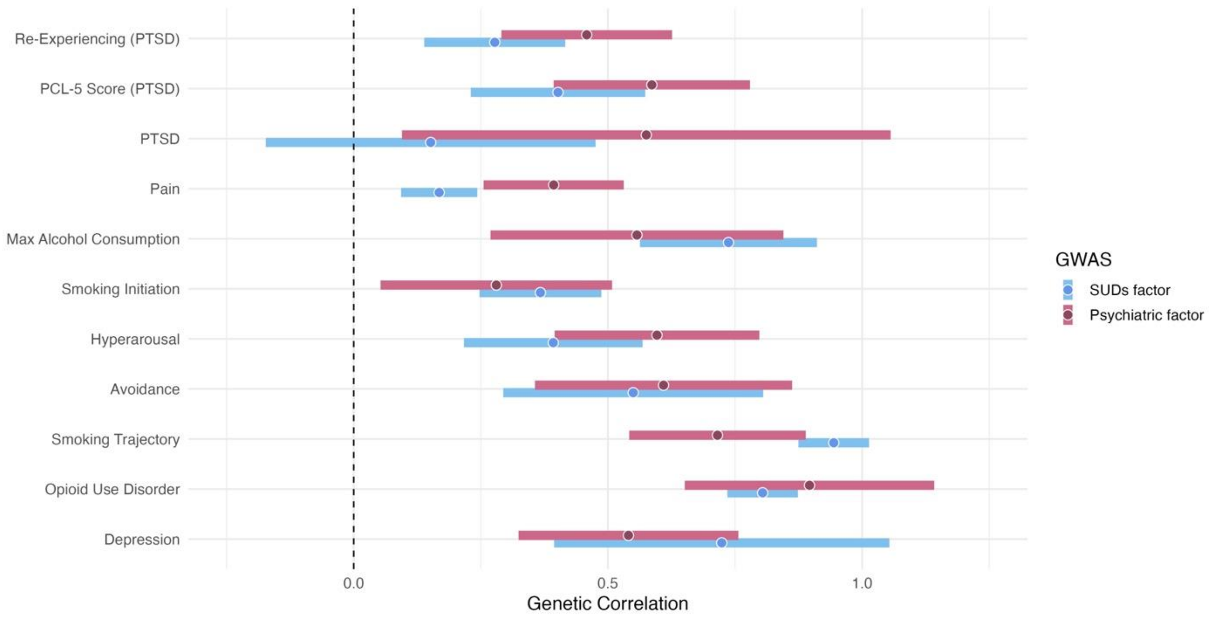

**Supplementary Figure 31. Genetic correlations between AFR ancestry common factors and psychiatric and substance use phenotypes**

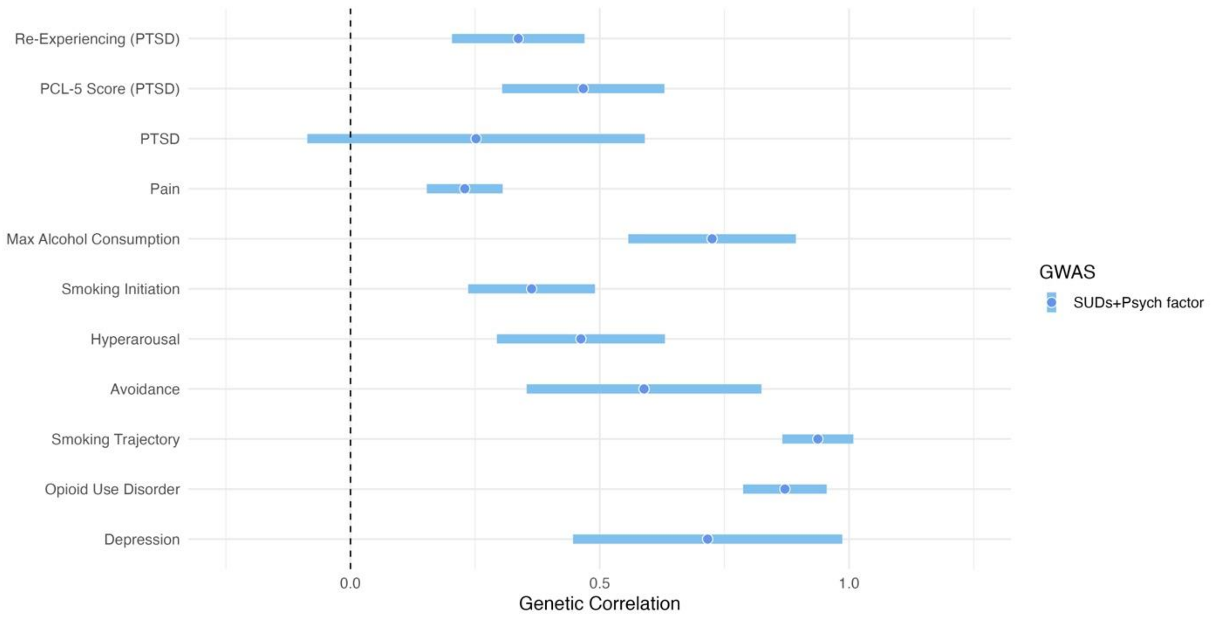

**Supplementary Figure 32. Genetic correlations between the AFR ancestry second-order common factor and psychiatric and substance use traits**

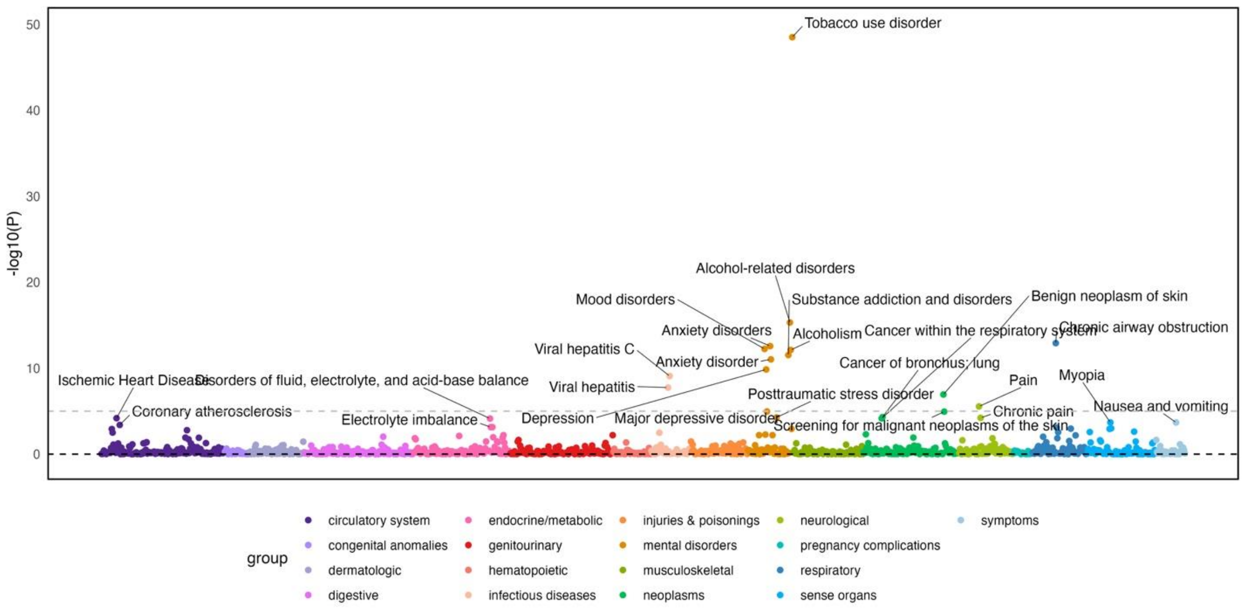

**Supplementary Figure 33. PheWAS results for the EUR substance use disorders factor in Penn Medicine BioBank**

The top 25 associations are shown. All p-values were adjusted using Benjamini-Hochberg false discovery rate (FDR) correction.

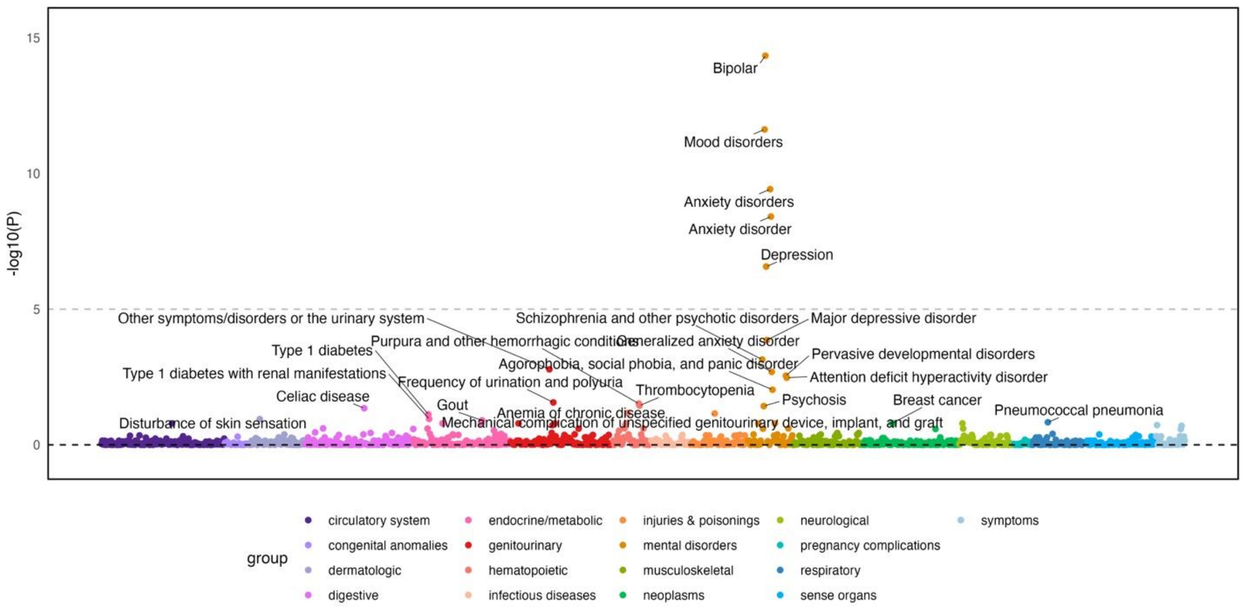

**Supplementary Figure 34. PheWAS results for the EUR psychotic disorders factor in Penn Medicine BioBank**

The top 25 associations are shown. All p-values were adjusted using Benjamini-Hochberg false discovery rate (FDR) correction.

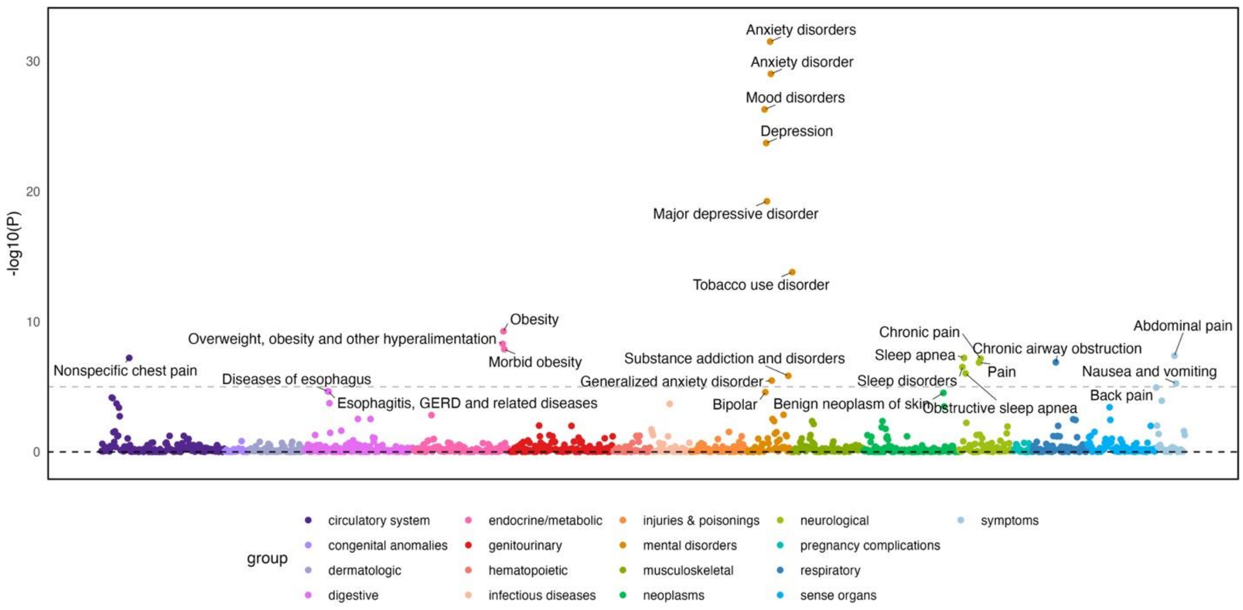

**Supplementary Figure 35. PheWAS results for the EUR mood disorders factor in Penn Medicine BioBank**

The top 25 associations are shown. All p-values were adjusted using Benjamini-Hochberg false discovery rate (FDR) correction.

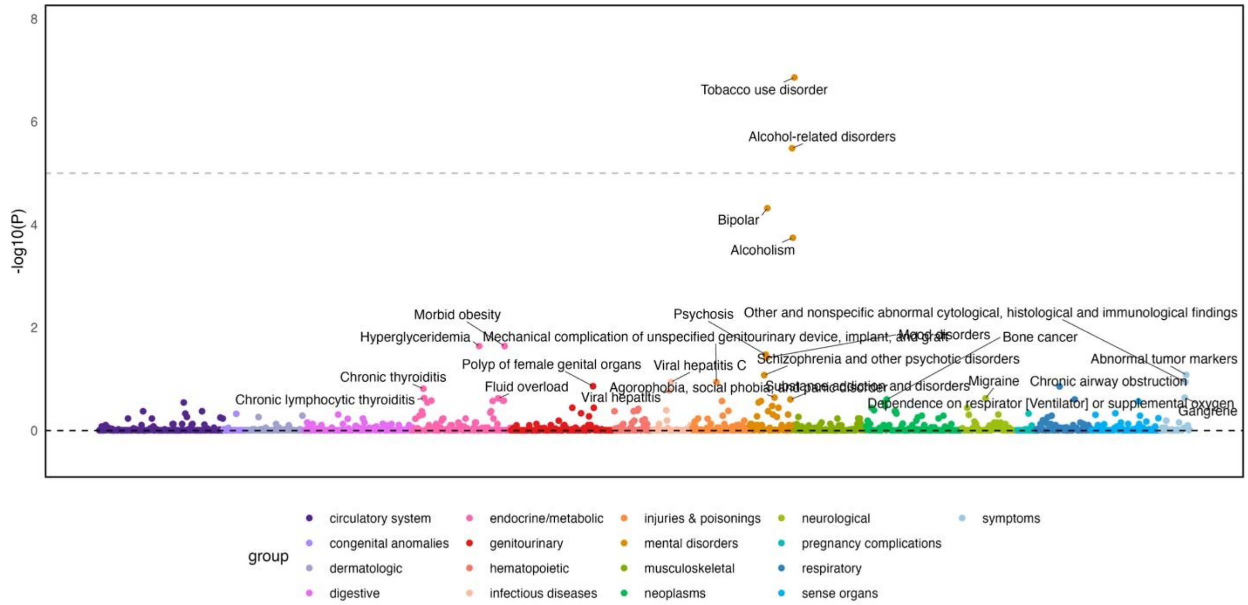

**Supplementary Figure 36. PheWAS results for EUR ancestry second-order common factor representing overlap in substance use and psychotic disorders in Penn Medicine BioBank**
The top 25 associations are shown. All p-values were adjusted using Benjamini-Hochberg false discovery rate (FDR) correction.

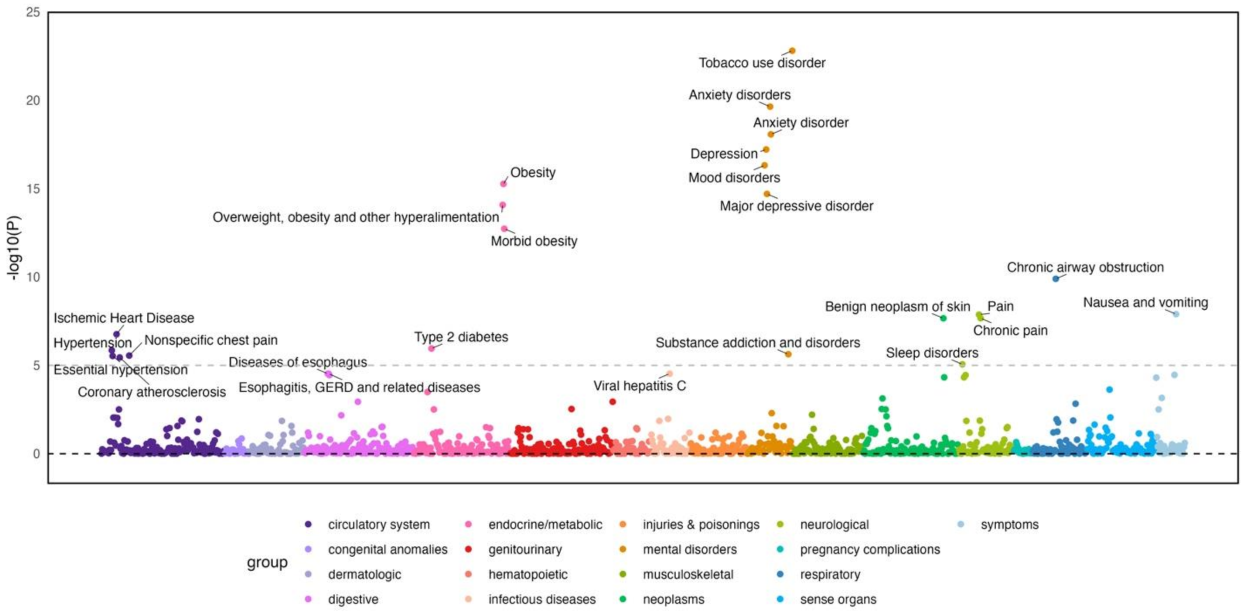

**Supplementary Figure 37. PheWAS results for EUR ancestry second-order common factor representing overlap in substance use and mood/anxiety disorders in Penn Medicine BioBank**

The top 25 associations are shown. All p-values were adjusted using Benjamini-Hochberg false discovery rate (FDR) correction.

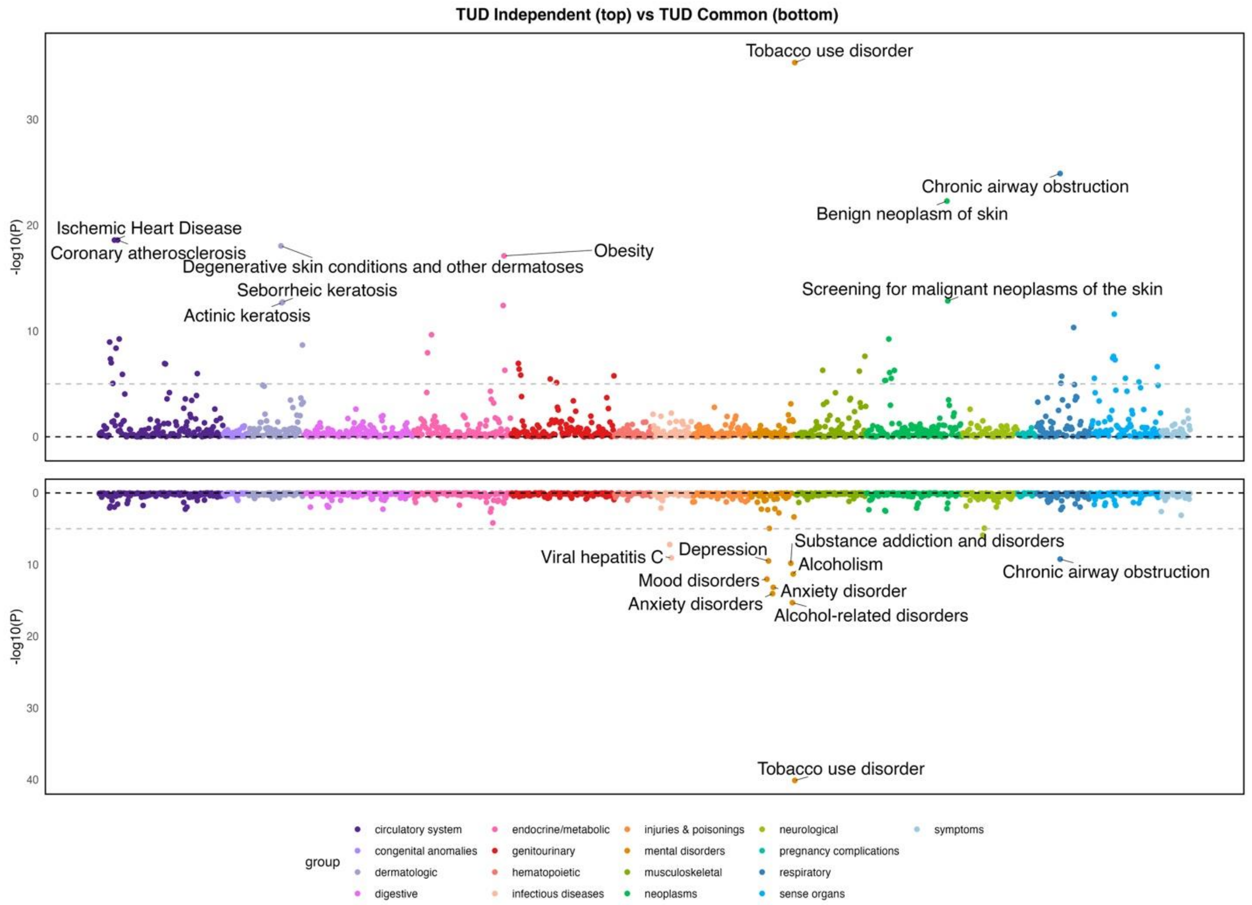

**Supplementary Figure 38. Hudson plot of PheWAS results for tobacco use disorders GWAS-by-subtraction in Penn Medicine BioBank**

The top 10 associations for each GWAS are shown. All p-values were adjusted using Benjamini-Hochberg false discovery rate (FDR) correction.

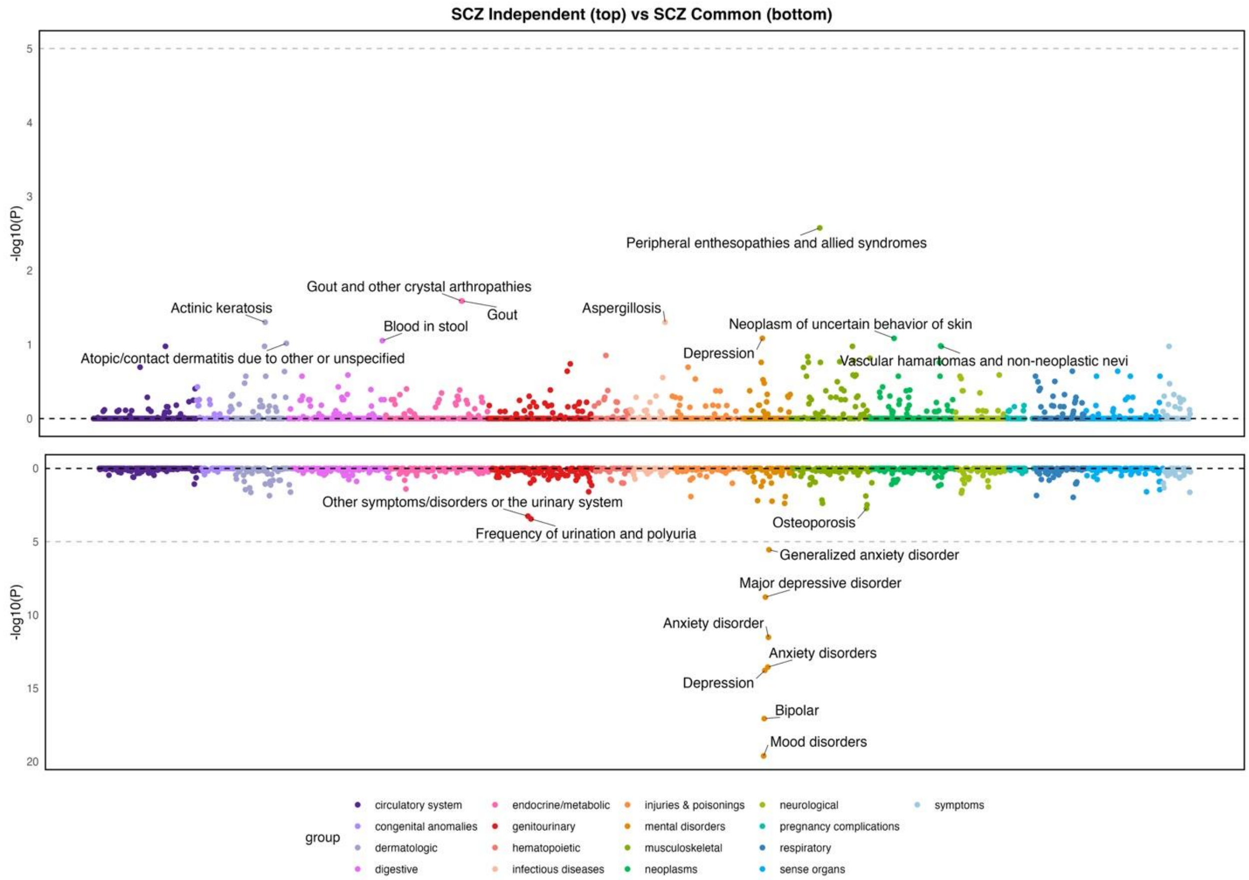

**Supplementary Figure 39. Hudson plot of PheWAS results for schizophrenia GWAS-by-subtraction in Penn Medicine BioBank**

The top 10 associations for each GWAS are shown. All p-values were adjusted using Benjamini-Hochberg false discovery rate (FDR) correction.

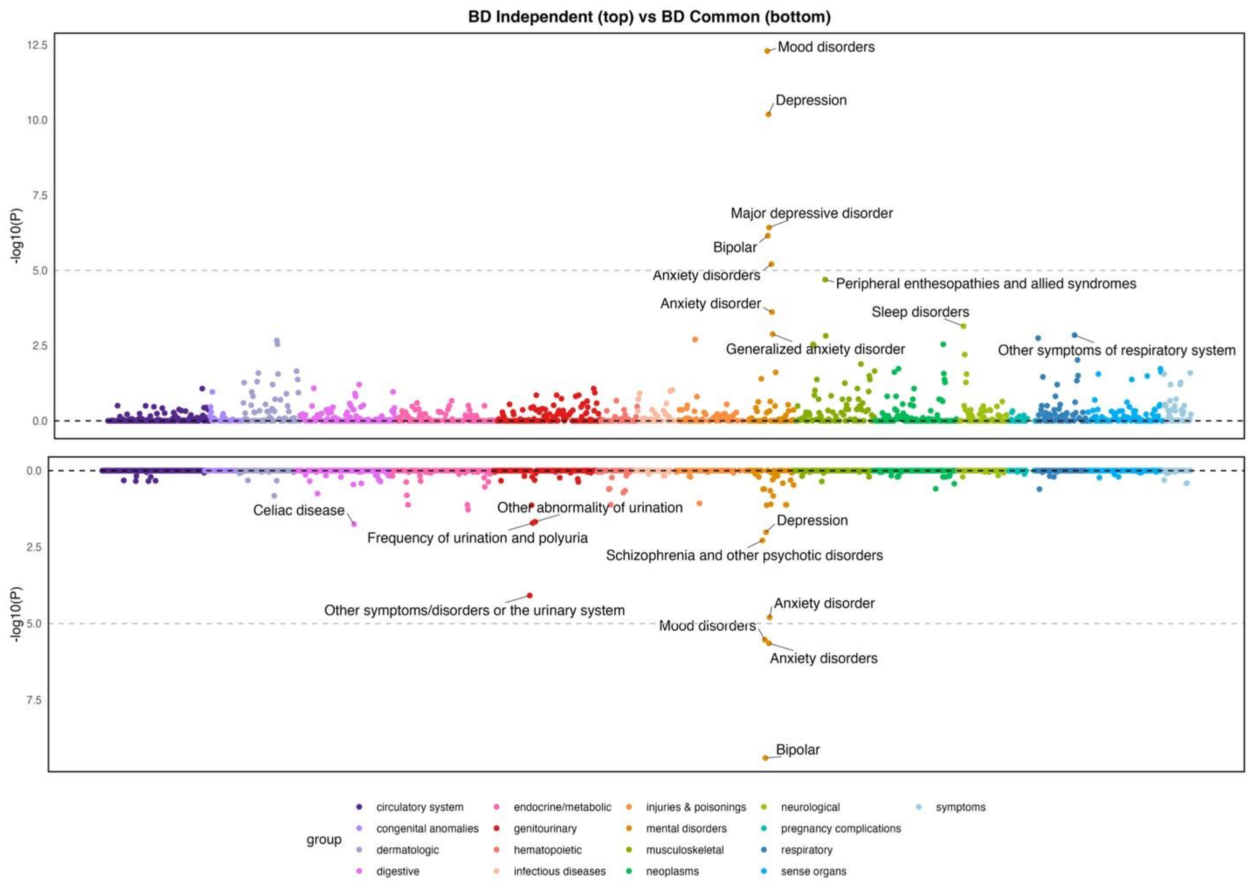

**Supplementary Figure 40. Hudson plot of PheWAS results for bipolar disorder GWAS-by-subtraction in Penn Medicine BioBank**

The top 10 associations for each GWAS are shown. All p-values were adjusted using Benjamini-Hochberg false discovery rate (FDR) correction.

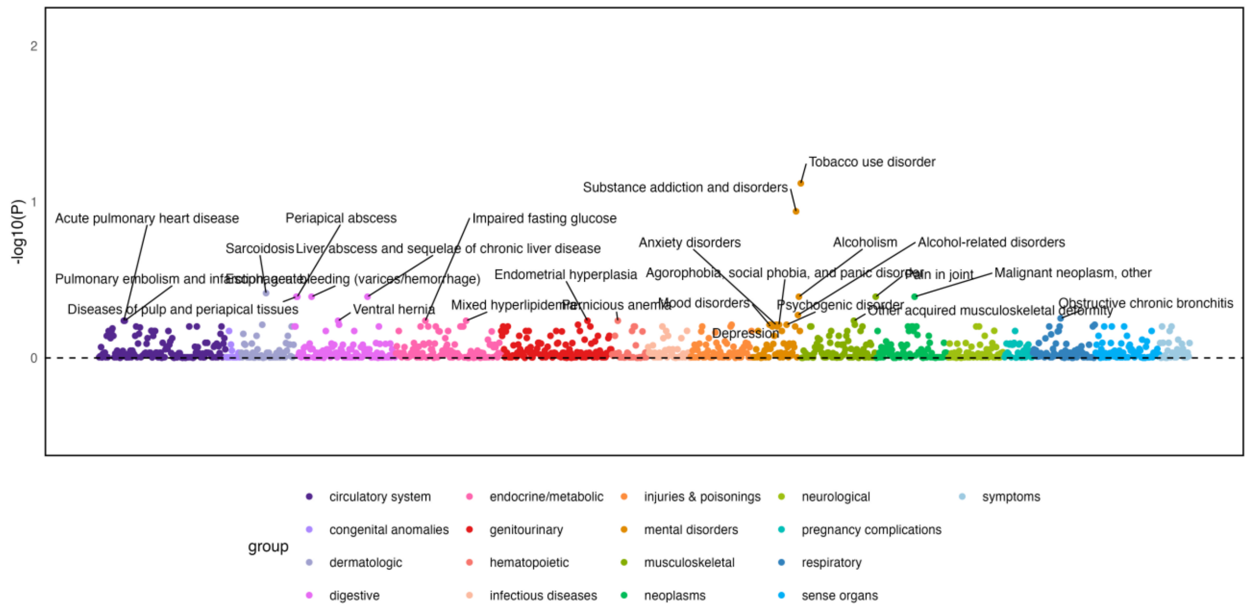

**Supplementary Figure 41. PheWAS results for AFR ancestry substance use disorders factor in Penn Medicine BioBank**

The top 25 associations are shown. All p-values were adjusted using Benjamini-Hochberg false discovery rate (FDR) correction.

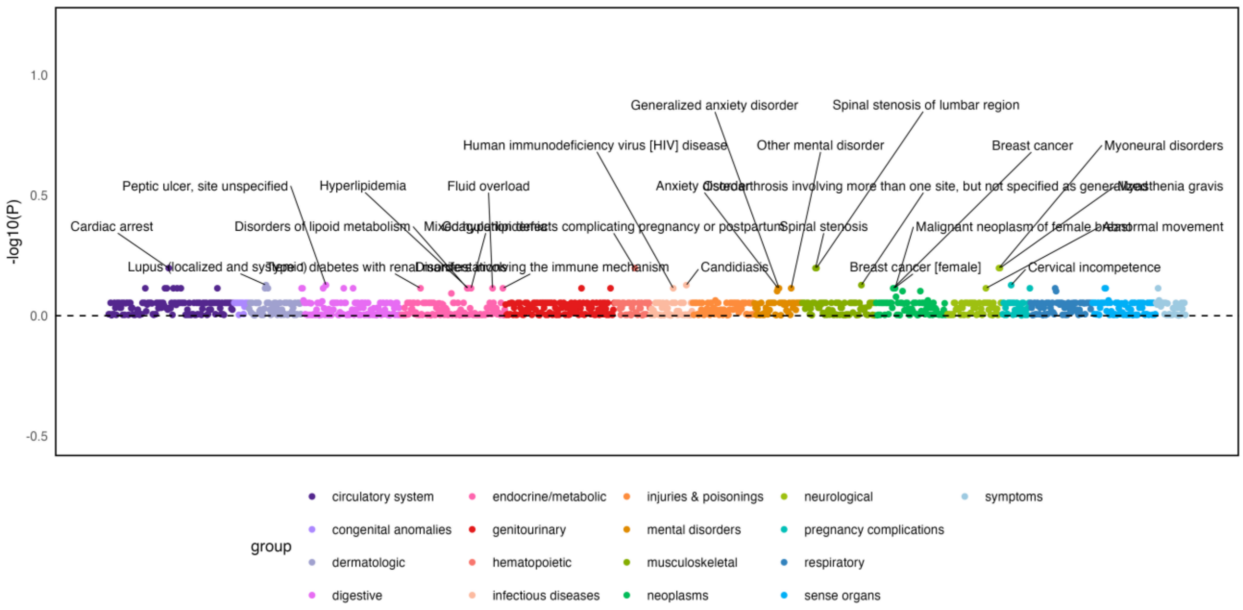

**Supplementary Figure 42. PheWAS results for AFR ancestry psychiatric disorders factor in Penn Medicine BioBank**

The top 25 associations are shown. All p-values were adjusted using Benjamini-Hochberg false discovery rate (FDR) correction.

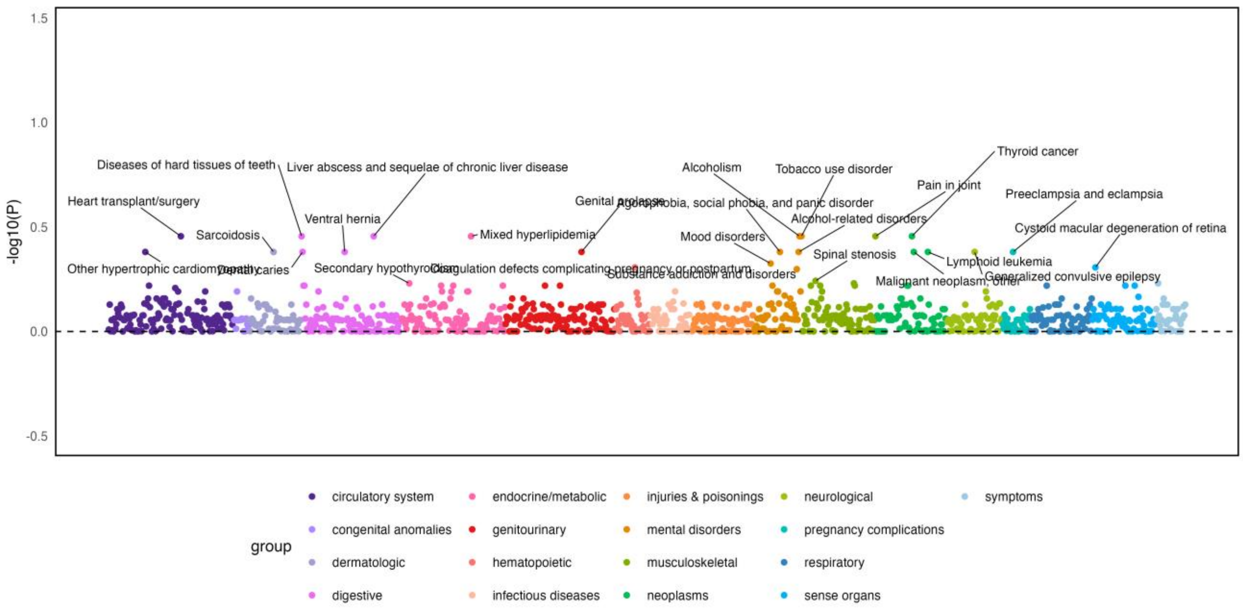

**Supplementary Figure 43. PheWAS results for AFR ancestry second-order common factor representing overlap in substance use and psychiatric disorders in Penn Medicine BioBank** The top 25 associations are shown. All p-values were adjusted using Benjamini-Hochberg false discovery rate (FDR) correction.
